# Mapping Dysfunctional Circuits in the Frontal Cortex Using Deep Brain Stimulation

**DOI:** 10.1101/2023.03.07.23286766

**Authors:** Barbara Hollunder, Jill L. Ostrem, Ilkem Aysu Sahin, Nanditha Rajamani, Simón Oxenford, Konstantin Butenko, Clemens Neudorfer, Pablo Reinhardt, Patricia Zvarova, Mircea Polosan, Harith Akram, Matteo Vissani, Chencheng Zhang, Bomin Sun, Pavel Navratil, Martin M. Reich, Jens Volkmann, Fang-Cheng Yeh, Juan Carlos Baldermann, Till A. Dembek, Veerle Visser-Vandewalle, Eduardo Joaquim Lopes Alho, Paulo Roberto Franceschini, Pranav Nanda, Carsten Finke, Andrea A. Kühn, Darin D. Dougherty, R. Mark Richardson, Hagai Bergman, Mahlon R. DeLong, Alberto Mazzoni, Luigi M. Romito, Himanshu Tyagi, Ludvic Zrinzo, Eileen M. Joyce, Stephan Chabardes, Philip A. Starr, Ningfei Li, Andreas Horn

## Abstract

Frontal circuits play a critical role in motor, cognitive, and affective processing – and their dysfunction may result in a variety of brain disorders. However, exactly which frontal domains mediate which (dys)function remains largely elusive. Here, we study 534 deep brain stimulation electrodes implanted to treat four different brain disorders. By analyzing which connections were modulated for optimal therapeutic response across these disorders, we segregate the frontal cortex into circuits that became dysfunctional in each of them. Dysfunctional circuits were topographically arranged from occipital to rostral, ranging from interconnections with sensorimotor cortices in dystonia, with the primary motor cortex in Tourette’s syndrome, the supplementary motor area in Parkinson’s disease, to ventromedial prefrontal and anterior cingulate cortices in obsessive-compulsive disorder. Our findings highlight the integration of deep brain stimulation with brain connectomics as a powerful tool to explore couplings between brain structure and functional impairment in the human brain.

---

Studying brain connectomics with deep brain stimulation (DBS) represents a compelling framework for identifying circuits that associate with successful neuromodulation therapy ^1,2^. This potential arises from modeling structural connections activated by variably placed electrodes across patients and relating modulated connections to symptom improvements ^1,3^. As a first-order approximation, effective DBS is seen to act akin to a functional lesion ^4^, achieving downregulation of dysfunctional networks that were involved in neurological or neuropsychiatric symptoms in the first place. By isolating circuits that exhibit the most favorable response to DBS interventions, we hence advance our understanding of the precise brain circuits associated with dysfunction in a particular disorder ^1–3^. As such, this methodology can be used to outline the human ‘dysfunctome’, i.e., the set of connections that are disrupted in given brain disorders – and may be tuned down by successful neuromodulation.

Disrupted interactions between the frontal cortex and basal ganglia lie at the root of numerous brain disorders. These interconnections govern motor, cognitive, and affective functions ^5,6^ and are implemented as fronto-subcortical circuits that cross-communicate ^7,8^, but retain a certain degree of segregation at cortical, striatal, pallidal/nigral, and thalamic levels ^7–11^. While the striatum has often been described as the primary input structure within the basal ganglia, the subthalamic nucleus (STN) has recently been recognized as a second direct input nucleus ^12^. The STN is much smaller than the striatum (∼240 mm^3^) ^13^, but similarly receives efference copies of projections from the entire frontal cortex ^7^. This property renders the STN an ideal gateway for modulating large-scale brain networks through direct electrical stimulation delivered by invasive electrodes.

Indeed, targeting the same nucleus has proven an effective therapy for a heterogeneous spectrum of disorders that includes Parkinson’s disease (PD) ^14^, dystonia (DYT) ^15,16^, obsessive-compulsive disorder (OCD) ^17,18^, and Tourette’s syndrome (TS) ^19,20^. At first glance, it may appear paradoxical that applying electrical stimulation to a subcortical structure of such constrained extent could alleviate symptoms in four disorders which manifest as differently from one another at a phenotypical level. However, this seeming paradox may open a unique opportunity: Since the same compact nucleus is used as a DBS target for different disorders, it acts as a network node that provides therapeutic access to different malfunctioning circuits in each of these conditions. By isolating circuitry whose modulation entails the most substantial treatment benefit, we may be able to disentangle whether one and the same – or rather multiple different – dysfunctional networks are implicated in these multiform phenotypic presentations.

Herein, we apply this concept by integrating 534 DBS electrodes – each implanted for treatment of either DYT, PD, TS, or OCD symptoms – and their corresponding clinical outcomes with detailed structural connectomes of the human brain. We analyze the dataset on both local and global network levels by implementing DBS Sweet Spot Mapping ^21^ and DBS Fiber Filtering ^22^ approaches. The resulting circuits segregate the frontal cortex and its hyperdirect and indirect pathway connections with the STN into distinct dysfunctional territories. We base this work on broad definitions of cardinal symptoms present in each of these disorder (as measured by established rating scales applied in clinical practice).

## Results

### Patient Demographics and Clinical Results

#### Discovery cohorts

Each of the four disorders was represented by two cohorts of bilaterally implanted STN-DBS patients (N = 197, 80 female, 394 DBS electrodes): DYT (N = 70, 38 female), PD (N = 94, 29 female), OCD (N = 19, 10 female), and TS (N = 14, 3 female). Average improvements from DBS ON to baseline were comparable between cohorts and centers. In DYT, the San Francisco cohort presented with an average improvement of 52 ± 42% and the Shanghai cohort of 65 ± 29% on the motor subscale of the Burke-Fahn-Marsden Dystonia Rating Scale (BFMDRS). Patients in the TS cohort from Pisa/Milan benefitted by 62 ± 18% and those from Shanghai by 62 ± 26% on the Yale Global Tic Severity Scale (YGTSS). In PD, Berlin patients improved by 45 ± 23% and the Würzburg cohort by 49 ± 24% on the Unified Parkinson’s Disease Rating Scale – Part III (UPDRS-III). DBS entailed a 45 ± 29% reduction within the Yale-Brown Obsessive-Compulsive Scale (Y-BOCS) for the OCD cohort from London after “STN-DBS only” stimulation, while Grenoble patients improved by 44 ± 32%. A comprehensive summary of demographic and clinical patient characteristics along with DBS and imaging specifications can be obtained from **Table S1**. **Tables S2-5** provide comprehensive patient-specific information. **Fig. 1** recapitulates applied methodological concepts in graphical form. Electrode localization confirmed electrode placement within the subthalamic region in all patients (**Fig. 2**).

**Fig. 1:**
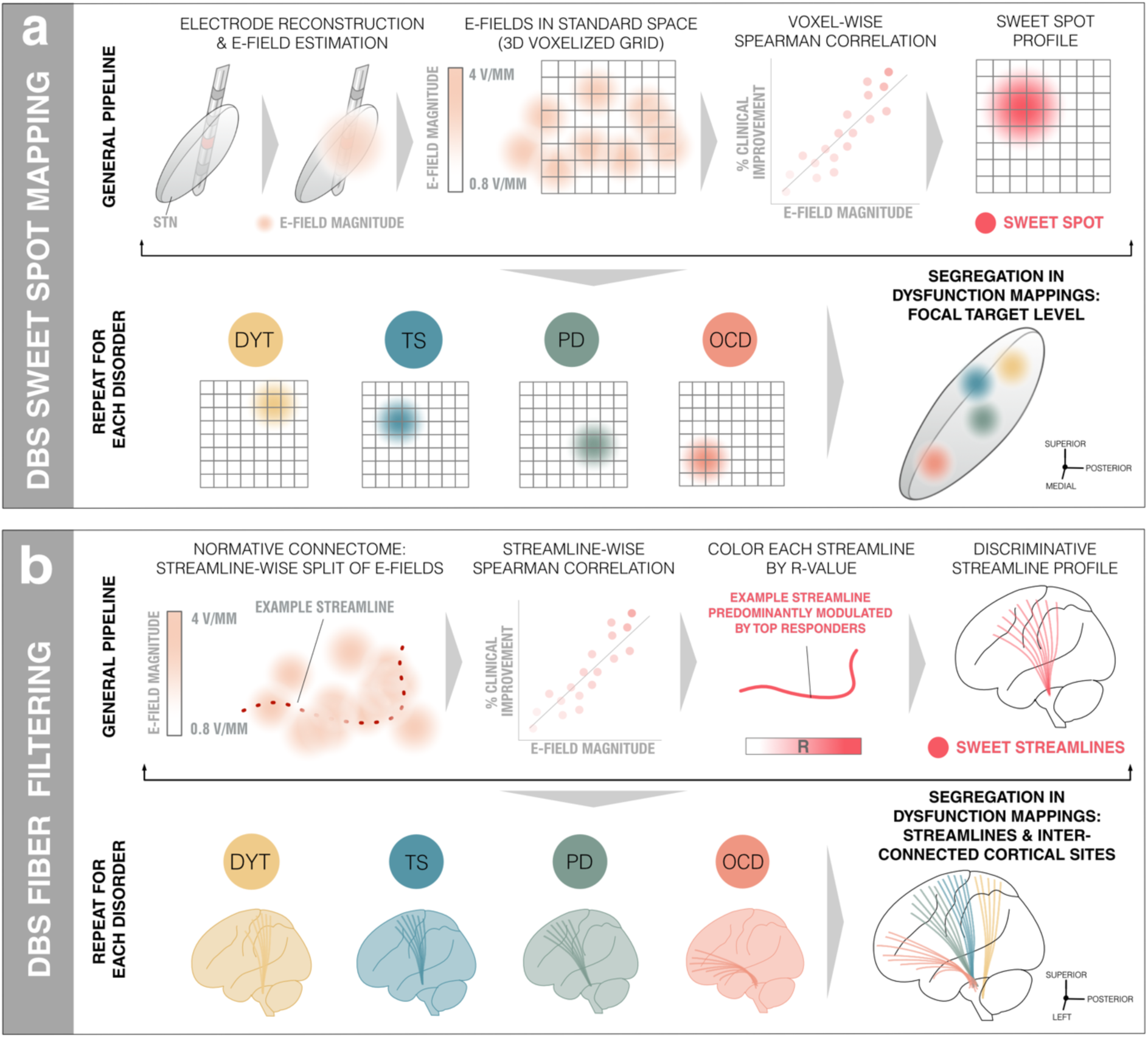
Overview of the two-fold group-level approach to (sub)cortical dysfunction mapping. **(a)** *DBS Sweet Spot Mapping* ^21^. Patient-specific electrode reconstructions were first derived relative to their precise position within the subthalamic nucleus (STN) region and integrated with individual stimulation parameters to estimate electric field magnitudes (E-fields). Subsequently, rank-correlations between E-field magnitudes of the vector and clinical improvements were performed (separately for each disease). Applying this procedure across voxels resulted in a detailed grid of positively (sweet spot) and negatively (sour spot, not shown here) associated stimulation sites. **(a)** *DBS Fiber Filtering* ^22^. Each streamline within a predefined normative connectome was weighted by its ability to discern optimal from poor responders in each respective cohort. To do so, the peak E-field magnitudes among samples drawn along the course of each streamline were rank-correlated with clinical outcomes. Streamlines predominantly modulated by high E-field magnitudes of good responders received high positive weights (sweet streamlines) whereas those associated with high E-field magnitudes of poor responders were attributed high negative weights (sour streamlines, not shown here).

**Fig. 2:**
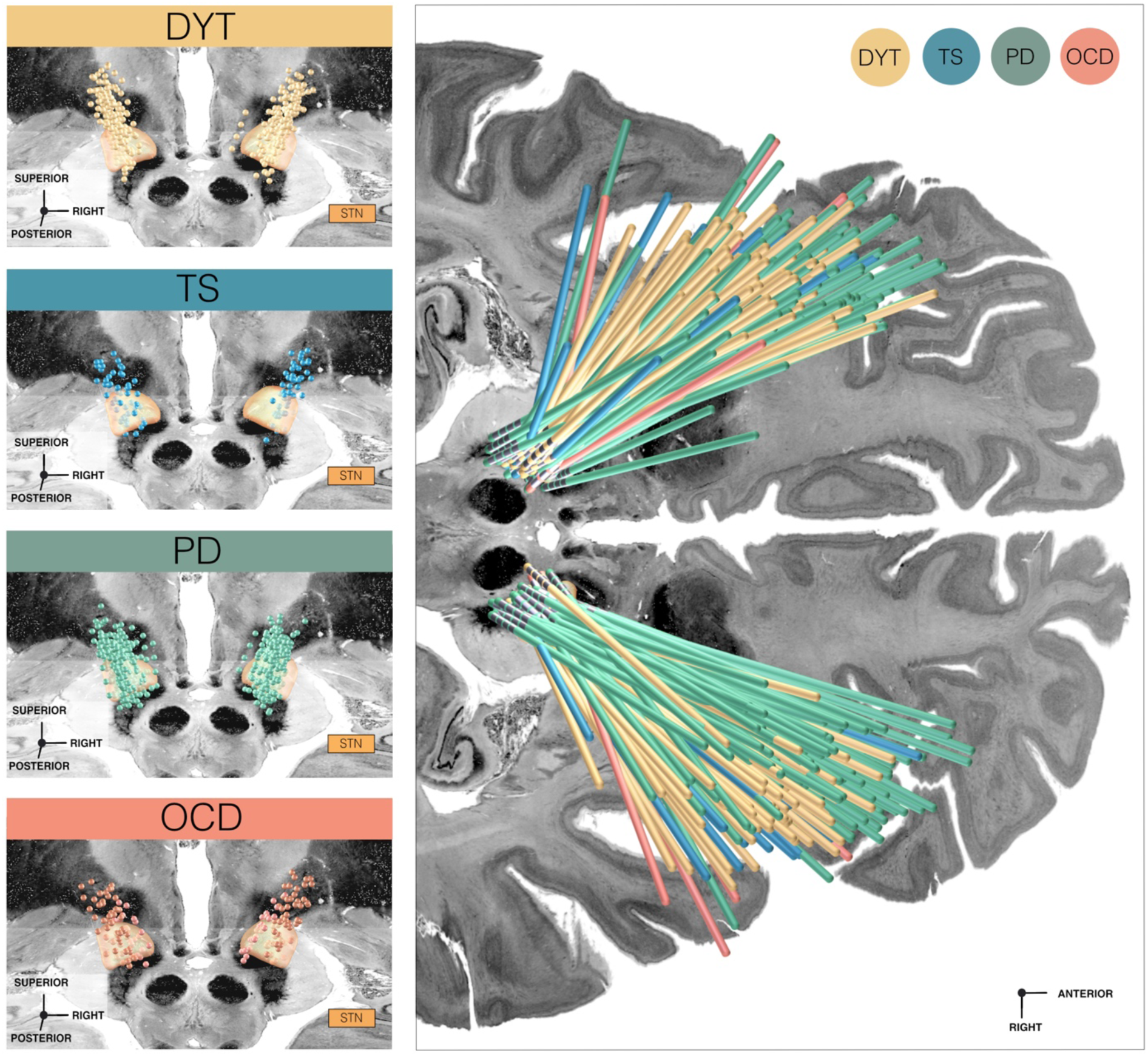
Overview of electrode placements relative to the subthalamic nucleus across discovery cohorts. *Left panels:* Deep brain stimulation (DBS) electrode placement is shown in relation to a posterior view of the subthalamic nucleus (STN) in dystonia (DYT), Parkinson’s disease (PD), Tourette’s syndrome (TS), and obsessive-compulsive disorder (OCD) cohorts, respectively. Electrode contacts are visualized as point clouds. *Right panel:* Visualization of all DBS leads of discovery cohorts investigated in the present study are featured in the axial plane and colored according to indication. STN defined by the DBS Intrinsic Template (DISTAL) atlas ^23^, with an axial plane of the BigBrain template in 100 µm resolution ^24^ displayed as a backdrop (y = -5 mm, z = -10 mm).

#### Validation cohorts

Validation of the PD streamline model was based on an additional STN-DBS cohort (N = 32, 10 female) from Würzburg, characterized by a mean reduction of 47 ± 21% on the UPDRS-III. In OCD, an additional patient cohort (N = 35, 18 female) receiving DBS to the ventral capsule/ventral striatum (VC/VS) region was pooled across Cologne, Boston, and London centers. Critically, electrodes of these novel cohorts were entirely independent from those used to create the streamline models (see methods for the special case of London patients). On average, Cologne patients benefitted from DBS by 31 ± 21%, those from London after “VC/VS-DBS only” by 53 ± 26%, and those from Boston by 40 ± 30% on the Y-BOCS. Additional cohort-averaged specifics are featured in **Table S6**, with patient-wise information in **Tables S7** and **S8**. Finally, two DBS patients (one with PD and OCD each) were prospectively reprogrammed, and one OCD patient was prospectively implanted and programmed as informed on the herein established streamline models (see below for more detailed case vignettes).

### Segregation of Dysfunction Mappings at the Subthalamic Level

#### Model definition

Disease-wise stimulation effects were mapped into anatomical space at the subthalamic level. A caudo-rostral lateral-medial organization emerged for peak voxels associated with beneficial stimulation ranging from DYT to TS, PD, and OCD (**Fig. 3**, center). This result was consistent with functional zones commonly associated with anatomical portions of the nucleus (DYT in the sensorimotor, TS in the motor, PD in the motor-premotor, and OCD in the associative-limbic domain). A detailed overview of the anatomical localization of sweet and sour spots for each disease is provided in **Fig. 3** (upper and lower panels). Peak voxel coordinates are reported in **Table S9**.

**Fig. 3:**
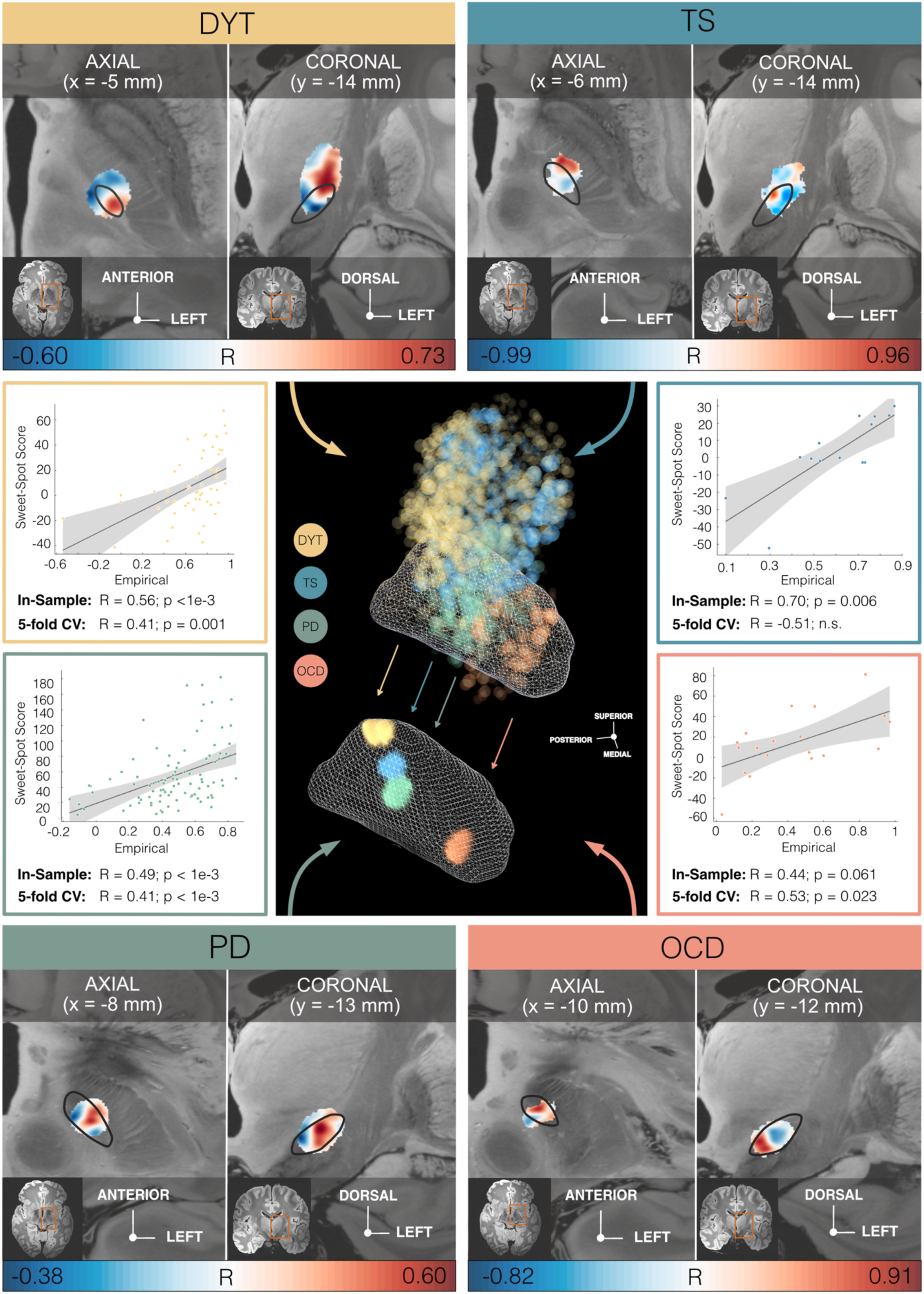
Segregation of dysfunction mappings at the subthalamic level by disease-specific stimulation effects. *Middle panel, center:* The topographical organization of disorder-specific deep brain stimulation (DBS) sweet spots in dystonia (DYT), Tourette’s syndrome (TS), Parkinson’s disease (PD), and obsessive-compulsive disorder (OCD) is shown as a density cloud plot relative to a three-dimensional model of the left subthalamic nucleus (STN) in template space derived from the DBS Intrinsic Template (DISTAL) atlas ^23^. Sphere size and transparency indicate correlation strength between stimulation impact and clinical improvements at a given coordinate, with bigger and less transparent spheres coding for higher correlations. Below, binarized and thresholded sweet spot peaks are projected onto the STN surface. *Upper and lower panels:* Axial and coronal views of disease-specific sweet and sour spots are displayed relative to the left STN (black outlines), superimposed onto an 100 µm ex-vivo brain template ^25^. Voxels are color-coded by degree of correlation (warm colors for positive and cool colors for negative associations) between electric field magnitudes (E-fields) and clinical improvements. *Middle panel, left and right:* Correlation plots show amounts of clinical outcome variance explained by similarity in E-field peaks with disease-wise models of sweet spots across the cohort, with grey shaded areas representative of 95% confidence intervals. *Abbreviations:* CV, cross-validation.

#### Estimation of outcomes based on the model

Spatial correlation of individual E-fields with the optimal pattern was performed to confirm the capability of sweet spot models in explaining clinical outcome variance (**Fig. 3**, middle panel, left and right). This analysis was carried out i) to compare results between diseases, and ii) to compare amounts of variance accounted for by sweet spots vs. streamlines. Critically, these in-sample analyses were circular in nature and should thus not be overinterpreted. To account for this limitation, analyses were repeated in a five-fold CV design (**Fig. 3**, middle panel, left and right) to investigate generalizability of findings, which yielded significant results in all disorders but the TS model (with the lowest N).

### Segregation of Dysfunction Mappings at Streamline and Cortical Levels

#### Model definition

Second, we mapped optimal stimulation effects to fronto-subcortical circuitry. Disease-specific data associated different sets of streamlines with optimal symptom improvements (**Fig. 4a & 5**). Peaks of beneficial DBS networks for DYT primarily interconnected with somatosensory (S1) and primary motor (M1) cortices. Electrode connectivity with M1 and supplementary motor areas (SMA) emerged as most critical for high stimulation benefit in TS, with premotor regions and SMA in PD, and with ventromedial prefrontal, dorsal anterior cingulate, dorsolateral prefrontal and orbitofrontal cortices in OCD. **Fig. S1** displays rotated views of these streamline segregations. Peak voxel coordinates of interconnected cortical sites are summarized in **Table S9**.

**Fig. 4:**
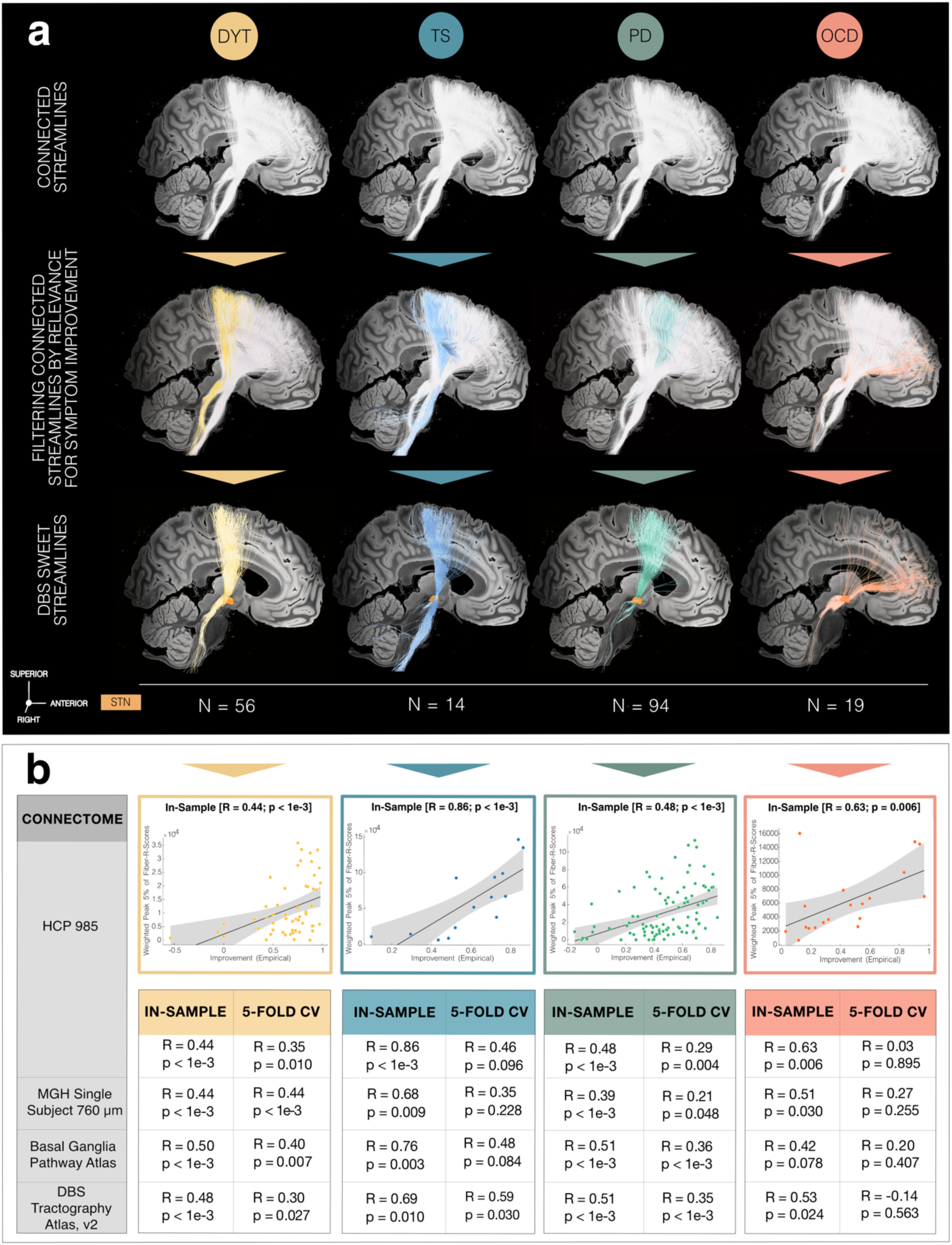
Disease-specific sweet streamline models in each discovery cohort. **(a)** Sweet streamlines in dystonia (DYT) (peak R = 0.36), Parkinson’s disease (PD) (peak R = 0.37), Tourette’s syndrome (TS) (peak R = 0.73) and obsessive-compulsive disorder (OCD) (peak R = 0.49) associated with beneficial stimulation outcomes were filtered from a population-based group connectome ^27^. The first row demonstrates the set of connections (in white) seeding from stimulation volumes across patients in each of the four disorders. Among these plain connections, only those were isolated via Deep Brain Stimulation (DBS) Fiber Filtering (highlighted in disease-specific color; second row) whose modulation correlated with clinical outcomes (third row). Results are shown against a sagittal slice (x = -5 mm) of the 7T MRI ex-vivo 100 µm human brain template ^25^, in conjunction with a three-dimensional model of the right subthalamic nucleus (STN) in template space from the DBS Intrinsic Template (DISTAL) atlas ^23^. **(b)** In-sample correlations and five-fold cross-validations (CV) are reported for models informed on four different normative connectomes. Plots in the top row represent the fitting of a linear model to determine the degree to which overlap of electric field magnitudes with selected HCP 985 Connectome streamlines explains clinical outcome variance across the cohort. Grey shaded areas indicate 95% confidence intervals. *Abbreviation:* MGH, Massachusetts General Hospital.

**Fig. 5:**
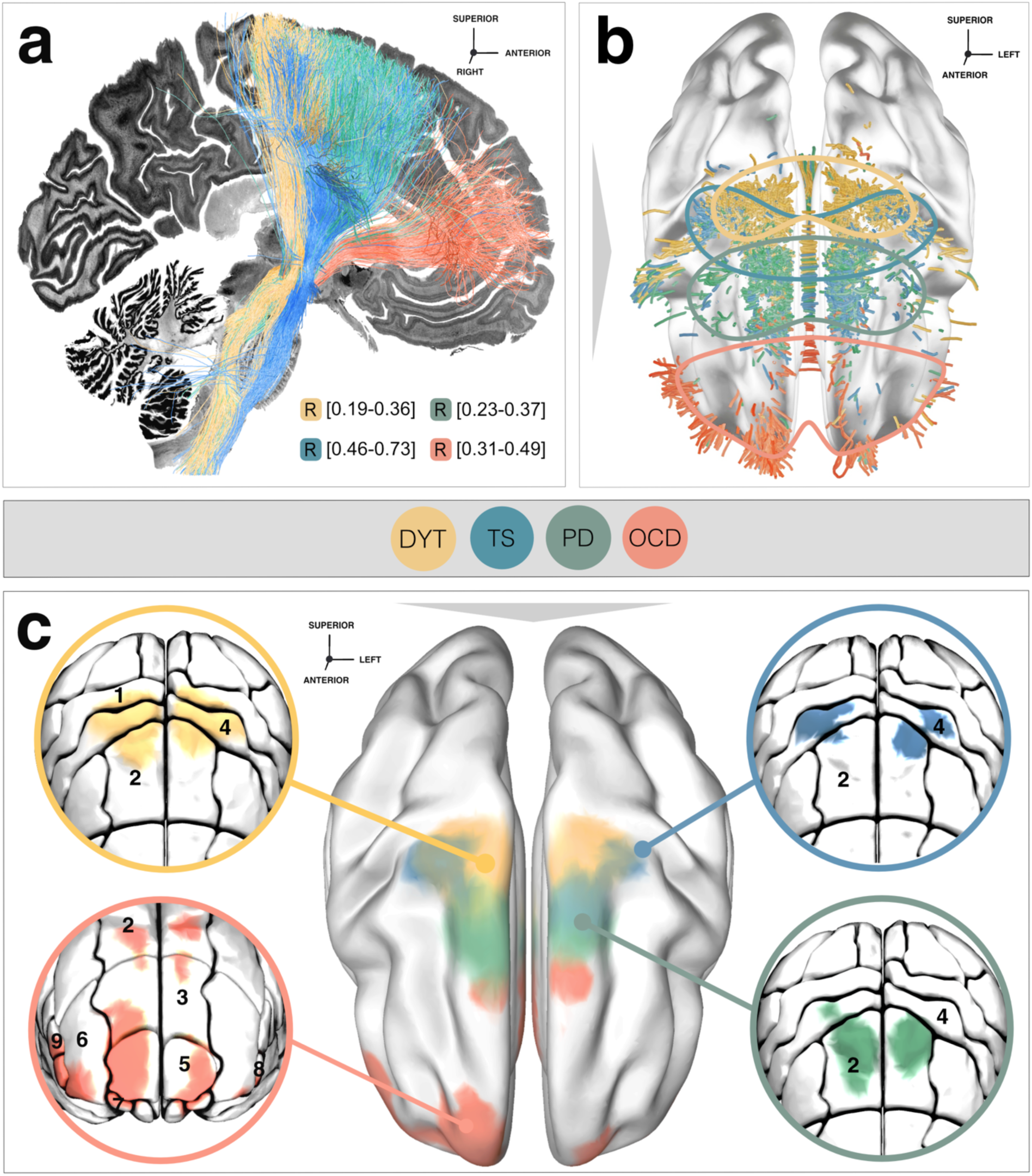
Topography of streamlines and interconnected cortical sites associated with therapeutic stimulation effects in each discovery cohort. **(a)** Segregation into therapeutic networks is achieved by means of deep brain stimulation (DBS) Fiber Filtering in dystonia (DYT), Parkinson’s disease (PD), Tourette’s syndrome (TS) and obsessive-compulsive disorder (OCD). Disease-specific optimal streamlines were isolated from a high-resolution normative group connectome ^27^ through association with clinical effects in each disorder and displayed against a sagittal slice (x = -5 mm) of a brain cytoarchitecture atlas in ICBM 2009b Non-linear Asymmetric (“MNI”) space ^24^. **(b)** Streamlines are shown in conjunction with a transparent brain in template space along with delineations that are color-coded by disease. **(c)** To derive the cortical topography of dysfunction mappings, smoothed density maps of sweet streamlines were projected onto a brain template in MNI space. Zoom-in circles show disease-wise interconnected cortical sites, anatomically characterized based on the Johns Hopkins University (JHU) atlas parcellation ^33^. Legend of relevant regions, with corresponding JHU atlas denominators in brackets: 1 (JHU: 23 & 24), *postcentral gyrus*; 2 (1 & 2), *superior frontal gyrus (posterior segment);* 3 (3 & 4), *superior frontal gyrus (prefrontal cortex);* 4 (25 & 26), *precentral gyrus;* 5 (5 & 6), *superior frontal gyrus (frontal pole);* 6 (9 & 10), *middle frontal gyrus (dorsal prefrontal cortex);* 7 (17 & 18), *lateral fronto-orbital gyrus;* 8 (13 & 14), *inferior frontal gyrus pars orbitalis;* 9 (15 & 16), *inferior frontal gyrus pars triangularis*.

While we consider the visualization of thresholded peaks as most meaningful, thresholding may also mask weaker local optima. **Fig. S2** provides more comprehensive, unthresholded landscapes of sweet and sour streamlines (associated with detrimental outcome). Moreover, we quantified statistical certainty per streamline (defined by the negative log(p) value) to visualize the influence of smaller sample sizes on mappings (**Fig. S3**). As expected, this revealed the lowest certainty for the TS cohort, with the smallest N.

#### Influence of electrode placement

Importantly, DBS Fiber Filtering results yield streamlines weighted by clinical improvements, which should not be confused with mere electrode connectivity. Indeed, mean implantation sites of the standard (second-to-lowest) DBS contact in DYT, PD and TS resided at negligible distance from each other along the y-axis of the STN (p of all independent two-sample t-test comparisons > 0.27) while a significantly different subthalamic aspect was targeted in OCD (all p < 0.01). Hence, we hypothesized the organization of dysfunction mappings to predominantly reflect the stimulation impact on different symptoms – rather than mere differences in electrode placement (at least in all disorders but OCD).

To test this assumption further in a data-driven fashion, we implemented a total of three control analyses. First, from the entirety of plain electrode connections per disease (**Fig. 4a**, first row), we isolated the subset common to all four disorders and compared it to disease-specific streamline models. Both four-sample and pairwise tests of equality of proportions suggested significant differences between proportions of overlap between shared and disease-specific streamlines (**Table S10**).

Second, we mapped sweet spots as three-dimensional Gaussian distributions that were fit to the DBS standard (second-to-lowest) electrode contacts across patients in each disorder. When seeding streamlines from these Gaussians, the resulting streamline profiles expectedly looked much less segregated than the dysfunction mappings achieved by DBS Fiber Filtering. While as anticipated, a slight segregation of the OCD bundle emerged given significantly different placement, streamlines seeding from the Gaussians of other disorders were inseparable. Further, the OCD streamline bundle was by far not as anteriorly located as in the model driven by clinical improvements (**Fig. S4**).

In a final control analysis, we color-coded therapeutic streamlines by a dedicated specificity value. This value was calculated by dividing each streamline’s R-value by the average of the R-values it had been tagged by within the remaining disorder-specific models. The resulting partitioning among disease-specific streamline bundles highly resembled the one achieved by our “conventional” mapping approach (**Fig. S5**), underlining specificity of dysfunction attributions.

#### Estimation of outcomes based on the model

Using optimal streamline profiles to estimate clinical improvements of individual patients based on a (circular) in-sample design resulted in significant correlations for all disorders (**Fig. 4b**). When subjected to five-fold CVs, DYT and PD models were robust across all connectomes but less so for disorders comprising smaller sample sizes (TS/OCD) (**Fig. 4b**). This is not surprising, since five-fold CVs for models calculated on 14 (TS) or 19 (OCD) patients are prone to failure, by design. Robustness of findings from the larger cohorts, however, make us confident about the general validity of methodological choices. Furthermore, the OCD response bundle identified here has been widely reproduced based on OCD cohorts stimulated to the STN and other subcortical targets ^26–32^.

#### Model specificity

Despite discernible segregation in dysfunction mappings, disease-wise streamline models – expectedly – also showed considerable overlap, most visibly among DYT and TS. To quantify the degree of specificity, each profile was hence used to cross-estimate outcomes in all remaining disorders. At large, streamline models explained significant amounts of variance uniquely in the disease for which they had initially been calculated (**Fig. S6**).

#### Influence of connectome

Main analyses were informed on a group connectome ^27^ calculated on diffusion-weighted magnetic resonance imaging (dMRI) based tractography of 985 healthy participants of the Human Connectome Project (HCP) ^34^. While representative of average/population brain connectivity, the choice of this particular connectome may bias results. We thus repeated our DBS Fiber Filtering analyses using five additional normative connectomes. First, we implemented a connectome of unprecedented spatial (760 µm) and angular resolution ^35^ based on a single healthy human brain which optimally lends itself for detailed visualization of dysfunction mappings (**Fig. S7b**).

A key problem of data-driven whole-brain connectomes (as the two above-mentioned), however, is their proneness to false-positive streamlines ^36^ and low accuracy in representing small subcortical tracts ^37^. To account for this, we repeated our analysis based on a pathway atlas manually curated by expert anatomists ^38^ (**Fig. S7c**). While this dataset is likely the most accurate atlas of subcortical streamlines that currently exists, a potential drawback lies in its proneness to false negatives (since not all fibers of the brain were delineated in this resource). Second, as the atlas is not based on empirical dMRI, small details of streamline trajectories in template space may be misaligned. Thus, we replicated analyses using a pathway atlas informed on population based fiber tracking combined with expert defined pathways ^38,39^ which we amended with interconnections between the STN and the entire frontal cortex (DBS Tractography Atlas, v2; **Fig. S7d**).

Finally, the generalizability of dysfunction mappings based on normative connectomes to disease-specific alterations remains uncertain. In view of potential therapeutic implications of mappings, we thus repeated our DBS Fiber Filtering approach in two exemplary disease-matched group connectomes – one informed on diffusion scans of 85 Parkinsonian ^23^ (**Fig. S8a**) and one on those of six OCD patients (**Fig. S8b)**. A comparable rostro-occipital organization emerged across connectomes, with the same order of motor disorders in sensorimotor and premotor cortices toward associative-limbic OCD connections. In-sample correlations and five-fold CVs based on normative and disease-matched connectome models are reported in **Fig. 4b** and **S8c**, respectively.

### Retrospective and prospective validations of streamline models

Given the potential clinical-translational relevance of identified streamlines in guiding treatment for optimized benefit, we carried out a total of five validation experiments in independent datasets. Since not all four diseases could be covered (due to the unavailability of additional retrospective cohorts and prospective enrollments of patients at associated centers), we focused on PD and OCD as two significantly distinct brain circuit disorders. First and second, overlaps between stimulation volumes with the respective streamline model (PD/OCD) were used to estimate outcomes in two additional retrospective cohorts. In both the STN-DBS cohort in PD (R = 0.37, p = 0.043) and the VC/VS-DBS patients in OCD (R = 0.35, p = 0.034), this procedure corroborated a good fit between estimates and empirical outcomes (**Fig. 6a**).

**Fig. 6:**
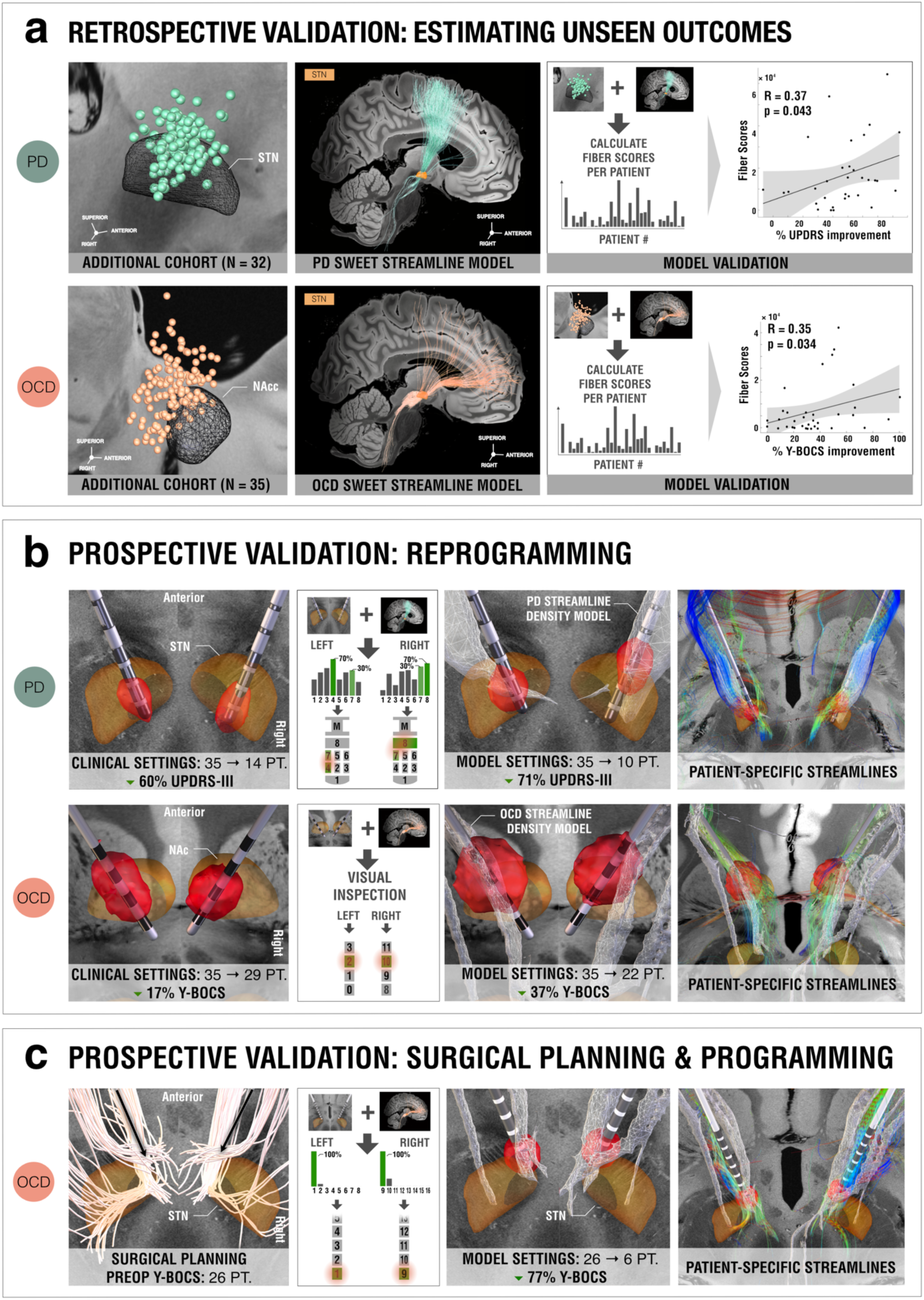
Retrospective and prospective validations of therapeutic streamline targets. To probe the validity of Parkinson’s disease (PD) and obsessive-compulsive disorder (OCD) streamline models, five validation experiments were carried out. **(a)** First and second, empirical outcomes of two additional independent datasets could significantly be estimated based on the degree of overlap of their stimulation volumes with the streamline models. **(b)** Third and fourth, prospective reprogramming was undertaken in two patients. In the PD patient, directional electrodes had been implanted, so the current was divided using a 70/30% rule based on the contacts with the strongest and second-to-strongest streamline overlaps. This led to an improvement of 71% on the Unified Parkinson’s Disease Rating Scale – Part III (UPDRS-III), compared to 60% using clinical settings. In the OCD case, the contact was selected based on visual inspection with the streamline model by the clinical team. This led to a reduction of 37% on the Yale-Brown Obsessive-Compulsive Scale (Y-BOCS), compared to 17% under clinician-selected parameters. **(c)** Fifth, a prospective case underwent streamline-guided deep brain stimulation (DBS) surgery. Electrodes were activated at the contact with the highest streamline overlaps (most ventral contacts on both sides), leading to a rapid Y-BOCS reduction of 77% already one month post-surgically. Depending on the respective target, reconstructed electrodes and stimulation volumes are featured relative to three-dimensional models of the subthalamic nucleus (STN) from the DBS Intrinsic Template (DISTAL) atlas ^23^, or of the nucleus accumbens (Nac) from the California Institute of Technology reinforcement learning (CIT168) atlas ^40^, and against anatomical slices of a 100 µm ex-vivo brain template ^25^.

Third and fourth, we reprogrammed two individual patients at Würzburg (PD) and Boston centers (OCD) with the intention of maximizing engagement of stimulation volumes with the respective streamline model (**Fig. 6b**). The first case comprised a male PD patient (age within range: 66-70 years) with nine years into an akinetic-rigid type PD diagnosis that had been implanted to the STN with directional leads. Three months postoperatively, his score of 35 on the UPDRS-III under DBS OFF improved to 14 points (60% reduction) under clinical DBS ON (with medication OFF in both cases). Under streamline-based parameters, symptoms further reduced to ten points (71% reduction).

The second reprogramming case (treated at the Boston center) was a female patient (age within range: 21-25 years) with severe, treatment-resistant OCD characterized by obsessions about food and water intake along with compulsions involving ingestion events and skin picking. Implantation of a conventional omnidirectional lead targeting the VC/VS region led to an improvement of six points (17%) on the Y-BOCS, from 35 (presurgical baseline) to 29 points postoperatively under clinical parameters. One month after streamline-based reprogramming, this score further reduced to 22 points (37% reduction).

Fifth, we surgically implanted a pair of subthalamic electrodes for treatment of a male patient (age within range: 31-35 years), who had suffered from refractory OCD since the age of 18 years, at the São Paulo center (**Fig. 6c**). After a depressive phase the patient had developed a compulsion of noting every word novel to him and transcribing its meaning from dictionaries, filling numerous notebooks. Later, he began experiencing intrusive death-related thoughts which eventually provoked compulsive religious rituals along with concomitant apathy and depression. Since according to the surgical plan, electrode localization had revealed by far the highest overlaps of the most ventral contacts with the streamline density image, these were activated unipolarly at three milliamperes per side. Only four weeks after surgery, the patient as well as his caregivers reported a dramatic improvement of obsessive-compulsive symptoms that had been notable within one day after switching on the DBS system. The Y-BOCS score had drastically improved to six points, from a pre-surgical baseline of 26 points (77% reduction).

In all three prospective cases, patient-specific tractography confirmed agreement between individual streamlines and normative models (right panels in **Fig. 6b**&**c**). **Table S11** summarizes these prospective patient cases.

### Segregation of Dysfunction Mappings at the Level of Indirect Pathways

While hyperdirect cortico-subthalamic interconnections are best suited to segregate the frontal cortex, the cortico-basal ganglia-thalamocortical system forms loops that include indirect projections from the striatopallidofugal system (and particularly the external pallidum, GPe) to the STN ^41^. These tracts are thought to intermix with the respective hyperdirect pathway interconnections prior to synapsing on STN neurons. Structural representations of the indirect pathway connecting GPe and STN are organized within Edinger’s comb system ^21,42^ and hard if not impossible to reconstruct from diffusion imaging given their orthogonal course to the highly anisotropic internal capsule ^38^.

To interrogate the topography of dysfunction attributions at the level of indirect connections, we thus repeated our DBS Fiber Filtering analysis based on pallido-subthalamic streamlines provided by the Basal Ganglia Pathway Atlas ^38^. Again, segregation was evident between disease-wise interconnected sites, and their organization largely consistent with that of hyperdirect pathway and sweet spot mappings **(Fig. 7)**. While therapeutic indirect connections in DYT projected from sensory/sensorimotor regions of the STN, those in PD connected to territories within the premotor zone of the nucleus. Interconnected sites in TS predominantly resided within associative and those of OCD within limbic subthalamic aspects (insets in **Fig. 7**).

**Fig. 7:**
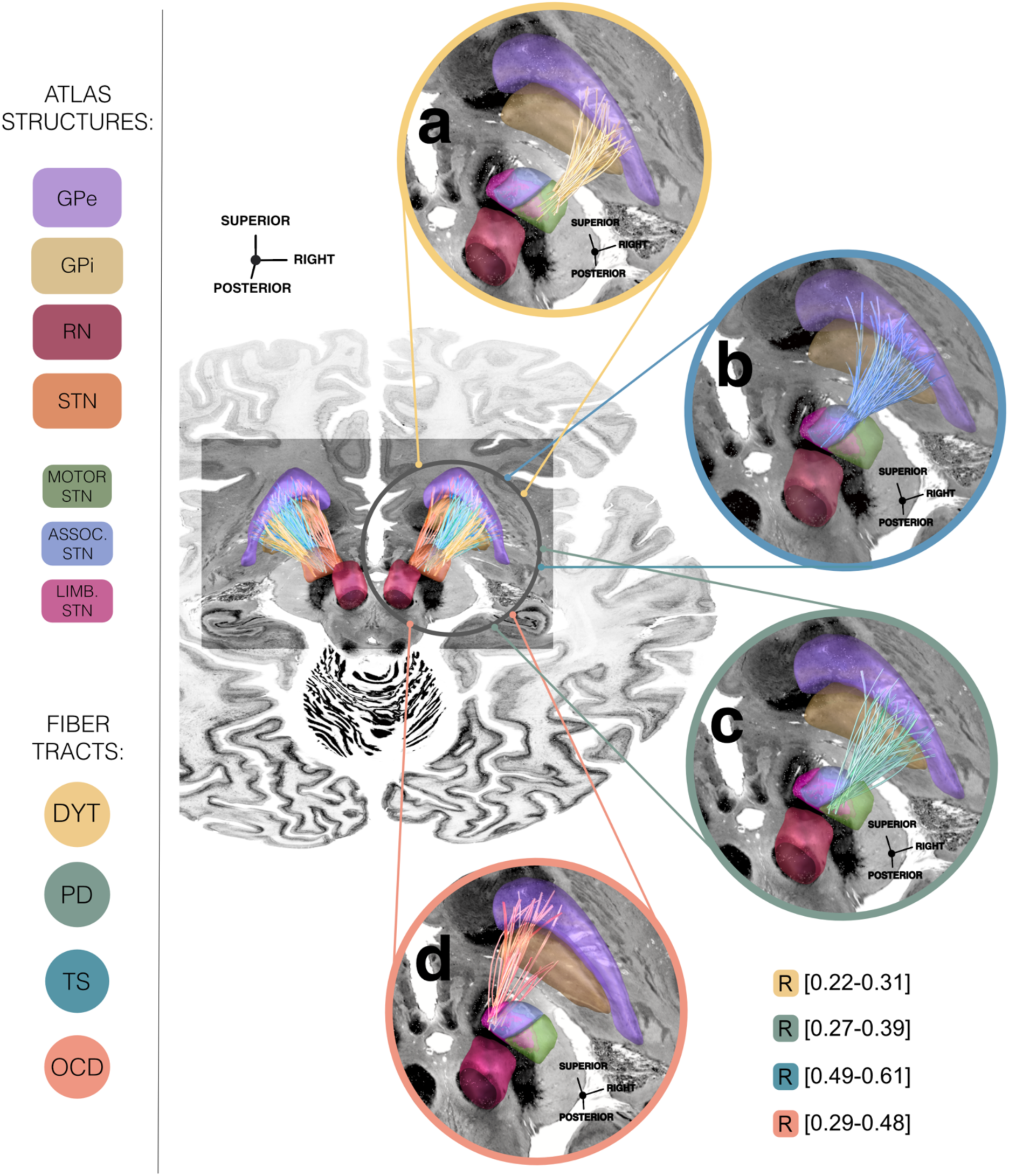
Conserved segregation of dysfunction mappings among indirect pallido-subthalamic connections. Disease-wise sweet streamlines retain a high degree of specificity along their indirect pathway trajectory interconnecting the subthalamic nucleus (STN) with the internal (GPi) and external pallidum (Gpe). Connectivity is modeled based on the Basal Ganglia Pathway Atlas ^38^. Sweet streamlines associated with optimal deep brain stimulation (DBS) outcomes in dystonia (DYT) are interconnected with sensorimotor **(a)**, in Tourette’s syndrome (TS) with associative **(b)**, in Parkinson’s disease (PD) with premotor **(c)**, and in obsessive-compulsive disorder (OCD) with limbic **(d)** STN territories. Streamlines are displayed relative to several anatomical structures from the DBS Intrinsic Template (DISTAL) atlas ^23^ and in conjunction with an axial slice (z = -10 mm) of the BigBrain template ^24^. *Abbreviations:* ass. STN, associative territory of the subthalamic nucleus; limb. STN, limbic territory of the subthalamic nucleus; motor STN, motor territory of the subthalamic nucleus; RN, red nucleus.

## Discussion

Derived from 534 invasive brain stimulation sites spanning eleven patient cohorts and three prospective patient cases treated for either DYT, TS, PD, or OCD across ten international institutions, we draw three key conclusions: First, we showcase the network effects of intracranial brain stimulation as a viable tool for systematically investigating the coupling between circumscribed cortical circuits and selective clinical dysfunctions. As a method, it is capable of mapping what we denote the human ‘dysfunctome’, i.e., the sets of connections that are disrupted and malfunctioning in consequence of given brain disorders. Second, we demonstrate the topographical organization of dysfunction mappings to be mirrored across neuroanatomical levels: i) among prefronto-subthalamic loops and interconnected cortical sites, ii) within pallido-subthalamic connections, and – in miniaturized fashion – iii) within subthalamic subterritories. Dysfunctional networks primarily interconnected the STN with sensorimotor and cerebellar cortices in DYT, while involving M1 and SMA in TS, premotor and supplementary motor areas in PD, and ventromedial prefrontal, anterior cingulate, dorsolateral prefrontal and orbitofrontal cortices in OCD. Third, by their association with previous treatment success, these attributions may hold clinical significance as therapeutic targets in stereotactic neurosurgery and non-invasive neuromodulation ^1–3^. We present first retrospective and prospective evidence of applying these results to inform clinical decision making. Notably, we implement identified circuits to improve treatment benefit in three prospective patient cases.

Methodologically, our study demonstrates the use of subcortical neuromodulation with connectomics as an effective strategy for probing relationships between neuroanatomy and functional impairments. This concept could be perceived as a network-based extension of historical studies that topographically mapped the sites of direct electrical stimulation (often applied cortically during epilepsy surgery) to specific symptoms. Among the most influential authors, Wilder Penfield and team assembled an exhaustive functional map of the cortex based on intraoperative mapping of sensorimotor phenomena ^43^. The present paper applies a paradigm that may be perceived as a derivative of Penfield’s work, combining brain stimulation administered to a specific small (but widely connected) nucleus deep inside the brain with connectomics. Demonstrating the utility of this approach could pave the way to similar work involving other subcortical and cortical neuromodulation sites. Its application at scale and in increasingly fine-grained manner (e.g., by investigating specific symptoms) may lead to more comprehensive definitions of the human ‘dysfunctome’.

Conceptually, our results identify roles of cortico-basal ganglia pathways in different brain dysfunctions using an invasive method ^2^. Attributions were specific to the predominant functional impairment in each disorder and did not merely reflect brain connectivity of differentially placed electrodes – but their relevance for successful symptom treatment. They further were evident irrespective of which healthy or disease-matched connectomes they had been calculated from. Before the teams at Ann Arbor and Johns Hopkins presented their work on what is now referred to as the Albin-DeLong model ^10,44^, the basal ganglia were conceptualized as a funnel that integrates information from different cortical strands to the motor cortex which then initiates action. In essence, the basal ganglia had primarily been categorized as motor structures. Work by Alexander et al. ^9^ challenged this traditional understanding, proposing the idea of parallel circuits involving motor, cognitive and limbic processing. While there is strong cross-communication at least on cortico-cortical, cortico-striatal, striato-nigral and thalamo-cortical levels, loops associated with different functions retain a degree of segregation throughout their cortico-basal ganglia-thalamo-cortical course ^9,45,46^.

The concept of the ’hyperdirect pathway’ builds on this framework, proposing that certain cortical neurons send direct projections to the STN, which bypass the striatum to create a direct link between the cortex and the basal ganglia ^7,47,48^. The functional grouping of subthalamic terminals within these hyperdirect projections can be best understood based on their cortical origins: In this vein, a dorsolateral motor aspect comprising connections to M1 and SMA is defined, along with a ventromedial cognitive territory with origin in superior, middle and inferior prefrontal cortices, and a limbic anteromedial tip connected with orbitofrontal, anterior cingulate and ventromedial prefrontal cortices, hippocampus and amygdala ^6,7,23^. The present results substantiate the general pattern of this distribution based on invasive stimulation sites in four different brain disorders ^2^.

Despite confirming a certain amount of segregation between loops, our findings are also compatible with the concept of cross-communication/integration and so-called open-loop architectures ^7,8,49^. Indeed, therapeutic targets identified here showed considerable overlaps (most notably between DYT and TS; also see **Fig. S2**). In line with the concept of ‘processing gradients’ (rather than entirely segregated loops), our analysis demonstrates preferential mappings between anatomy and stimulation effects. This notion fits with evidence on partial convergence between terminals from different cortical projection sites and interaction between functional subthalamic subterritories ^7,50^.

Clinically, the identified circuits directly represent therapeutic targets that could inform stereotactic targeting in neurosurgery, and potentially noninvasive neuromodulation at the cortical level ^1^. This is underscored by the successful retrospective and prospective validations of the OCD and PD streamline targets in the present study, which provide first evidence for clinical applications of the present findings. We must emphasize, however, that the degree of certainty varies between the studied disorders given the sizes of available samples – especially in TS as a relatively novel application for subthalamic DBS with only few implantations performed world-wide.

Sensorimotor and cerebellar loops have been linked to symptom improvements in DYT by investigations that used comparable sources of information (such as lesions or brain stimulation) ^21,51,52^. The sensorimotor cortex along with its basal ganglia interconnections ^53^, but also the cerebellum and cerebello-thalamic pathway ^54,55^, have been related to dystonic pathophysiology, with noninvasive sensorimotor-cortical ^56^ as well as cerebellar ^57^ stimulation yielding clinical benefit. Under DBS, aspects of motor control such as motor sequence learning, voluntary movement coordination, sensorimotor adaptation, and the control of specific body parts – which depend on the accurate functioning of cerebellar or sensorimotor loops (or an integration of both) ^58^ – may improve.

In PD, fronto-subthalamic loops connecting the SMA to the STN have been deemed critical in both historical ^59^ and recent work ^60–62^. Structural connectivity between subthalamic electrodes and SMA as well as pre-motor areas correlated with motor improvements in PD – and overlap of stimulation volumes with this set of streamlines was associated with outcomes in independent patients ^60^. This is in line with PD motor benefit observed under cortical SMA stimulation ^63^. Functionally, these therapeutic effects may relate to involvement of the SMA in movement selection, preparation and initiation ^64^.

The streamline bundle identified in the OCD cohort emerged as an effective ‘OCD response streamline target’ beyond stimulation of the STN region ^27–30,32^. Conceptualized as an associative-limbic hyperdirect pathway with passage through the internal capsule on its trajectory towards the STN and other mesencephalic nuclei, this bundle is connected to diverse prefronto-cortical regions such as the anterior cingulate and dorsolateral prefrontal cortex ^6,7^ – as confirmed via functional mappings ^28,65^. Concurrently, these cortical sites have been FDA-approved as transcranial neuromodulation targets for OCD ^66^. Functionally, recalibrating this streamline bundle may resolve repetitive thoughts and behavior by interrupting a pathological control signal emitted through hyperactive prefrontal regions ^28,67^.

Network correlates to restore functionality in TS are less established, especially not via the more recent application of subthalamic DBS ^19,20^. Here, the most highly weighted hyperdirect streamlines showed connectivity to M1 and SMA. These regions align with tic-related alterations ^68,69^, have been associated with tic reduction under thalamic ^70,71^ or pallidal DBS ^72,73^, and probed as noninvasive neuromodulation targets for TS ^74^. The SMA, in interaction with M1, may be involved in tic preparation, relaying signals to areas underpinning action monitoring or tic execution ^75^. Effective treatment may facilitate voluntary tic suppression ^64^, or reduce the premonitory urge to tic ^76^.

There are several limitations to our study. First, analyses mainly relied on retrospective data which may bias the interpretation of clinical outcomes. Aggregation of a large multi-center sample (N = 534 electrodes) further inevitably introduced different sources of variability. Still, most of our models extrapolated across differences in targeting strategies between surgeons and centers, imaging modalities and protocols, electrode models, stimulation paradigms, or clinical assessment strategies. Further, we validated results on unseen cohorts and prospectively tested PD and OCD circuits in individual patients.

Second, the underlying physiological effect may not be fully captured by the simplified biophysical model employed here to approximate the amount of tissue activated. For instance, E-field models neglect the impact of stimulation onto glial cells, intracellular processes, or synaptic reorganization ^77^. Also, varying stimulation parameters may entail differential consequences ^78^. For these reasons, refined modeling of the focal stimulation impact might contribute to a further increased validity of results ^37^.

Third, warping lead localizations into template space may have introduced slight mismatches. We sought to counteract this bias as much as possible by using an advanced processing pipeline that included brain shift correction ^79^, multispectral normalization, subcortical refinements, and phantom-validated electrode reconstructions ^80^. We further applied a normalization strategy with comparable performance in STN segmentation as manual delineations by anatomical experts in two independent evaluations ^81,82^. In addition to meticulous visual inspection and refinement of outputs, the subthalamic atlas fit was manually optimized via the WarpDrive toolbox ^83^.

Fourth, with patient-specific dMRI largely unavailable, anatomical delineations were based on normative or disease-matched connectivity. Undoubtedly, the use of connectivity acquired outside of the patient sample in question introduces limitations regarding the anatomical accuracy. Nevertheless, an advantage of normative connectomes lies in their higher resolution and signal-to-noise-ratio than what would be attainable during clinical routine. This unprecedented quality results from the possibility of longer scanning durations and reduced movement artifacts in healthy participants as compared to movement disorder and neuropsychiatric patient populations, as well as the frequent use of advanced acquisition tools. Our study aimed at deriving a “broad lens” description of dysfunctional networks in the average human brain and mappings were largely consistent across four normative connectomes. Additionally, we demonstrated the validity of segregations in the face of disease-specific connectivity alterations. Although here and in previous research ^22,27,60,65^, normative models could significantly account for variance in clinical outcome outside of the discovery sample and showed prospective clinical benefit, the reported streamlines should be further validated in patient-specific data before application in targeting and programming.

Finally, the cohorts of two disorders (OCD and TS) were comparably small and most of the resulting models did not survive cross-validations. While the same methodology was applied to all disorders and hence the approach itself could be validated on the other disorders (PD and DYT), this limitation still stands. We further validated the OCD streamline model using an additional patient cohort and two prospective cases, increasing its credibility. Concurrently, the same bundle has been described ^27,32^ and validated ^26,29–31,84^ in earlier work to be relevant for OCD (for a review see ^28^).

In conclusion, our study demonstrates the potential of invasive brain stimulation as a “flashlight” pointing from the subcortex onto the topography of the human ‘dysfunctome’. The organization of beneficial stimulation effects as a function of symptom domain at the level of fronto-subthalamic circuits and their interconnected cortical sites was mirrored at the subcortical level. This scaling effect on dysfunction attributions across neuroanatomical levels provides a compelling answer to the conundrum of similar clinical effects following stimulation to different access nodes of a shared therapeutic network.

## Methods

### Patient Cohorts, Imaging, and Clinical Assessments

#### Discovery cohort

The present study sought to establish models of optimal focal stimulation sites and streamlines, harnessing a retrospective discovery sample of eight patient cohorts (N = 197) spanning across seven international DBS centers (San Francisco, Shanghai, Berlin, Würzburg, Grenoble, London, and Pisa/Milan). Each of these included patients had been bilaterally implanted with subthalamic DBS for treatment of either DYT (N = 70), PD (N = 94), TS (N = 14), or OCD (N = 19). The full sample consisted of two patient cohorts per disease, with the Shanghai center contributing two cohorts (DYT and TS data). **Table S1** summarizes these cohorts, with more detailed patient-wise demographic and clinical information listed in **Tables S2-5**.

#### Retrospective validation cohorts

To further validate streamline models in two exemplary disorders based on out-of-sample data, two additional patient cohorts were integrated. The first consisted of an additional cohort of PD patients from Würzburg receiving STN-DBS (N = 32), and the second of a cohort of OCD patients, pooled across London, Cologne, and Boston centers, with DBS of the VC/VS region (N = 35). Crucially, these patients contributed entirely independent data points that had not been used to inform the previous streamline model setup. The only exception was formed by the OCD-DBS cohort from London, in which patients had received a set of electrodes each to both targets (STN & VC/VS, with N = 4 electrodes per patient in total), that had been activated independently during the original study ^85^. For this cohort, stimulation settings and clinical scores with “optimized” stimulation of both targets combined or of each target separately were available. For model generation within the discovery cohort (with subthalamic focus), stimulation parameters and corresponding Y-BOCS improvement values collected during the “optimized STN-DBS only” phase were implemented, while corresponding information acquired during the “optimized VC/VS-DBS only” phase was used to inform the retrospective model validation. **Table S6** summarizes these two additional retrospective cohorts. Patient-specific information can be derived from **Tables S7** (PD) and **S8** (OCD).

#### Prospective patient cases

Streamline models for PD and OCD were further prospectively validated by reprogramming DBS settings in a PD and in an OCD patient (from Würzburg and Boston, respectivel), guided by the aim of maximized engagement of their stimulation volumes with the corresponding streamline model. Finally, a single case with OCD (from São Paulo) underwent DBS surgery and programming as informed by the OCD streamline model. **Table S11** summarizes these three patient cases.

Procedures of all clinical trials and studies leading to the collection of this data were carried out according to the declaration of Helsinki from 1975 and all participants signed an informed consent prior to study participation. Post-hoc analyses performed for the purpose of the present manuscript were approved by the institutional review board of Charité – Universitätsmedizin Berlin (master vote EA2/186/18) and treatment of data complied with all relevant ethical regulations.

To inform surgical planning and for exclusion of structural abnormalities, all patients received high-resolution multispectral structural MRI that had been acquired at three Tesla field strength. High imaging quality was ensured through visual inspection by a multidisciplinary team during stereotactic planning, and in case of movement artifacts, preoperative acquisitions were repeated under general anesthesia. Intraoperative microelectrode recordings and macrostimulations as well as either postoperative MRI (N = 73) or computed tomography (CT) of the head (N = 188) (**Tables S1-8 & S11**) were acquired to confirm accurate lead placement.

Specifics on electrode models implanted in each cohort used for the model set-up are summarized in **Table S1**, while the same information for retrospective and prospective model validation cohorts is provided in **Table S6** and **S11**, respectively. Stimulation settings and corresponding clinical improvement scores for all cohorts were selected from times of follow-up to which stimulation effects had sufficiently stabilized (**Tables S1, S6, & S11**).

Times of follow-up available for some patients within the N = 58 cohort of DYT patients from Shanghai were shorter than those of other disease cohorts. In addition, as DYT is a heterogeneous disease of several forms (e.g., generalized, segmental and focal somatotopic expressions), preoperative BFMDRS summary scores in some Shanghai patients were considerably lower than those of patients in the San Francisco cohort. To ascertain stabilized and comparable DBS effects across cohorts, main analyses were thus carried out on the DYT sample including a subcohort of Shanghai patients (N = 44) which sufficed to more conservative inclusion criteria (baseline BFMDRS scores ≥ 5 and follow-up ≥ 6 months). However, we repeated our results on the complete DYT sample (N = 70) including the full Shanghai cohort (N = 58) to demonstrate stability of effects (**Fig. S9**).

Clinical improvement was measured via relative change from preoperative baseline to postoperative follow-up under DBS ON (or from postoperative OFF to ON DBS conditions in the case of PD) within the primary outcome assessment of each disease cohort: BFMDRS in DYT, UPDRS-III in PD, Y-BOCS in OCD, and YGTSS in TS.

### DBS Electrode Localization and Electric Field Modeling

DBS electrodes of all patients were localized based on default settings in an advanced, state-of-the-art processing pipeline as implemented in Lead-DBS software, v3.0 (https://www.lead-dbs.org) ^83^. MATLAB R2022b, v9.13.0.2105380 (The MathWorks Inc., Natick, MA, USA) was used to apply this Lead-DBS based analysis stream. In brief, our approach involved linear coregistrations of postoperative head CT or MRI scans to preoperative T1-weighted images by means of Advanced Normalization Tools (ANTs, http://stnava.github.io/ANTs/) ^86^. Coregistration results were subsequently corrected for potential intraoperative brain shift via an automatized subcortical refinement module (as implemented in Lead-DBS), but also needed to conform to meticulous visual inspection by two expert users (BH and NL). This latter step led to manual refinement in cases where aberrations were detected.

All preoperative acquisitions were used for multispectral spatial normalization into ICBM 2009b Non-linear Asymmetric (“MNI”) template space ^87^ using the Symmetric Normalization (SyN) approach included in ANTs with the “effective: low variance + subcortical refinement” preset in Lead-DBS. This method had outperformed comparable approaches for subcortical normalizations (including STN segmentation) across >10,000 nonlinear warps and different normalization techniques in two independent studies, with precision approaching manual expert segmentation ^81,82^. To maximize registration accuracy further, normalization warp-fields were manually refined using the “WarpDrive” toolbox included in Lead-DBS ^83^, with particular attention to the STN as the anatomical structure in focus (see supplementary methods for further detail). **Fig. S10** shows examples of such optimized normalization warp fields following manual WarpDrive refinements (ANTs + WarpDrive) in direct comparison to unrefined direct results of the automated pipeline (ANTs only). Across analyses and visualizations of results, atlas definitions of the STN were based on the DBS Intrinsic Template (DISTAL) atlas ^23^, a precise subcortical atlas explicitly created for use within Lead-DBS and based on convergent information from multimodal MRI, histology as well as structural connectivity.

Subsequently, electrodes were pre-localized using the phantom-validated Precise and Convenient Electrode Reconstruction for Deep Brain Stimulation (PaCER) algorithm ^80^ in the case of postoperative CT. In the case of postoperative MRI, the trajectory search/contact reconstructions (TRAC/CORE) algorithm ^88^ was applied instead. The resulting pre-localizations were visually inspected and manually refined by two expert users (BH and NL).

Integrating patient-specific active electrode contacts with corresponding stimulation parameters, the electric field (E-field) as the gradient distribution of electrical potential in space was simulated in native patient space via an adaptation of the SimBio/FieldTrip pipeline (https://www.mrt.uni-jena.de/simbio/; http://fieldtriptoolbox.org/) ^89^ as implemented in Lead-DBS^83^. Using a finite element (FEM) approach, a volume conductor model was created on the basis of a four-compartment mesh ^79^, which involves a realistic three-dimensional model of electrodes (metal and insulating electrode aspects) and surrounding anatomy (gray and white matter). Again, gray matter was defined using the DISTAL atlas ^23^. Finally, electrodes and E-fields were transformed into template space based on the manually optimized warp-fields priorly determined during normalization of preoperative MRI acquisitions. These steps allowed for visualization and analysis of electrodes and stimulation fields at the group-level using the Lead-Group toolbox ^61^ as well as DBS Sweet Spot and Fiber Filtering Explorers ^83^.

### Segregation of Dysfunction Mappings at the Subthalamic Level

#### Model definition (**Fig. 1a**)

Our group-level approach intended to delineate and compare the organization of disorder-specific stimulation effects across different neuroanatomical levels, namely i) that of the subthalamic target site (DBS Sweet Spot Mapping), as well as ii) that of prefronto-subthalamic pathways and their interconnected cortical sites (DBS Fiber Filtering).

In the first part of our analysis stream, DBS Sweet Spot Mapping ^21^ (**Fig. 1a**) was performed in each disease cohort separately to identify subthalamic voxels linked to optimal stimulation-related improvements within each respective cardinal dysfunction. For this purpose, information from patient-specific E-fields was integrated with corresponding clinical outcome scores. The E-field denotes the first derivative of the estimated voltage distribution administered to voxels in space, exhibiting greater intensity near active electrode contacts and diminishing rapidly as distance increases. To account for variability in voxels covered across E-fields within each cohort and to circumvent biased results based on too few data points, the region of interest was limited to brain voxels encompassed by at least 50% of E-fields exceeding a magnitude threshold of 200 V/m. While this E-field magnitude corresponds to a commonly assumed estimate of voltage needed to activate axons ^60,90,91^, sweet spot modeling and corresponding quantitative validations were repeated for a range of different thresholds (i.e., 180, 200, and 220 V/m) to demonstrate robustness of results (see **Fig. S11**).

Across voxels encompassed by the group of thresholded E-fields in template space, Spearman’s rank correlations were calculated between E-field magnitudes and relative clinical improvements. This procedure resulted in a map of positive peak voxels associated with beneficial stimulation (sweet spot), as well as negative peak voxels related to detrimental effects (sour spot). Of note, these correlation coefficients should not be interpreted as significant results due to the mass-univariate (voxel-wise) nature of our analysis. Instead, they were validated by probing model performance in estimating clinical outcomes in a five-fold CV design (see below).

#### Estimation of outcomes based on the model

To quantify the capability of each disease-wise sweet spot map in estimating clinical improvements, magnitudes of individual E-fields were multiplied with the model in a voxel-wise fashion and results were averaged across voxels. This procedure resulted in one “Sweet Spot Score” per E-field. Each patient receiving bilateral DBS, the Sweet Spot Scores of the two E-fields were finally averaged, leading to a single Sweet Spot Score per patient.

Our modeling approach followed the logic that E-fields in which peaks spatially overlapped highly with the sweet spot (receiving a high Sweet Spot Score) would be associated with high clinical improvements, while peripheral or no overlaps with the sweet spot (low Sweet Spot Scores) would be linked with low improvements. To probe the tenability of this hypothesis, we performed in-sample correlations between Sweet Spot Scores and empirical clinical outcomes. While in-sample correlation results represent circular outcomes, they allowed to compare results i) across disorders, and ii) between sweet spot and sweet streamline findings.

To investigate the generalizability of results, we further tested whether models were robust when subjected to a five-fold CV design, where the sweet spot model was built on a subset of the discovery cohort in each of the five folds and results were used to estimate clinical outcome of the remaining (held-out) patients. Crucially, since data of remaining patients was not used to inform the model, respectively, the CV strategy was not biased by circularity. Across analyses, p-values were derived based on permuted testing building on 5,000 iterations. For greater detail, the sweet spot modeling and validation approach applied here is described within the supplementary methods section in a narrative fashion.

#### Visualization of subthalamic dysfunction mappings

Disease-wise sweet spots were smoothed by a kernel of two at full width at half maximum using Statistical Parametric Mapping (SPM12) software (https://www.fil.ion.ucl.ac.uk/spm/) to visualize the organizational pattern of subthalamic dysfunction mappings across disorders. Smoothed profiles were projected onto the surface of a three-dimensional model of the STN in ICBM 2009b Non-linear Asymmetric space derived from the DISTAL atlas ^23^ using SurfIce software, v.1.0.20211006 (https://www.nitrc.org/projects/surfice). Three-dimensional density plot renderings of sweet spots were further generated by plotting R-value magnitudes coded by spheres with different sizes and alpha values in space using Lead-DBS. Namely, the size and alpha value (transparency) of spheres was weighted by the correlation of modulating the coordinate with clinical outcomes. Two-dimensional axial and coronal views of sweet and sour spots were additionally displayed separately for each disorder using 3D Slicer software, v5.2.1 (https://www.slicer.org/).

### Segregation of Dysfunction Mappings at Streamline and Cortical Levels

#### Model definition (**Fig. 1b**)

The second part of our analysis stream followed the intention of deriving the topographical organization of dysfunction mappings i) at the level of hyperdirect prefronto-subthalamic streamlines, and ii) that of interconnected sites within the frontal cortex.

To understand the relationship between modulation of specific streamlines via DBS and a given effect on a dysfunctional domain, we thus harnessed a previously validated structural connectivity analysis, termed DBS Fiber Filtering ^32^ (**Fig. 1b**), in an adapted form for implementation in (non-binarized) E-fields ^22^. Structural connectivity was primarily defined by a population-based group connectome derived from dMRI based tractography data of 985 healthy participants acquired within the HCP 1,200 subjects release ^34^. Details on the calculation procedure of this connectome are reported in Li et al. ^27^. While by design, normative connectivity is unable to fully account for patient-specific anatomical variability, it is optimally suited for “broad-lens” insight into the average human brain at particularly high resolution as aimed at in the present investigation. Although small inter-individual differences in the topography of human fronto-subcortical interconnections exist, at least a general agreement can be presumed (see supplementary results and **Fig. S12**).

Again, the streamline modeling procedure was performed in each disease cohort separately. Per disorder-wise cohort, we first isolated the subset of streamlines from the normative connectome that passed in proximity of at least a minimal number of electrodes. These were characterized in the form of streamlines traversing a rather high E-field magnitude (> 0.8 V/mm) close to active contacts in more than 0.5% of E-fields within that cohort. Iterating through this subset of streamlines one at a time, the stimulation impact per E-field on each streamline was estimated by the peak value among E-field magnitudes collected from points along its passage. This resulted in a “streamline by E-field peaks” matrix in which each entry denoted the peak impact of each E-field on each streamline.

Second, the entries of the “streamlines by E-field peaks” matrix were Spearman’s rank correlated with clinical improvements across the disease cohort. Following this procedure, each streamline was tagged by an R-value coding for the association strength of its modulation with beneficial outcome. The resulting streamline profile can be seen as a model of optimal connectivity for maximal clinical improvements, where streamlines with positive weights would be strongly modulated by E-fields of good performers (sweet streamlines) and such with negative weights by E-fields of poor performers (sour streamlines). As these correlation coefficients relied on a mass-univariate approach, streamline profiles were later validated by probing their capability to estimate clinical improvement in data unseen by the model (see below).

#### Estimation of outcomes based on the model

To determine how well disease-wise optimal sweet streamline profiles would perform in estimating clinical improvement in single patients, the peaks of their E-fields were overlapped with the streamline model of optimal electrode connectivity. Specifically, streamlines from the model touched by that E-field were first isolated. Iterating through this subset, the R-value of each streamline was subsequently multiplied by the peak E-field magnitude to account for the strength of its modulation by this E-field. The peak 5% of weighted R-values were then summed up to form a “Fiber R-Score” per E-field. Since each patient within the herein considered cohorts had been implanted to the STN at both hemispheres, the Fiber R-Score was finally averaged across bilateral E-fields to result in one single value per patient.

Following the logic of this procedure, E-field peaks displaying high overlaps with beneficial streamlines would receive high clinical scores while those with low overlap would receive low clinical estimates. A subset of the most relevant sweet streamlines was selected for this validation step using an R-value threshold set at the top 1% of the cumulative distribution function of R-values of all streamlines. This R-value threshold was implemented to discard streamlines with noisy correlations from the model.

Model validation within the discovery cohort followed a similar strategy as implemented in the case of sweet spot mapping. Correlating the weighted peak 5% of Fiber R-Scores with empirical clinical improvements across patients, the ability of streamline models to explain in-sample variance was scrutinized for comparability of results i) across disorders and ii) with sweet spots. Ultimately, all models were subjected to CVs in a five-fold design to investigate the generalizability of their explanatory value in hold-out data. Again, the supplementary methods section comprises are more narrative and detailed description of the modeling and CV procedure. *Visualizations of cortical dysfunction mappings:* To elucidate the topographical organization of interconnected fronto-cortical regions, disease-wise sets of sweet streamlines were first converted to voxelized images (streamline-density maps). The resulting maps were then smoothed using an eight millimeter Gaussian kernel at full width half maximum as implemented in SPM12 (https://www.fil.ion.ucl.ac.uk/spm/) and projected onto the cortical surface of the MNI template using SurfIce software, v.1.0.20211006 (https://www.nitrc.org/projects/surfice). Anatomical correlates of disease-wise cortical sites interconnected with sweet streamlines were then defined based on the Johns Hopkins University (JHU) atlas parcellation ^33^.

#### Quantification of spatial uncertainty

Further, we aimed to quantify and visualize the degree of spatial uncertainty per streamline within disorder-wise dysfunction mappings at the streamline level. For this purpose, the thickness of each streamline was determined by the negative log(p)-value, meaning that thicker streamlines would be illustrative of lower p-values.

#### Influence of electrode placement

Subsequently, we aimed at scrutinizing the relative impact of different model inputs. Besides the choice of a normative connectome, DBS Fiber Filtering results are mainly determined by two major sources of variability across patients: namely, i) by the precise placement of the stimulation volume, and ii) by clinical improvements. In three out of the four disorders of interest in the present study (DYT, PD, and TS), stereotactic targeting aims at the same site within the dorsolateral aspect of the STN, while the OCD target resides more anteromedially. This observation points toward the notion that the partitioning of dysfunction mappings among various disorders could predominantly be driven by the stimulation impact on clinical outcomes. Consequently, it appears that this phenomenon may not solely be reliant on differential electrode placement.

To investigate this hypothesis, we implemented a total of three data-driven control analyses. First, plain streamline connections seeding from bilateral stimulation volumes were isolated for each disorder. These comprised the entirety of structural connections activated by a bilateral E-field, irrespective of – and, unlike DBS Fiber Filtering results, unweighted by – the importance of their modulation for clinical outcome. Among these, only the subset of streamlines shared across disorders was retained and contrasted to disease-specific sweet streamlines. Four-sample and pairwise tests for equality of proportions were performed to compare the degree of overlap between them.

Second, we fit a three-dimensional Gaussian distribution to the standard (second-to-lowest) electrode contacts of all patients with a specific disorder, leading to four blurred volumes within the STN. Streamlines were then seeded from each of these Gaussians as regions of interest. The partitioning among the resulting disease-wise connectivity profiles was consequently visually compared to the streamline segregation model that had been achieved using DBS Fiber Filtering (where streamlines connected to empirical stimulation volumes of patients had been weighted by stimulation-related outcomes within the four different domains of dysfunction).

Third, each connection within each respective sweet streamline model per disorder was color-coded by a specificity value, which was calculated by dividing its R-value by the average of R-values it received across the three remaining disorders. The segregation result of this approach was finally visually compared to that of the connectivity profile that had been established based on “conventional” color-coding as informed by a streamline’s unbiased R-value (resulting from our DBS Fiber Filtering analysis).

#### Model specificity

Besides validation of each model within the respective disorder that it had been calculated on, we were further interested in the degree of specificity of disease-wise models in their ability of explaining outcome variance. To demonstrate specificity, we considered the sweet streamline models for each disorder and overlapped E-fields of patients in all remaining three disorders with the model to predict clinical outcomes in a disorder-by-disorder fashion. Details of this cross-prediction approach were equivalent to those of the CV strategy described above. In the case of specificity of dysfunction mappings, each of the models would show predictive utility uniquely for clinical improvements within the corresponding outcome measure, but not for those of other clinical scales.

#### Influence of choice of connectome

Further, we aimed to scrutinize the influence imposed by a particular normative resource chosen to inform connectivity in our DBS Fiber Filtering analyses. To do so, we repeated modeling and model validation procedures using five additional connectomes based on otherwise equivalent model parameters. The first such resource consisted of a normative whole-brain connectome, derived from a multi-shell diffusion-weighted imaging dataset at 760 µm isotropic diffusion acquired in-vivo in a single healthy participant over a total duration of 18 scanning hours ^35^ (Massachusetts General Hospital [MGH] Single Subject 760 µm Connectome; openly available from https://datadryad.org/stash/dataset/doi:10.5061/dryad.nzs7h44q2). While generalizability of results derived using this connectome to a larger population is naturally limited by its single-patient origin, it lends itself particularly well for detailed anatomical insight and visualization by dint of its unprecedented imaging resolution.

Second, an axonal pathway atlas ^38^ (Basal Ganglia Pathway Atlas; openly available from https://osf.io/mhd4z/) was implemented, which did not rely on tracking of streamlines based on dMRI data and thus circumvents some of the most important drawbacks of dMRI based tractography (such as the possibility of integrating false-positive connections) ^36^. Instead, streamlines included in this tractogram were manually defined by expert anatomists within an advanced augmented reality (holography) framework. Guided by control points, this technique allows for precise localization and reconstruction of basal ganglia anatomy aided by three-dimensional images created from laser beams. While the expert-characterized nature of this resource ensures a highly accurate representation of empirically existing (true-positive) connections, it is limited by a higher degree of false-negative streamlines (as the focus in its creation by the expert anatomists lay in accuracy at the expense of exhaustiveness).

Third, we employed a custom-made pathway atlas (DBS Tractography Atlas, v2; openly available from https://www.lead-dbs.org/helpsupport/knowledge-base/atlasesresources/normative-connectomes/) informed on previously defined pathway atlases, including the DBS Tractography Atlas, v1 ^39^ and the aforementioned Basal Ganglia Pathway Atlas ^38^, which was completed by additional streamlines of particular relevance to this work (see supplementary methods). We created this atlas explicitly for the purpose of the present investigation with a focus on subthalamic connections that were not delineated in other available atlas resources.

Last, we aimed to demonstrate the validity of our findings in the face of disease-specific connectivity alterations in consequence of two exemplary brain circuit disorders. To this end, we repeated our DBS Fiber Filtering approach in a group connectome informed on data by N = 6 OCD patients (1 female, mean age = 45.50 ± 10.52 years). A second disease-matched group connectome ^23^ was further implemented that had previously been calculated based on data by N = 85 Parkinsonian patients (28 female; mean age = 59.48 ± 10.39 years) from the Parkinson’s Progressive Marker Initiative ^92^ (PPMI; www.ppmi-info.org). This connectome (PPMI-85, v1.1; openly available from https://www.lead-dbs.org/helpsupport/knowledge-base/atlasesresources/normative-connectomes/) has been repeatedly used in the DBS context^22,60,93,94^. Information on specifics of the calculation procedure of both these connectomes can be found in the supplementary methods.

Of note, the terms “fibers” or “tracts” should ideally be reserved for anatomical images and not be used to refer to derivatives of tracking algorithms delineating pathways based on water molecule diffusion within the brain. Instead, dMRI-based tractography is an indirect estimate of physical connections – or axons – and cannot inform on their directionality within the brain ^6^. Thus, we speak of “streamlines” to refer to tracking results throughout the manuscript. For reasons of consistency with previous publications, we solely maintain the term “DBS Fiber Filtering” here to denote our streamline modeling approach.

### Retrospective and Prospective Validations of Streamline Models

#### Retrospective model validations

Models informed on data points from the discovery cohort were further externally validated based on fully independent data (see above for the exception of the OCD patients from London). In the two retrospective validation analyses, this strategy was carried out using the exact same approach as the CV analyses performed within the discovery sample, i.e., by calculating the peak magnitude of each E-field at the intersection with each streamline of the respectively corresponding model (PD/OCD) and correlating the resulting aggregated value (“weighted peak 5% of Fiber R-Score”) with empirical clinical improvements.

#### Prospective model validations

In the first reprogramming case of a PD patient from the Würzburg center implanted to the STN, UPDRS-III scores were taken under DBS and dopaminergic medication OFF (following a twelve-hour-long washout period), under active DBS (medication OFF) with clinical DBS settings, as well as under settings that maximized overlap of stimulation volumes with the PD streamline model. All conditions were assessed three months after surgery, and both clinical and streamline-informed DBS settings had been active for at least 24 hours at the time of testing.

In the second case of an OCD patient from the Boston center receiving DBS to the VC/VS region, reprogramming of stimulation settings was guided by the goal of optimized coverage of the OCD streamline model by the patient’s bilateral stimulation volumes. Y-BOCS scores were taken one month after surgery under clinical parameters and under suggested parameters based on consideration of the OCD streamline model, which were both compared to presurgical baseline.

Third, the OCD patient case from the São Paulo center was treated via bilateral lead implantation surgery to the STN as well as stimulation parameter programming, that were both fully informed on the OCD streamline model. Y-BOCS outcome one month following streamline-based surgery and programming was finally contrasted to preoperative OCD symptomatology.

In all three prospective patient cases, a preoperative diffusion scan had been acquired, so that patient-specific tractography could be performed to confirm agreement between individual streamlines and normative connectivity models.

### Segregation of Dysfunction Mappings at the Level of Indirect Pathways

Besides hyperdirect fronto-cortical interconnections, the STN receives indirect projections from the striatopallidofugal system ^41^. In second instance, we thus sought to understand whether and how indirect anatomical connections would be partitioned as a function of stimulation impact on disorder-wise core symptom domains. To this end, we appended an additional DBS Fiber Filtering analysis informed on pallido-subthalamic connections that had been extracted from the Basal Ganglia Pathway Atlas ^38^ while keeping remaining model parameters consistent.

## Data availability

Detailed patient-wise demographic and clinical information is available in the supplementary materials in anonymized form. Patient imaging data cannot be publicly shared as this would compromise patient privacy according to current data protection regulations. It is, however, available from the principal investigators of the collecting sites upon reasonable request within the framework of a data sharing agreement. The sweet streamline atlas and sweet spots of all four disorders are openly available within Lead-DBS software, v3.0 (www.lead-dbs.org). While a processed version of the HCP 985 Connectome ^27^ can be requested from the corresponding authors, source data are freely accessible via the repository of the HCP (https://www.humanconnectome.org/study/hcp-young-adult/document/1200-subjects-data-release). Further, the DBS Tractography Atlas, v2 can be openly downloaded via the Lead-DBS knowledge base (https://www.lead-dbs.org/helpsupport/knowledge-base/atlasesresources/normative-connectomes/). The HCP-1,065 diffusion source data ^95^ used to inform this atlas can be openly accessed via DSI-Studio (https://sites.google.com/a/labsolver.org/brain/diffusion-mri-data/hcp-dmri-data). The following normative resources have been made openly available by the original authors: the MGH 760 µm Connectome ^25^ (https://datadryad.org/stash/dataset/doi:10.5061/dryad.nzs7h44q2) and the Basal Ganglia Pathway Atlas ^38^ (https://osf.io/mhd4z/). The PD-matched PPMI-85 connectome can be openly and publicly derived via the Lead-DBS knowledge base (https://www.lead-dbs.org/helpsupport/knowledge-base/atlasesresources/normative-connectomes/). Source data used for calculation of this connectome can be freely accessed via the homepage of the PPMI (www.ppmi-info.org). The OCD-matched connectome can be shared by the corresponding authors upon reasonable request. Source data of OCD patients employed to calculate this connectome cannot be publicly shared due to patient privacy restrictions.

## Code availability

The entirety of code used in the analyses presented in this work is openly available within the Lead-DBS environment (https://github.com/leaddbs/leaddbs).

## Acknowledgements

Computation has been performed on the High-Performance Compute (HPC) for Research cluster of the Berlin Institute of Health. The authors would moreover like to express their gratitude to Charles G. Jennings, PhD, for very helpful general advice and feedback on the present work. Further, they would like to extend their thanks to the patients and their families for their participation in the clinical trials that allowed for collection of the valuable data analyzed here.

## Authorship contribution statement

B.H.: Conceptualization, Writing - original draft, Data curation, Methodology, Formal analysis, Visualization, Funding acquisition. J.L.O.: Data curation, Resources, Writing - review & editing. I.A.S.: Data curation, Formal analysis, Writing - review & editing. N.R.: Methodology, Software, Resources, Data curation, Writing - review & editing. S.O.: Methodology, Formal analysis, Software, Writing - review & editing. K.B.: Methodology, Software, Formal analysis, Data curation, Writing - review & editing. C.N.: Investigation, Visualization, Writing – review & editing. P.R.: Conceptualization, Methodology, Formal analysis, Visualization, Writing – review & editing. P.Z.: Visualization, Data curation, Formal analysis, Writing – review & editing. M.P., H.A., M.V., C. Z., B.S..: Resources, Data curation, Writing - review & editing. P.N., & M.M.R.: Investigation, Resources, Data curation, Writing – review & editing. J.V.: Resources, Data curation, Writing - review & editing. F.-C.Y.: Methodology, Software, Writing – review & editing. J.C.B., T.A.D., & V.V.-V.: Resources, Data curation, Writing - review & editing. E.J.L.A., & P.R.F.: Investigation, Resources, Data curation, Writing - review & editing. C.F.: Writing - review & editing. A.A.K.: Resources, Data curation, Writing - review & editing. P.N.: Investigation, Writing - review & editing. D.D.D., & R.M.R.: Investigation, Resources, Data curation, Writing - review & editing. H.B. & M.R.D.: Writing - review & editing. A.M., L.M.R., H.T., L.Z., E.M.J., S.C., & P.A.S.: Resources, Data curation, Writing - review & editing. N.L.: Conceptualization, Methodology, Software, Formal analysis, Resources, Data curation, Writing - original draft, Visualization, Supervision, Project administration. A.H.: Conceptualization, Methodology, Software, Formal Analysis, Resources, Data curation, Writing - original draft, Visualization, Supervision, Project administration, Funding acquisition.

## Abbreviations

ANTs: Advanced Normalization Tools
BFMDRS: Burke-Fahn-Marsden Dystonia Rating Scale
CIT168 atlas: California Institute of Technology reinforcement learning atlas
CV: Cross-validation
CT: computed tomography
DBS: deep brain stimulation
DISTAL: DBS intrinsic template atlas
dMRI: diffusion-weighted magnetic resonance imaging
DYT: dystonia
E-field: electric field magnitude
FEM: finite element method
fMRI: functional magnetic resonance imaging
GPe: external pallidum (globus pallidus, external segment)
GPi: internal pallidum (globus pallidus, internal segment)
HCP: Human Connectome Project
JHU atlas: Johns Hopkins University atlas
MGH: Massachusetts General Hospital
MNI space: ICBM 2009b Non-linear Asymmetric template space
M1: primary motor cortex
MRI: magnetic resonance imaging
OCD: obsessive-compulsive disorder
PD: Parkinson’s disease
PaCER algorithm: Precise and Convenient Electrode Reconstruction for Deep Brain Stimulation algorithm
SMA: supplementary motor area
SPM12 software: Statistical Parametric Mapping software
STN: subthalamic nucleus
SyN approach: Symmetric Normalization approach
TRAC/CORE algorithm: trajectory search/contact reconstructions algorithm
TS: Tourette’s syndrome
UPDRS-III: Unified Parkinson’s Disease Rating Scale – Part III
VC/VS: ventral capsule/ventral striatum
Y-BOCS: Yale-Brown Obsessive-Compulsive Scale
YGTSS: Yale Global Tic Severity Scale

