## Supplementary Figures & Methods for "Mapping Dysfunctional Circuits in the Frontal Cortex Using Deep Brain Stimulation"

### Table of Contents


**Supplementary Figures** p. 2

**Fig. S1:** Anatomical relationship of dysfunction mappings at the streamline level across disorders. p. 2

**Fig. S2:** Unthresholded sweet and sour streamline landscapes. p. 3

**Fig. S3:** Visualization of spatial uncertainty in dysfunction mappings at the streamline level. p. 4

**Fig. S4:** Sweet spots and connected streamlines based on a Gaussian fit to standard electrode contacts. p. 4

**Fig. S5:** Visualization of specificity in sweet streamline segregations. p. 5

**Fig. S6:** Model specificity of disease-wise sweet streamlines in explaining clinical outcome variance. p. 6

**Fig. S7**: Influence of choice of connectome on the topography of dysfunction mappings at the streamline level. p. 7

**Fig. S8:** Topographical organization of dysfunction mappings informed on disease-matched connectomes. p. 8

**Fig. S9:** Comparable results based on restricted vs. unrestricted dystonia samples. p. 9

**Fig. S10:** Optimization of normalization warp fields at the subthalamic level. p. 10

**Fig. S11:** Influence of electrical field threshold on sweet spot mapping results. p. 11

**Fig. S12:** Comparison of the topographical organization of cortico-subthalamic interconnections across individuals. p. 13

**Supplementary Methods** p. 14

Manual refinement of normalization displacement fields for electrode reconstruction p. 14

Creation of the DBS Tractography Atlas, v2 p. 14

Creation of Disease-Matched Connectomes p. 16

Narrative Description of Model Set-Up and Validation p. 17

**Supplementary Results** p. 20

Influence of Electrical Field Thresholds on Sweet Spot Mappings p. 20

Interindividual Anatomical Variability in Cortico-Subthalamic Connections p. 20

**References** p. 21

### Supplementary Figures

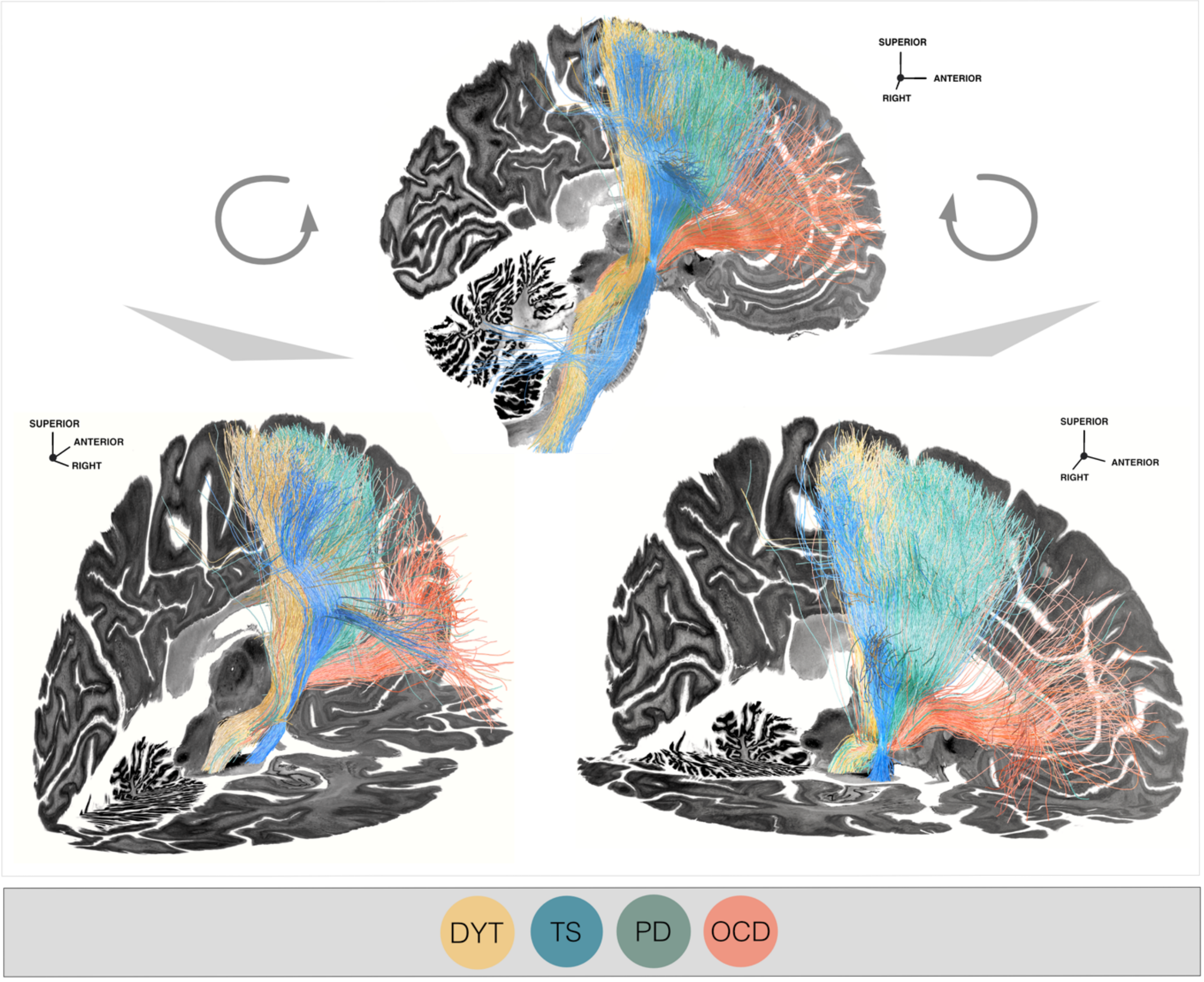


**Fig. S1: Anatomical relationship of dysfunction mappings at the streamline level across disorders.** Posteriorly **(a)** and anteriorly **(b)** tilted views of dysfunction mappings resulting from deep brain stimulation (DBS) Fiber Filtering in dystonia (DYT), Parkinson's disease (PD), Tourette's syndrome (TS), and obsessive-compulsive disorder (OCD). Disease-specific analyses were informed on streamlines extracted from a population-based group connectome [1]. Streamlines connected to stimulation volumes within each respective disorder were weighted by their association with clinical improvement in the disorder-specific clinical improvement measure. Results are displayed in relation to a sagittal (x = 5 mm) and an axial slice (z = -17 mm) of the Big Brain template [2] in ICBM 2009b Non-linear Asymmetric ("MNI") space for orientation.

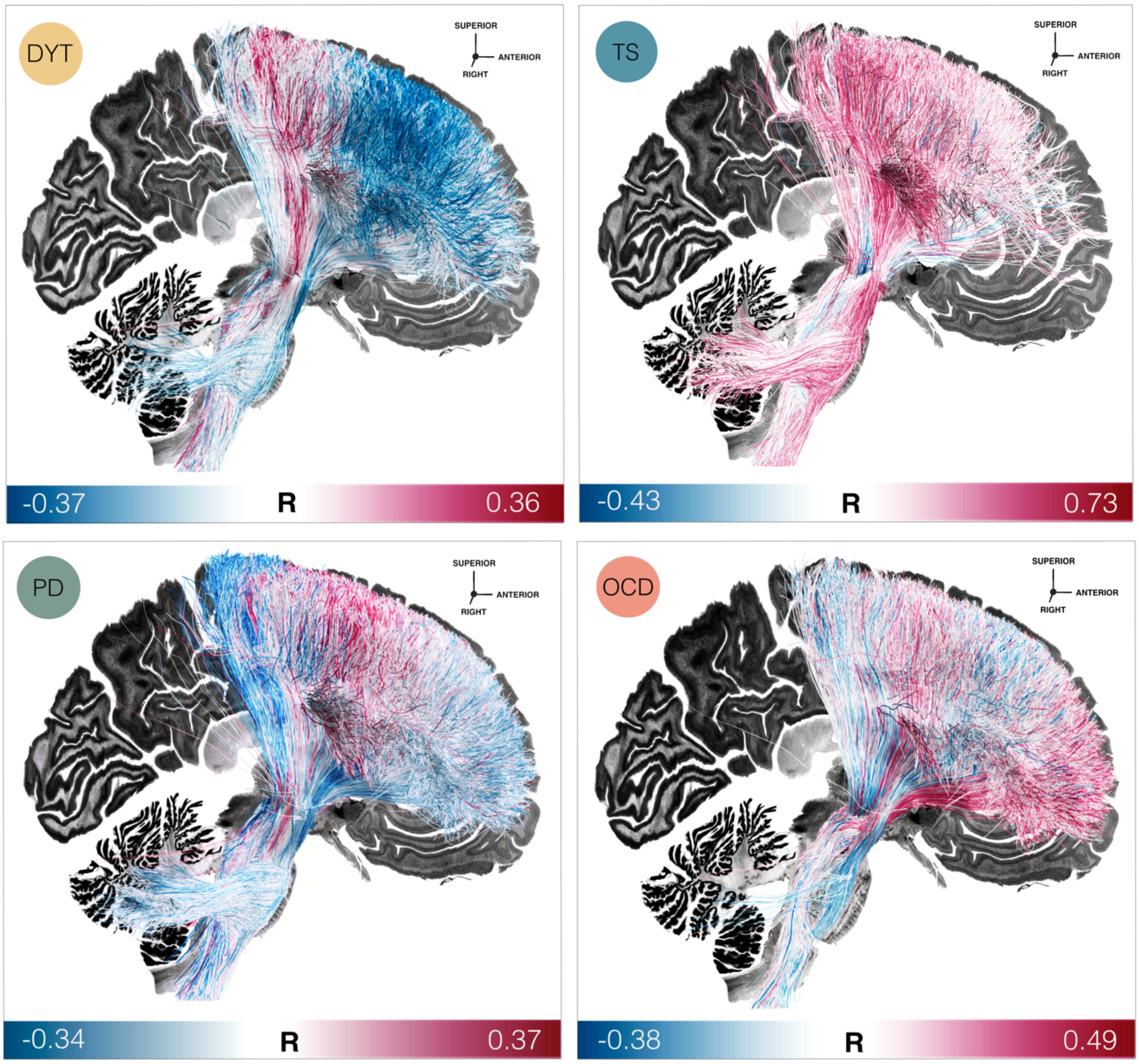


**Fig. S2: Unthresholded sweet and sour streamline landscapes.** Unthresholded models of optimal electrode connectivity for maximized stimulation-related improvement (sweet streamlines, in red) and worsening (sour streamlines, in blue) in disease-specific dysfunction resulting from Deep Brain Stimulation (DBS) Fiber Filtering in dystonia (DYT), Parkinson's disease (PD), Tourette's syndrome (TS), and obsessive-compulsive disorder (OCD). Each streamline comprised within disease-specific models is color-coded by an R-value which denotes the association between the strength of its modulation with clinical outcomes across patients in the respective disorder. Accordingly, streamlines whose modulation is of highest importance for treatment success receive dark red colors, while those whose modulation is most relevant for suboptimal outcomes are colored in dark blue. The whiter the streamline, the less discriminative it was in coding for optimal vs. suboptimal outcomes. Disorder-wise results are overlayed on top of a sagittal slice (x = -5 mm) of the Big Brain template [2].

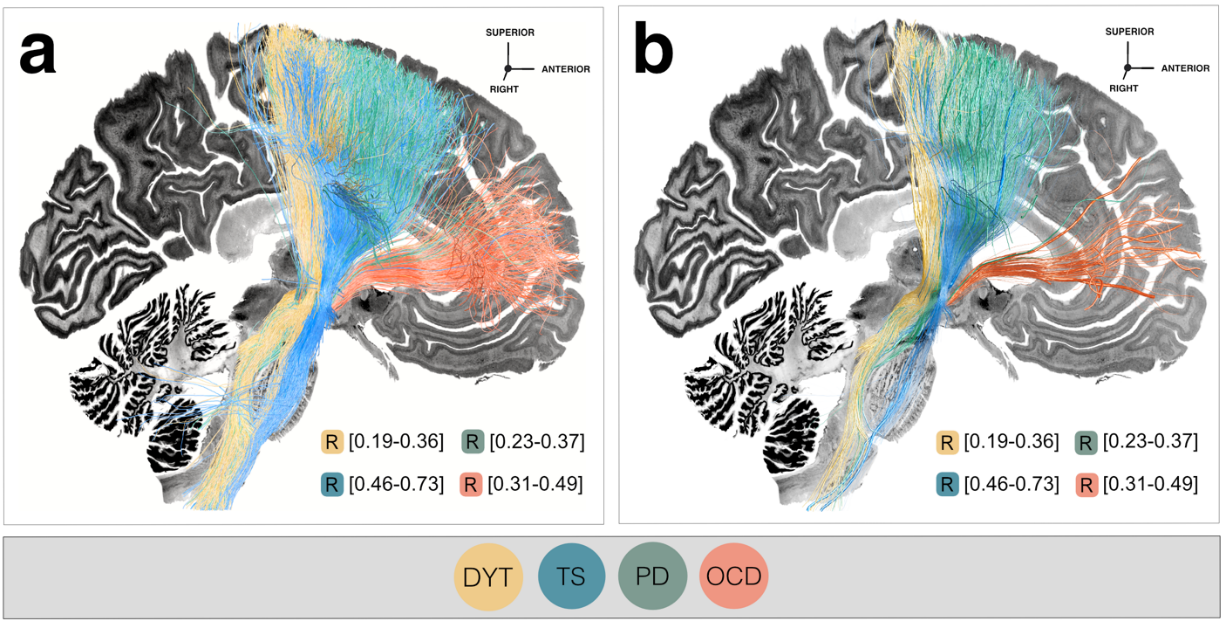


**Fig. S3: Visualization of spatial uncertainty in dysfunction mappings at the streamline level. (a)** Segregation into therapeutic streamline bundles achieved by means of deep brain stimulation (DBS), with streamline thickness attributed in a universal fashion across the connectome (as done in the visualizations illustrating the main analysis of the present manuscript). **(b)** The same DBS Fiber Filtering results are shown, this time with streamline thickness informed based on the negative log(p)-value of the correlation coefficient the respective streamline had rec in the corresponding disorder-wise model. Accordingly, lower p-values are representative of thicker streamlines. Applying this procedure demonstrates a higher degree of uncertainty of TS results (given the low N) but reinforces the OCD results which were also confirmed in an unseen validation cohort (**Fig. 6**). Results are represented against the backdrop of a sagittal slice (x = -5 mm) of the Big Brain template [2]. Abbreviations: DYT, dystonia; OCD, obsessive-compulsive disorder; PD, Parkinson's disease; TS, Tourette's syndrome.

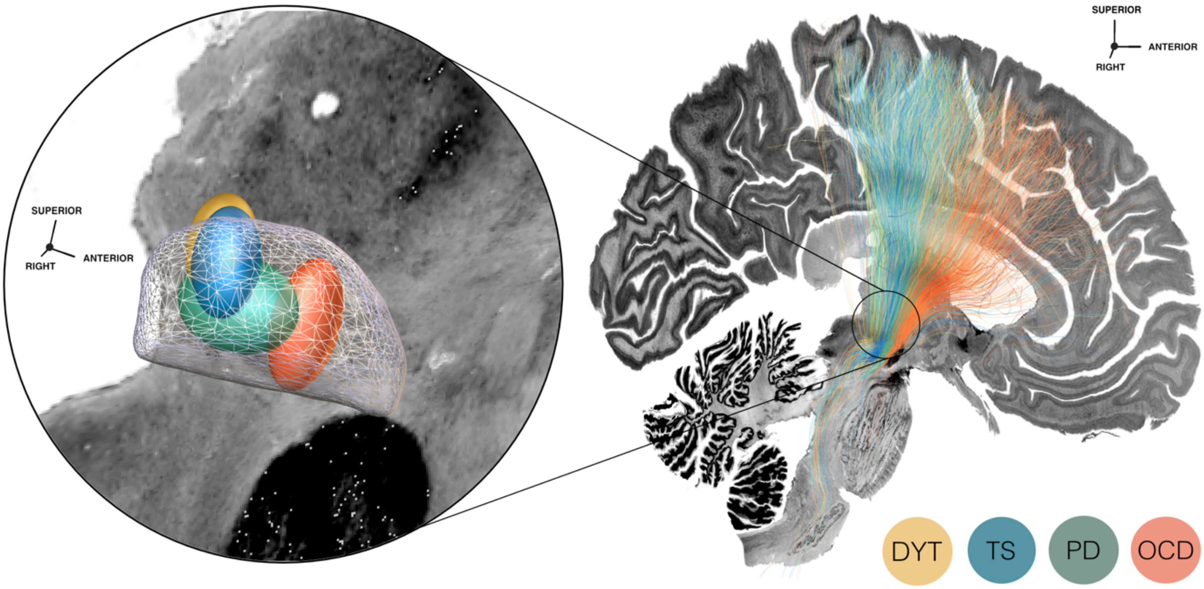


**Fig. S4: Sweet spots and connected streamlines based on a Gaussian fit to standard electrode contacts.** (**a**) Sweet spot maps are fit to the standard (second-to-lowest) electrode contact in form of a Gaussian for each disease cohort. Results are represented relative to a three-dimensional model of the left subthalamic nucleus in template space derived from the DBS Intrinsic Template (DISTAL) atlas [3]. **(b)** Streamlines are seeded from the Gaussian sweet spot maps per disorder and displayed against a sagittal slice (x = -5 mm) of the Big Brain template [2]. Note that the partitioning of disease-wise sweet streamline bundles expectedly vanishes entirely for those disorders known to be treated by implantation to the same posterolateral site of the subthalamic nucleus, encompassing dystonia (DTY), Tourette's syndrome (TS), and Parkinson's disease (PD). Segregation is retained for OCD streamlines which is expected given the different (anteromedial) subthalamic implantation site typically chosen by surgeons to treat this disorder. Nonetheless, the resulting streamlines course much more dorsally compared to the sweet streamline mapping retrieved using Deep Brain Stimulation (DBS) Fiber Filtering [4]. These findings support the notion that segregations in DBS Fiber Filtering-based dysfunction mappings are not merely driven by a-priori differences in electrode placement between disease-wise cohorts, but majorly influenced by clinical outcomes.

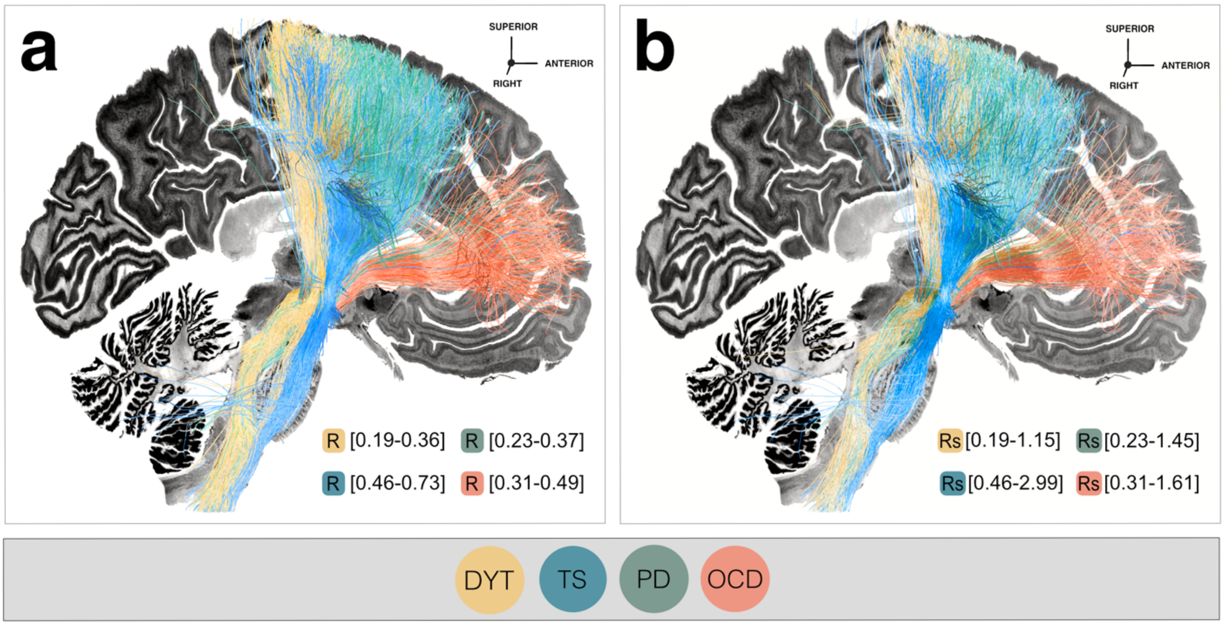


**Fig. S5: Visualization of specificity in sweet streamline segregations. (a)** Mappings of improvements in disease-wise dysfunction following deep brain stimulation at the level of streamlines in dystonia (DYT), Parkinson's disease (PD), Tourette's syndrome (TS), and obsessive-compulsive disorder (OCD). Coloring of streamlines is graded by the magnitude of its R-value per disease, with streamlines whose modulation was associated with higher treatment benefit receiving more intense color values. **(b)** In each disorder, coloring of the same therapeutic streamline models is graded by a dedicated specificity value (denoted as 'Rs'). For each streamline, this specificity value was calculated by dividing its R-value by the average of R-values that streamline had received within data of the remaining three diseases. In direct comparison, the resulting segregations look highly similar, which corroborates the specificity of segregated dysfunction attributions resulting from Deep Brain Stimulation Fiber Filtering [4]. Both views are shown in relation to an axial slice (x = -5 mm) of the Big Brain template [2] in ICBM 2009n Non-linear Asymmetric ("MNI") space.

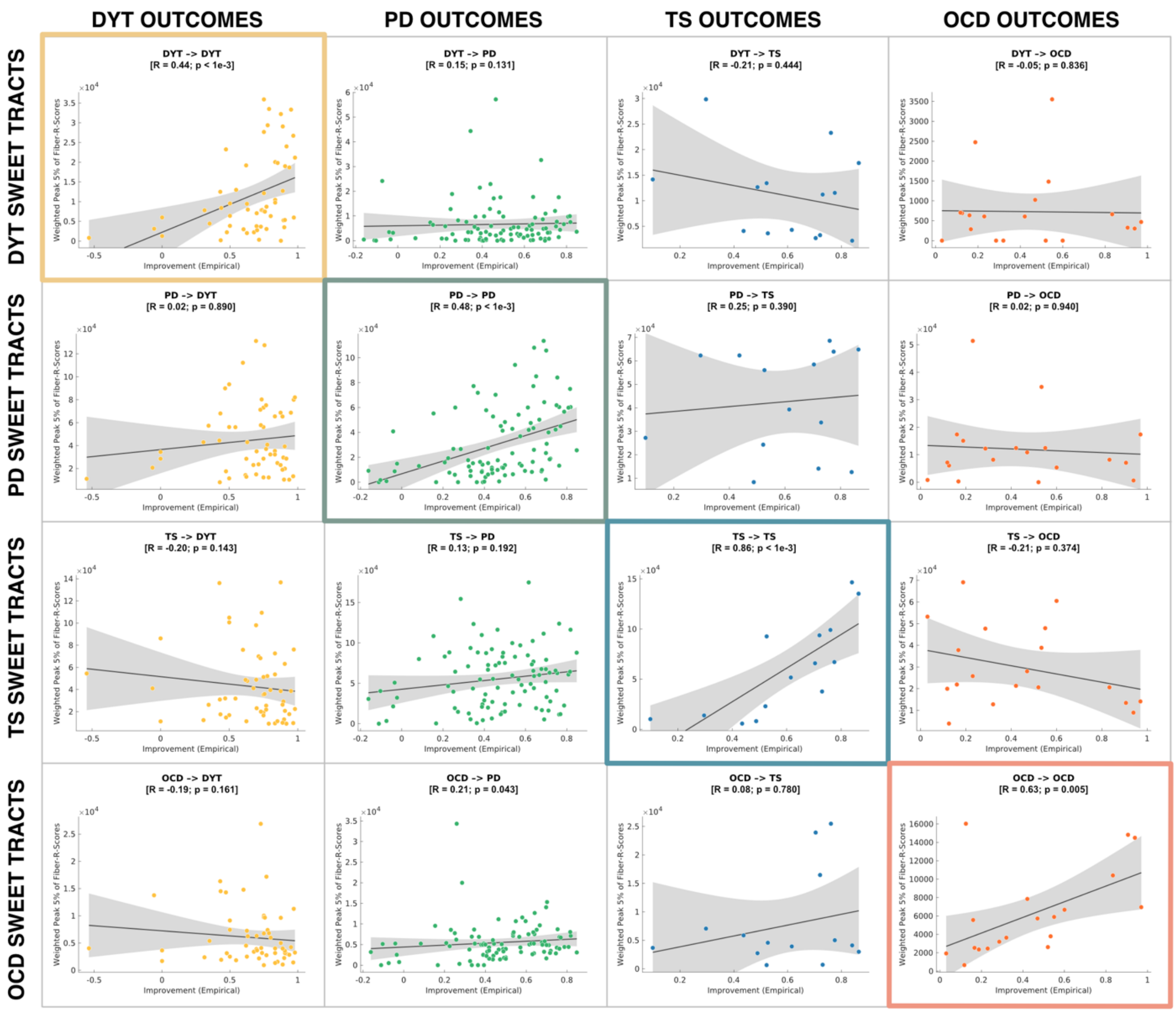


**Fig. S6: Model specificity of disease-wise sweet streamlines in explaining clinical outcome variance.** Based on the degree of overlap of electrical fields with sweet streamline profiles of all remaining three disorders, clinical improvement in each disease cohort is cross-predicted. This is done to confirm model specificity in explaining outcome variance within the respective domain of dysfunction. Streamline models considered here had been informed on a connectome calculated from 985 healthy participants from the Human Connectome Project [5]. P-values of these Spearman's correlations are based on permutation tests of 5,000 randomizations. Grey shaded areas represent 95% confidence intervals. *Abbreviations:* DYT, dystonia; TS, Tourette's syndrome; OCD, obsessive-compulsive disorder; PD, Parkinson's disease.

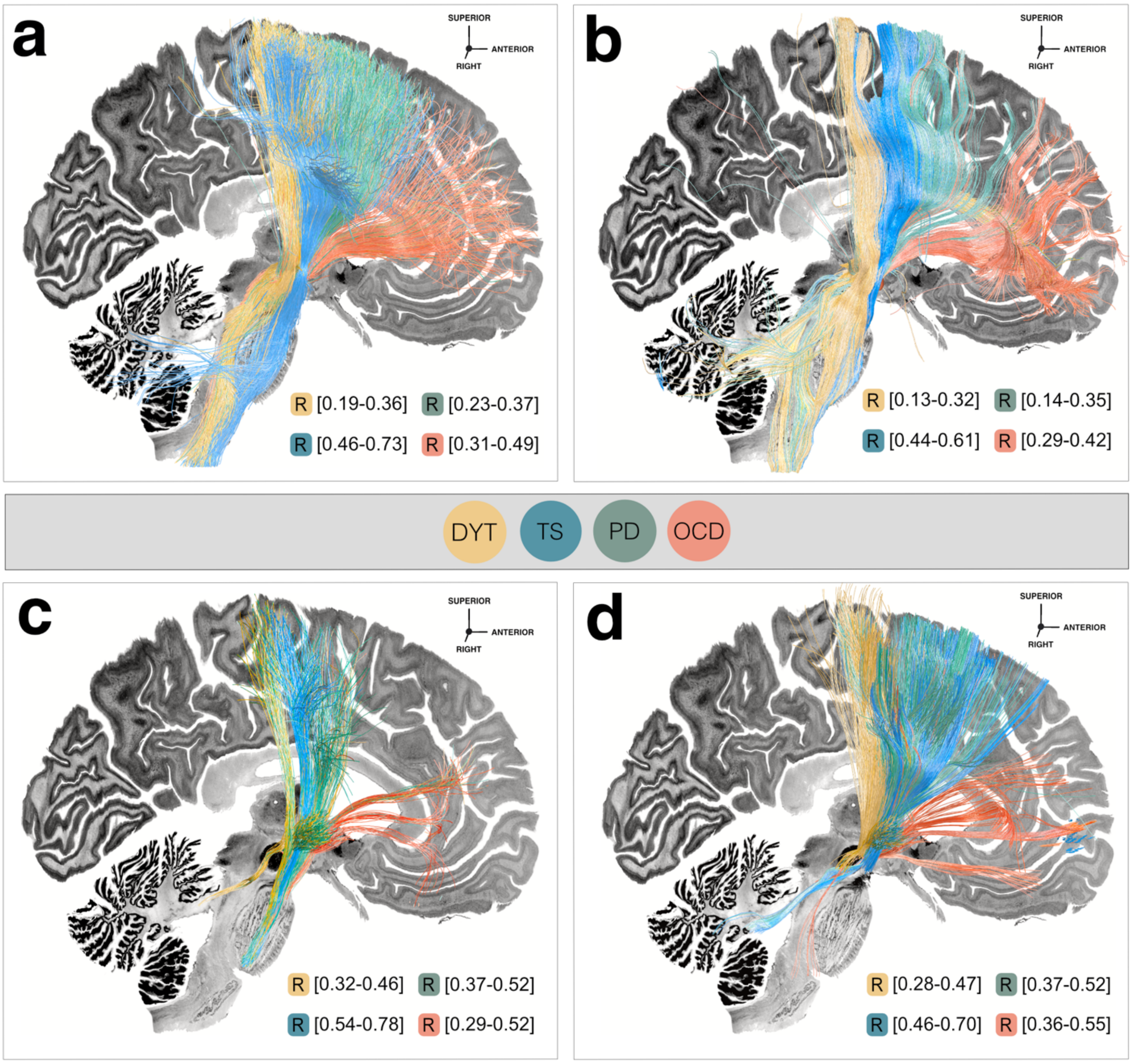


**Fig. S7: Influence of choice of connectome on the topography of dysfunction mappings at the streamline level.** To scrutinize the impact of choosing a specific connectomic resource on the topographical organization of dysfunction attributions, we recalculated disease-wise deep brain stimulation (DBS) Fiber Filtering analyses using four different connectomes. A similar caudo-rostral topography emerged for clinically beneficial sets of streamlines filtered from the Human Connectome Project (HCP) 985 Connectome [1,5] **(a)**, the Massachusetts General Hospital Single Subject 760 µm Connectome [6] **(b)**, the Basal Ganglia Pathway Atlas [7] **(c)**, as well as the DBS Tractography Atlas, v2 (see supplementary methods) **(d)**. Results are shown against a sagittal slice (x = -5 mm) of the Big Brain template [2]. *Abbreviations:* DYT, dystonia; OCD, obsessive-compulsive disorder; PD, Parkinson's disease; TS, Tourette's syndrome.

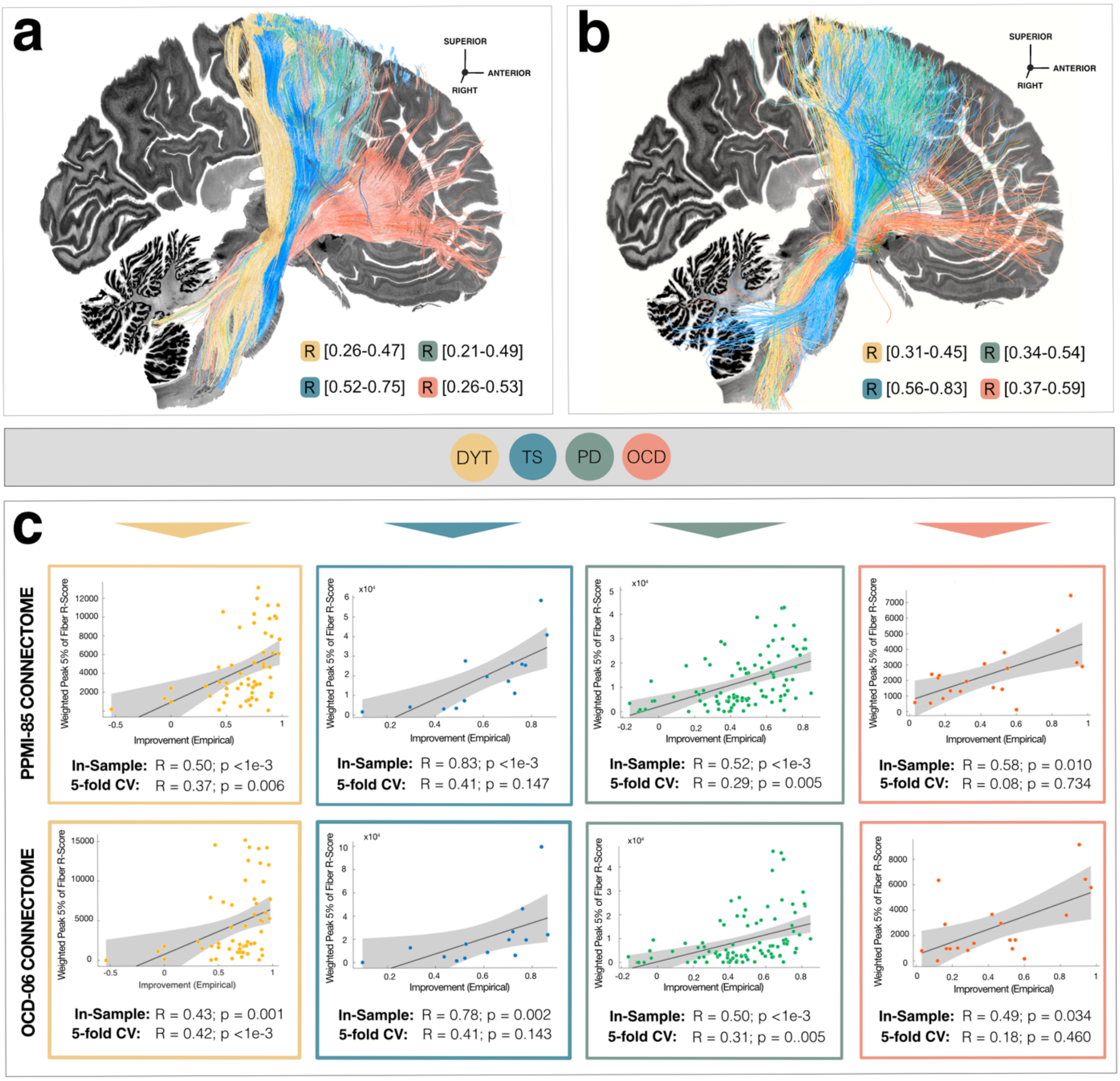


**Fig. S8: Topographical organization of dysfunction mappings informed on disease-matched connectomes.** To investigate the influence of disease-specific connectivity alterations on mappings reported in the main manuscript, the deep brain stimulation (DBS) Fiber Filtering analysis was repeated on **(a)** an obsessive-compulsive disorder (OCD) matched connectome calculated on diffusion-weighted imaging based tractography of N = 6 OCD patients (see supplementary methods). Second, the analysis was informed on **(b)** a previously derived Parkinson's disease (PD) matched connectome [3] based on data by N = 85 PD patients from the Parkinson's Disease Progressive Marker Initiative [8] (PPMI; www.ppmi-info.org). Results are overlayed on top of a sagittal slice (x = -5 mm) of the Big Brain template [2]. **(c)** Finally, in-sample correlations and five-fold cross-validations (CV) were carried out to scrutinize the capability of streamline models to explain outcome variance within the respective clinical scale. Grey shaded areas are indicative of 95% confidence intervals. Abbreviations: DYT, dystonia; TS, Tourette's syndrome.

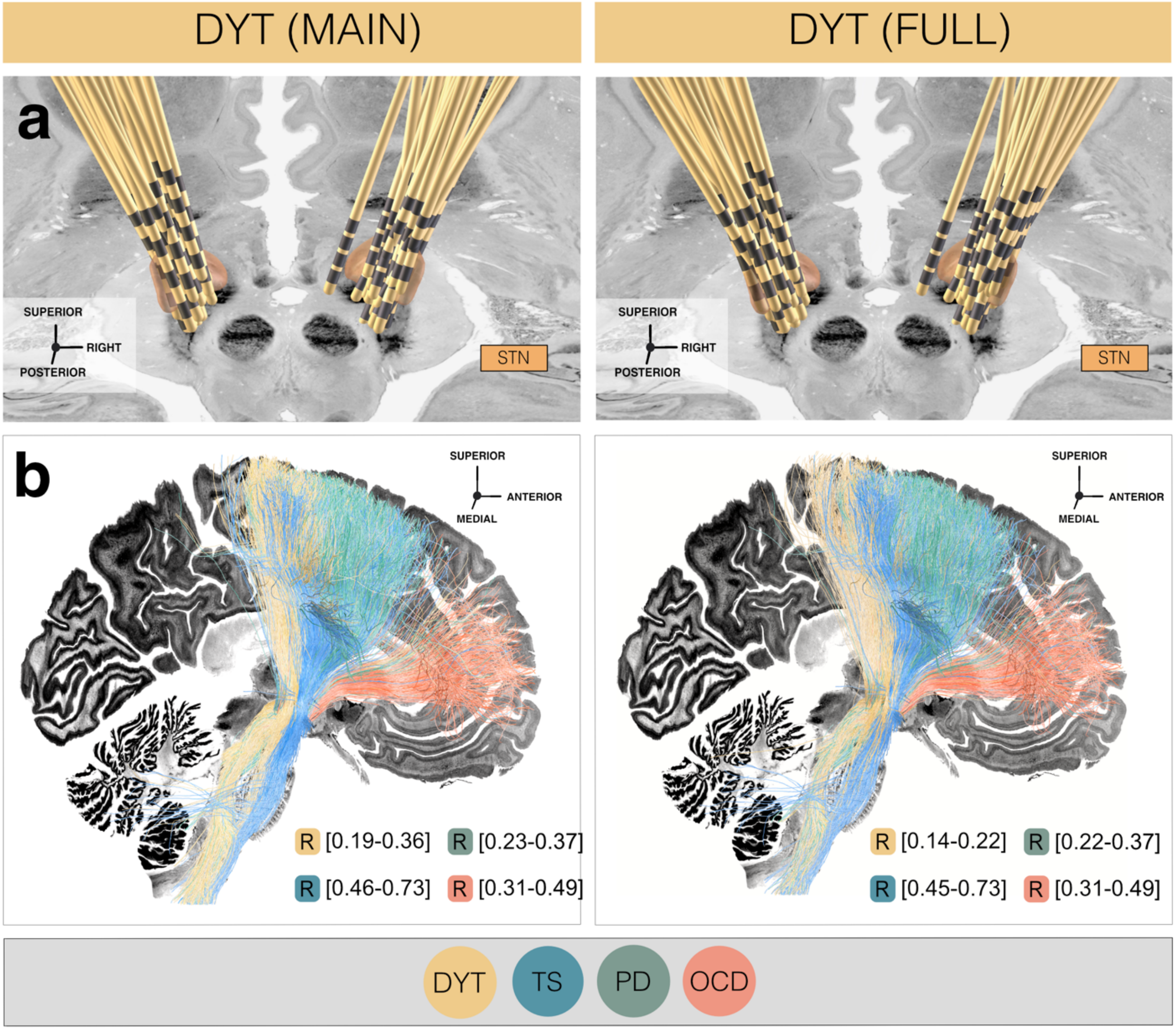


**Fig. S9: Comparable results based on restricted vs. unrestricted dystonia samples.** Respectively left panels show anatomical electrode placement in the restricted (main) dystonia (DYT) sample relative to the subthalamic nucleus (STN) **(a)**, as well as disease-wise deep brain stimulation (DBS) Fiber Filtering **(b)** results. Data of DYT patients reported within the main body of the manuscript followed a more conservative scheme of exclusion criteria. In this case, only patients with baseline scores in the Burke-Fahn-Marsden Dystonia Rating Scale (BFMDRS) ≥ 5 and follow-up ≥ 6 months were considered (N = 56) to ensure comparable and sufficiently stabilized stimulation effects across cohorts. The respectively right panels show electrode placement and results comprising the unrestricted (full) DYT sample (N = 70) using the N = 14 additional patients from Shanghai. STN defined by the DBS Intrinsic Template (DISTAL) atlas [3], with an axial plane of the BigBrain template in 100 μm resolution [2] displayed as a backdrop. *Abbreviations:* TS, Tourette's syndrome; OCD, obsessive-compulsive disorder; PD, Parkinson's disease.

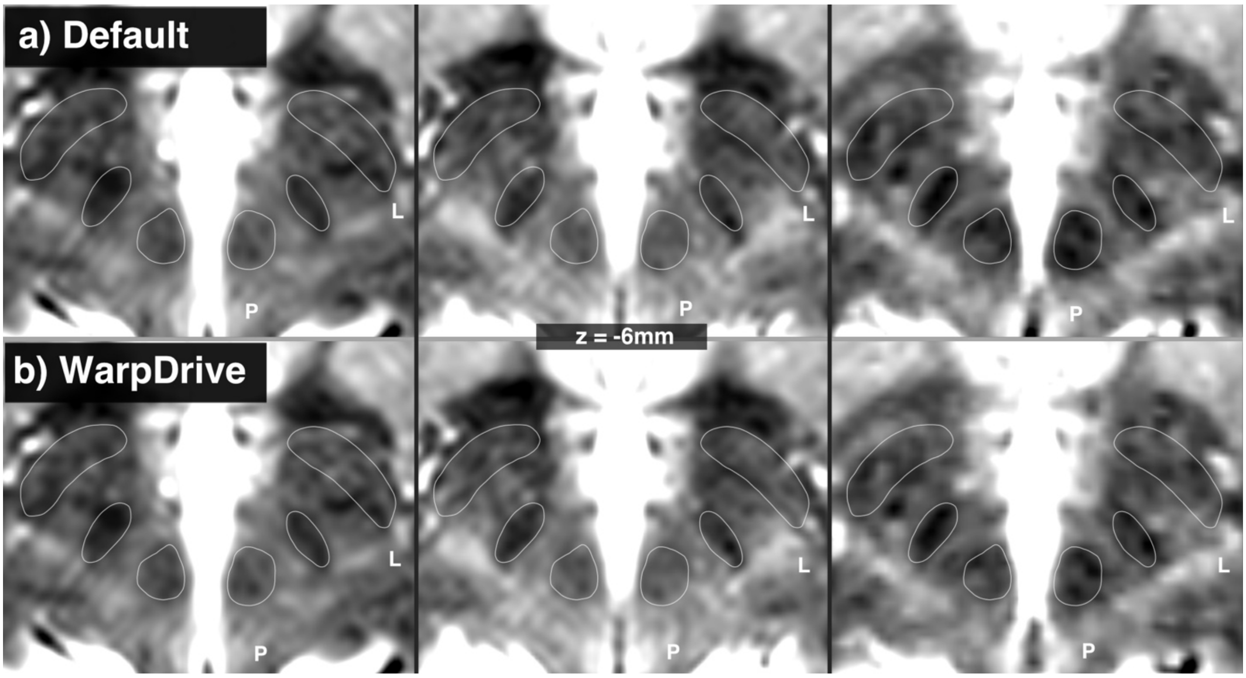


**Fig. S10: Optimization of normalization warp fields at the subthalamic level.** Three example cases of registrations between atlas template and individual brain anatomy are shown. Panel **(a)** demonstrates results following automatized normalization. Panel **(b)** shows results after the additional manual refinement of small mismatches with focus on the subthalamic nucleus (STN) region using the WarpDrive toolbox [9,10] as implemented in Lead-DBS, v3 software [10]. Outlines of displayed atlas structures are defined based on the DBS Intrinsic Template (DISTAL) atlas [3]. In most cases, no substantial changes needed to be carried out (such as in the leftmost case, in which automated results were excellent). In others, small refinements to better fit the patient STN to the atlas STN were made using WarpDrive.

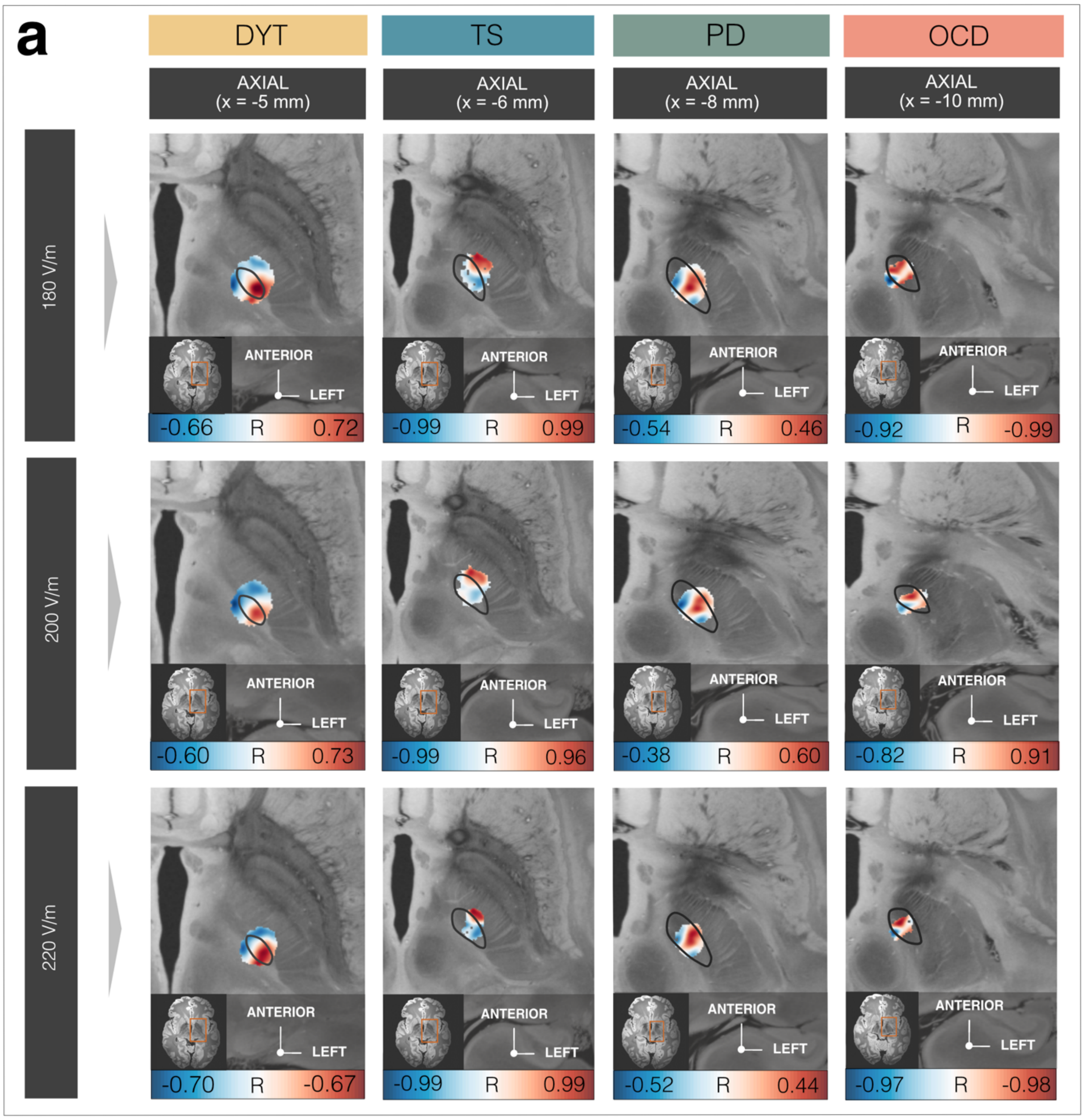


**b**

| E-FIELD THRESHOLD | IN-SAMPLE | 5-FOLD CV | IN-SAMPLE | 5-FOLD CV | IN-SAMPLE | 5-FOLD CV | IN-SAMPLE | 5-FOLD CV |
|---|---|---|---|---|---|---|---|---|
| 180 V/m | R = 0.57; p < 1e-3 | R = 0.42; p = 0.002 | R = 0.48; p < 1e-3 | R = 0.38; p < 1e-3 | R = 0.81 p < 1e-3 | R = -0.49; n.s. | R = 0.52; p = 0.027 | R = 0.36 p = 0.129 |
| 200 V/m | R = 0.56; p < 1e-3 | R = 0.41; p = 0.001 | R = 0.49; p < 1e-3 | R = 0.41; p < 1e-3 | R = 0.70; p = 0.006 | R = -0.51; n.s. | R = 0.44; p = 0.061 | R = 0.53; p = 0.023 |
| 220 V/m | R = 0.56; p < 1e-3 | R = 0.40; p = 0.001 | R = 0.49; p < 1e-3 | R = 0.44; p < 1e-3 | R = 0.71; p = 0.005 | R = -0.50; n.s. | R = 0.47; p = 0.046 | R = 0.52; p = 0.0025 |

**Fig. S11: Influence of electrical field threshold on sweet spot mapping results. (a)** Sweet and sour spot configurations remain consistent across different electrical field (E-field) thresholds when remaining model parameters are kept consistent. Relative anatomical position of disease-wise maps based on E-field thresholds of 180 V/m, 200 V/m, and 220 V/m are shown in axial view of a comparable slice per disorder. Sweet and sour spots are presented with regard to the left subthalamic nucleus (STN; black outlines) in template space, as derived from the DBS Intrinsic Template (DISTAL) atlas [3], and in superposition to an 100 μm ex-vivo template [11]. Color-coding of voxels is representative of correlation strength (warm colors for positive and cool colors for negative associations) between E-field magnitudes and clinical improvements. **(b)** In-sample correlations as well as five-fold cross-validation (CV) results are comparable with results reported in the main manuscript. *Abbreviations:* CV, cross-validations; TS, Tourette's syndrome; OCD, obsessive-compulsive disorder; PD, Parkinson's disease.

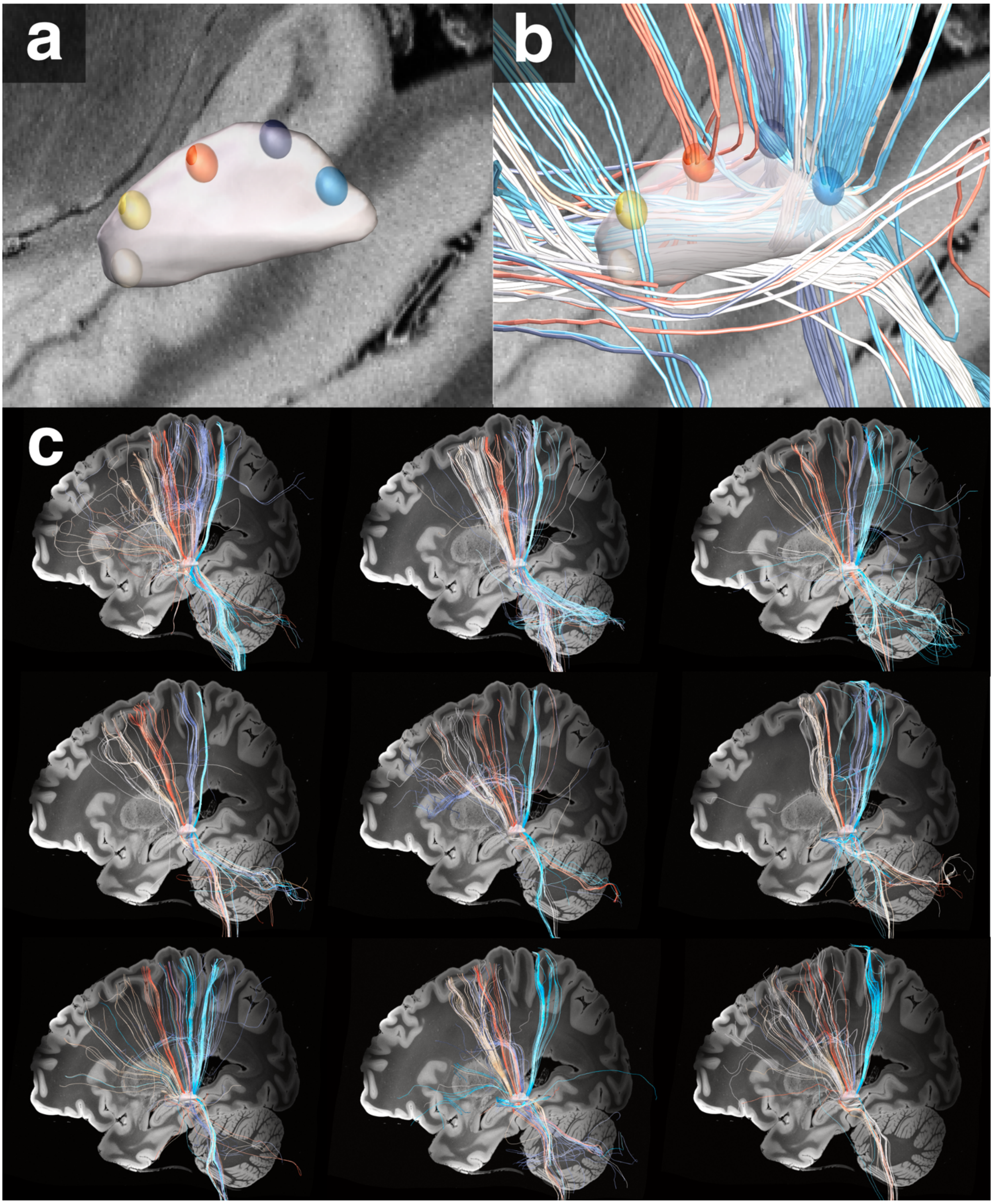


**Fig. S12: Comparison of the topographical organization of cortico-subthalamic interconnections across individuals.** (**a)** To interrogate the degree of inter-individual variability in cortico-subthalamic anatomical connectivity, five seed points were placed along the dorsal convex shape of the subthalamic nucleus (STN) in standard space, as represented by a three-dimensional model of the left STN from the DBS Intrinsic Template (DISTAL) atlas [3]. **(b)** Streamlines were tracked from these seed points in nine randomly selected healthy participants of the Human Connectome Project (HCP) [5], respectively. **c)** Despite similarities in the general topographical organization across individual brains, visible differences in the granularities of streamline representations emerged (as shown in color-coded fashion). Streamline tracking results are displayed against a sagittal slice (x = -30 mm) of the 7T MRI ex-vivo 100 µm human brain template [11].

### Supplementary Methods

#### Manual refinement of normalization displacement fields for electrode reconstruction

High precision in the registration between a template brain with the brain anatomy of an individual represents one of the most important requirements to accurately reconstruct electrode localizations. Although the precision of automated normalization algorithms is steadily increasing [12,13], the accuracy of registrations between a template and an individual's brain anatomy can vary depending on the neuroanatomical structure in focus. Especially the low contrast of basal ganglia structures on imaging sequences typically applied in clinical routine care renders the registration between an individual subthalamic nucleus (STN) and atlas STN challenging [13,14].

"WarpDrive" [9,10] is conceived as a dedicated but optional module (accessible from within Lead-DBS software v3.0 [10]) that allows to manually counteract small misalignments after automated normalization has been performed. By means of three different tools (point-to-point or line-to-line fiducials as well as a smudge application), a mismatched warp field can be precisely optimized through manual interaction by an anatomically experienced user. The manually placed fiducials are applied with a Gaussian smooth kernel and finally fed into the Plastimatch software [15] (accessed from within WarpDrive by a Slicer command line module). By means of the resulting transform, electrode and contact positions are updated to achieve a precise(r) mapping between template space and electrode implantation coordinates.

In the present study, wherever mismatches in the registration were clearly visible following careful visual inspection, manual refinements of the atlas fit were performed based on WarpDrive software building on automated normalization results. These consisted of a multispectral spatial normalization into ICBM 2009b Non-linear Asymmetric ("MNI") template space [16] of the Symmetric Normalization (SyN) approach included in ANTs with the "effective: low variance + subcortical refinement" preset in Lead-DBS. The STN was defined based on the Deep Brain Stimulation (DBS) Intrinsic Template (DISTAL) atlas [3]. **Fig. S10** displays three example cases of refinements to the atlas fit through careful use of the WarpDrive tool in addition to automated normalization (ANTs + WarpDrive) when compared to the results of the automated normalization algorithm alone (ANTs only). Across the entire cohort, displacements of 0-1 mm were applied in N = 304 electrodes, of 1-2 mm in N = 86 electrodes, and of >2 mm in N = 16 electrodes.

#### Creation of the DBS Tractography Atlas, v2

To validate our streamline segregation results and scrutinize the influence of specific characteristics of different connectomic resources, we repeated our analysis based on four normative connectomes of healthy human brains. One of these consisted of a custom-curated pathway atlas including a comprehensive description of subthalamic interconnections with multiple cortical and subcortical nodes based on a finite set of 6,525,876 streamlines. It was created specifically to represent streamlines that had previously lacked delineation in other resources and was informed by both streamline tracking and other (previously established) pathway atlases.

To generate this atlas resource, a first subset of streamlines connecting the STN to different cortical regions was derived via streamline tracking based on the HCP-1,065 diffusion data, which scanned 1,065 young and healthy adults [17]. This data is openly available within DSI Studio (https://sites.google.com/a/labsolver.org/brain/diffusion-mri-data/hcp-dmri-data). Using DSI Studio (https://dsi-studio.labsolver.org/), the STN as defined within the DISTAL atlas [3] was specified as an end region, and 1,500 streamlines were tracked from each of nine cortical Brodmann areas (BAs) as regions of interest from the digitized Brodmann atlas [18]. These comprised BA1/2/3 (primary and secondary somatosensory cortex), BA4 (primary motor cortex), BA6 (supplementary motor area), BA10 (fronto-parietal cortex), BA13 (insular cortex), BA24/32 (cingulate cortex), BA25 (subgenual anterior cingulate cortex), as well as BA45/47 (frontal gyrus). Two further ROIs of the subcortical region – the substantia nigra pars compacta and pars reticulata – were added from the California Institute of Technology reinforcement learning atlas (CIT168) [19].

To enable cortical branching, the angular threshold was set to a range of 60°-90°. Sampling was thresholded at a minimum length of 5 mm to avoid the inclusion of short streamlines and to prioritize long-range cortico-subthalamic projections. Streamline tracking between STN and substantia nigra aspects was performed based on a minimal tracking length of 10 mm, considering the distance between these two regions. To account for the exploratory nature of these connections, 20 iterations of topology-informed pruning were further implemented to limit the possibility of including false-positive streamlines [20]. Because of the role of these cortical regions in neuromodulation for affective disorders [21], streamline tracking between BA24/25/32 was seeded from the limbic aspect of the STN (based on its definition within the DISTAL atlas [3]). In this case, a two-fold dilation of the limbic STN was implemented in order to allow for limbic regions adjacent to the anterior STN to be included [22], and streamlines were mirrored between hemispheres (thus increasing the streamline count per sampling to a total of 3,000) to limit the occurrence of potential spurious lateralization effects.

In addition, the creation of this anatomically inclusive pathway atlas was complemented by representations of the anterior thalamic radiation, the cerebellothalamic tract, the dentato-rubro-thalamic tract, as well as the fasciculus subthalamicus from the DBS Tractography Atlas, v1 [23]. Finally, definitions of the ansa and fasciculus lenticularis as well as subthalamic-pallidal connections between STN and all pallidal nuclei were added from the Basal Ganglia Pathway Atlas [7], comprising interconnections between STN and internal pallidum (GPi), STN and external pallidum (GPe), Gpi and STN (associative and somatomotor aspects), as well as Gpe and STN (associative and somatomotor aspects).

**Creation of Disease-Matched Connectomes**

To probe the generalizability of our findings to connectomes altered in a disease-specific fashion, we repeated the main DBS Fiber Filtering analysis based on two exemplary disease-matched connectomes calculated on dMRI data of one cohort of OCD and PD patients each.

For this purpose, we first informed disease-wise streamline models on a whole-brain PD group connectome previously calculated [3] on the basis of dMRI data of N = 85 PD patients (28 female; mean age = 59.48 ± 10.39 years) openly available from the Parkinson's Progression Markers Initiative [8] (PPMI; www.ppmi-info.org). This connectome has been repeatedly used to inform analyses within the DBS context [4,24–28]. Details on scanning parameters can be derived from the project website (www.ppmi-info.org) and specifics on the creation of the resulting PD group connectome are reported elsewhere [3]. In brief, a fast diffeomorphic image registration algorithm [29] (Diffeomorphic Anatomical Registration Through Exponentiated Lie Algebra, DARTEL), as implemented in SPM12 (http://www.fil.ion.ucl.ac.uk/spm/software/spm12/) and Lead-DBS [10,30,31] (www.lead-dbs.org) was applied per subject, to estimate a nonlinear deformation field into ICBM 2009b NLIN asymmetric space based on T2-weighted acquisitions. To determine global streamline sets, a generalized q-sampling approach [32] was run as available in DSI-Studio (http://dsi-studio.labsolver.org/). Specifically, 20,000 streamlines were sampled for each patient, using seeds within a white-matter mask derived from segmenting the T2-weighted acquisitions using SPM12. Subsequently, the streamline set per patient was standardized into MNI space, employing the methodology described in [33,34].

Second, the same DBS Fiber Filtering analysis was repeated in an OCD group connectome that was explicitly calculated for the purpose of the current manuscript. This connectome was derived from diffusion scans acquired before surgery in the N = 6 OCD patients (1 female; mean age = 45.50 ± 10.52 years) from the London cohort analyzed in the present study. The supplementary materials of the original publication [35] contain more detailed information on scanning parameters. In brief, transformations from patient space into ICBM 2009b NLIN asymmetric space were derived using Advanced Normalization Tools (ANTs, http://stnava.github.io/ANTs/) [36]. dMRI data were preprocessed using FMRIB Software Library (FSL) [37] (topup and eddy). A generalized q-sampling approach [32] was performed as implemented in DSI-Studio (http://dsi-studio.labsolver.org/). 500,000 streamlines were deterministically tracked per patient with an angular threshold of 30 and a smoothing factor of one, while default settings were kept for all remaining parameters. The organization of dysfunction mappings resulting when the DBS Fiber Filtering analysis was informed on either of these PD or OCD group connectomes remained largely consistent to the one seen in the normative connectomes of healthy brains (**Figure S8a & b**). The same was the case for in-sample correlations and five-fold CVs (**Figure S8c**).

**Narrative Description of Model Set-Up and Validation**

Sweet spots (i) and sweet streamlines (ii) were modeled independently of each other, for each disease cohort – and for each connectome in the case of DBS Streamline Filtering – separately.

For the creation of both these models per disease cohort, patient-wise relative clinical improvement scores within the respective primary outcome assessment were integrated (Burke-Fahn-Marsden Dystonia Rating Scale in DYT, Unified Parkinson's Disease Rating Scale in PD, Yale-Global Tic Severity Scale in TS, and Yale-Brown Obsessive-Compulsive Scale in OCD). Relative improvement was calculated by comparing scores at baseline (i.e., before surgery or during the postoperative OFF DBS state in the case of PD) to postoperative DBS ON conditions and dividing by the score at baseline, resulting in a vector of clinical improvement values of the length of the respective patient sample. Besides clinical outcome, information contributed by patients comprised the stimulation impact on the respective aspect of neuroanatomy in focus (subthalamic voxels or a set of streamlines within the considered connectome). This stimulation impact on neuroanatomy was determined in the following fashion:

- i) *Sweet spots:* Per patient, an electric field (E-field) was modeled in the patient's native space and warped into template space (ICBM 2009b Non-linear Asymmetric "MNI" space). This E-field was represented in the form of a three-dimensional NIfTI volume in space. On a voxel-by-voxel basis, the E-field magnitude in each voxel encompassed by the E-field volume was then denoted for each patient across the cohort. Importantly, not all voxels in the brain were covered by an equal number of E-fields. Since this may bias results toward unrepresentative models, only voxels touched by at least half of E-fields across the respective disease cohort were considered. For each voxel in the brain, this procedure resulted in a vector of E-field magnitude values of the length of the respective patient sample.
- ii) *Sweet streamlines:* The connectomic modeling approach was performed on a streamline-by-streamline basis. Only the subset of streamlines from the respective connectome was considered that received high modulation (> 0.8 V/mm, i.e., passing in proximity to active electrode contacts) by at least a minimal number of patients (more than 0.5% of E-fields contained within the cohort). The peak E-field amplitude by which each streamline of this subset was modulated was subsequently derived per patient. This was achieved by sampling E-field magnitudes from multiple points along the course of each tract and denoting the magnitude of the point at which the highest impact had been detected. For each streamline, this procedure resulted in a vector of peak E-field magnitude values of the length of the respective patient sample.

In the second step, i) an optimal focal stimulation target (sweet spot) as well as ii) an optimal set of sweet streamlines were modeled that were linked to maximized clinical improvements within each disease-wise primary outcome measure. This procedure was implemented in the following manner:

- *i) Sweet spots:* Again, the procedure was applied on a voxel-by-voxel basis. For each brain voxel, the vector of E-field magnitudes was rank correlated with the vector of clinical improvements across patients. Iterating through all brain voxels, this procedure led to an R-map which represents the degree to which the modulation of each voxel via DBS was related to clinical improvement. The resulting model can be conceived as an optimal map of where E-fields should ideally stimulate the focal anatomy to maximize treatment success within the respective domain of dysfunction. This Sweet Spot Mapping approach was first introduced in Horn et al. (2022) [38].
- *ii) Sweet streamlines:* Connectivity-based modeling was again performed in a streamline-by-streamline fashion. For each streamline, the vector of peak E-field magnitudes by which the respective streamline had been modulated was Spearman's rank correlated with the vector of clinical improvements across patients. Iterating through all streamlines of the respective connectome, this procedure resulted in one R-value per tract. This R-value indicates the degree to which the modulation of each tract via DBS was related to clinical improvements. The resulting streamline profile can be seen as an optimal model of which streamlines E-fields should ideally stimulate to maximize treatment success within the respective dysfunctional domain. This DBS Fiber Filtering approach was first introduced in Irmen et al. (2019) [4].

Once optimal sweet spot (i) and sweet streamline models (ii) had been established for each disease cohort (and for each connectomic resource in the case of DBS Fiber Filtering) separately, each model was tested individually for their explanatory value for clinical outcome variance within the respective discovery cohort in question. This was achieved in the following way:

- *i) Sweet spots:* Each E-field of an unseen patient (i.e., a patient that had not been used to inform the model) was spatially compared to the R-map model. To do so, E-field magnitudes were multiplied with previously assigned R-values for each voxel within the model, and results were finally averaged across voxels. This led to the attribution of one "Sweet Spot Score" per E-field and scores were finally averaged across bilateral E-fields to achieve one single Sweet Spot Score per patient. This model is conceived to represent a set of "optimal" voxels whose stimulation leads to maximal clinical improvements within the modeling cohort. Consequently, high agreement of the E-fields of a novel (unseen) patient with this sweet spot model (leading to the attribution of a high Sweet Spot Score) should be associated with excellent clinical outcome. The opposite pattern should hold true for the case of small or no overlap of the patient's stimulation volumes with the sweet spot pattern (leading to the attribution of a low Sweet Spot Score and suboptimal clinical outcome).

- *ii) Sweet streamlines:* Again, each E-field of an unseen patient (i.e., a patient that had not been used to inform the model) was considered separately. The sweet streamline model taken into consideration for model validation was further restricted to only such streamlines that had been tagged by particularly high R-values (based on a threshold set at the top 1% of the cumulative distribution function of R-values of all sweet streamlines within the model). This preselection step was intended to only include such streamlines where high modulation led to high improvement, i.e., to streamlines with high meaningfulness for the model, while potentially less relevant or noisy correlations were discarded. Thereafter, only the subset of streamlines from this preselected profile was retained that was covered by that E-field. Iterating through this subset of streamlines, the peak E-field magnitude was multiplied with the previously assigned R-values of each streamline. This was done to account for (or "weigh R-values by") the modulation strength above and beyond the mere "connection status" of the E-field with the sweet streamline. Moreover, only the peak 5% of these weighted sweet streamline R-values were retained. This step was implemented to maximally account for the most impactful and impacted on streamlines by that E-field. These top 5% of weighted R-values were then summed up, so that each E-field was tagged by one "weighted peak 5% of Fiber R-Scores". Finally, this score was averaged across bilateral E-fields, leading to one single value per patient. Since the model is assumed to represent a set of "optimal" streamlines whose stimulation leads to maximal clinical improvements within the model cohort, E-fields with high overlap with these sweet streamlines (leading to the attribution of high weighted peak 5% of Fiber R-Scores) should be associated with good clinical outcomes for the respective patients. On the contrary, E-fields with only peripheral or no overlap with the model (leading to the attribution of low weighted peak 5% of Fiber R-Scores) would receive low treatment benefit.

Thus, the tenability of the sweet spot and sweet streamline models for estimating clinical outcome could be probed by correlating Sweet Spot Scores – or weighted peak 5% of Fiber R-Scores – with empirical clinical improvements across patients, with higher resulting correlation coefficients representative of increased model validity. Within the discovery sample, we did so based on two strategies. For in-sample analyses, sweet spot and sweet streamline models were derived based on E-fields of all patients within each disease cohort and validated using each patient of the respective cohort (circular analysis). For five-fold CVs, each patient cohort was randomly split into five folds. The model was built on four folds of patients and validated on the remaining fold. This strategy was repeated five times so that validation was performed for all patients.

Above and beyond model validations based on the discovery cohort, PD and OCD streamline models were further validated in an additional retrospective patient sample. Crucially, these patients provided entirely independent data points that had not been used to inform the streamline model (see methods of the main manuscript for the exception of OCD patients from London). In both cases, weighted peak 5% of Fiber R-Scores were determined for each patient within the novel cohort in the same fashion as described above (i.e., based on the overlap of stimulation volumes of novel patients with the streamline model that had been built on the respective discovery cohort). Correlations of these scores with empirical clinical improvements across novel patients could finally be used as a means of validating the ability of the streamline model to estimate clinical outcome variance in unseen data.

### Supplementary Results

#### Influence of Electrical Field Thresholds on Sweet Spot Mappings

Sweet Spot Mapping reported in the main body of the manuscript had been calculated based on an E-field magnitude threshold of 200 V/m. Although this value represents a commonly assumed magnitude of voltage needed to activate axons [27,39,40] and has been repeatedly successfully applied in models of DBS effects on anatomy surrounding the active electrode contact across disorders [10,31,41–45], the choice of this parameter has the potential to influence results. To demonstrate stability of sweet spot configurations and five-fold CVs across different E-field magnitudes, we repeated Sweet Spot Mapping at magnitudes of 180 V/m and 220 V/m. Sweet and sour spot maps as well as in-sample correlations and five-fold CVs remained largely consistent across these thresholds (see **Fig. S11**).

#### Interindividual Anatomical Variability in Cortico-Subthalamic Connections

To assess the general topography of interconnections between cortex and STN, we seeded streamlines from five seed points that were manually placed along the dorsal convex shape of the STN in standard space (**Fig. S12a**). We normalized streamlines that originated from nine random individual HCP subjects and selected streamlines that connected to each of the seed points in individualized connectomes (**Fig. S12b**). This made it possible to visually compare the topography of STN connections. While, as expected, the general fronto-rostral topography of interconnections between cortex and STN was revealed in all nine individuals (**Fig. S12c),** the results also show individual variance. In this context, we must mention that only an unknown fraction of these differences should be attributed to true anatomical variance. As shown by several authors using test-retest analysis of brains that were scanned multiple times with dMRI, a substantial fraction of individual differences needs to be attributed to noise and distortions in the data, the choice of MRI machine and tractography algorithms [46,47].
