## Supplementary Tables for "Mapping Dysfunctional Circuits in the Frontal Cortex Using Deep Brain Stimulation"

### Table of Contents


**Supplementary Tables** **p. 2**

**Table S1:** Summary of demographic and clinical patient characteristics within each discovery cohort. **p. 2**

**Table S2:** Patient-wise demographic and clinical characteristics of the dystonia discovery cohorts. **p. 4**

**Table S3:** Patient-wise demographic and clinical characteristics of the Tourette's syndrome discovery cohorts. **p. 7**

**Table S4:** Patient-wise demographic and clinical characteristics of the Parkinson's disease discovery cohorts. **p. 8**

**Table S5:** Patient-wise demographic and clinical characteristics of the obsessive-compulsive disorder discovery cohorts. **p. 12**

**Table S6.** Summary of demographic and clinical patient characteristics within each retrospective model validation cohort. **p. 13**

**Table S7:** Patient-wise demographic and clinical characteristics of the Parkinson's disease validation cohorts. **p. 15**

**Table S8:** Patient-wise demographic and clinical characteristics of the obsessive-compulsive disorder validation cohorts. **p. 17**

**Table S9:** Peak voxel coordinates of subthalamic sweet spots and of disease-specific cortical sites of interconnection with sweet tracts. **p. 19**

**Table S10:** Overlap between connected streamlines shared among disorders and disease-specific sweet streamlines. **p. 20**

**Table S11:** Patient-wise demographic and clinical characteristics of patient cases for prospective model validations. **p. 20**

**References** **p. 22**

**Table S1:** Summary of demographic and clinical patient characteristics within each discovery cohort.

| **Disease cohort** | **DYT** | | **PD** | | **TS** | | **OCD** | |
|---|---|---|---|---|---|---|---|---|
| | | | | **Demographic Information** | | | | |
| **Cohort** | San Francisco | Shanghai | Berlin | Würzburg | Pisa/Milan | Shanghai | London | Grenoble |
| **Surgical DBS center** | University of California San Francisco | Ruijin Hospital Shanghai | Charité - Universiäts-medizin Berlin | University Hospital Würzburg | Fondazione IRCCS Instituto Neurologico Carlo Besta Milan | Ruijin Hospital Shanghai | National Hospital for Neurology and Neurosurgery | University Hospital Grenoble |
| **N (female)** | 12 (7) | Main*: 44 (23); Full: 58 (31) | 51 (17) | 43 (12) | 4 (2) | 10 (1) | 6 (1) | 13 (9) |
| **Age at time of surgery (mean ± SD; range; in years)** | 56.08 ±16.06; 16-68 | Main*: 39.43 ± 21.06, 7-74; Full: 41.90 ± 20.60, 7-74 | 59.98 ± 7.93.; 42-75 | 60.49 ± 8.23; 46-74 | 34.00 ± 8.83; 26-46 | 23.10 ± 9.39; 14-38 | 45.50 ± 10.52; 37-62 | 39.15 ± 8.23; 27-53 |
| **Disease duration at time of surgery (mean ± SD; range; in years)** | 9.50 ± 9.54; 2-32 | Main*: 5.94 ± 7.01, 0.5-30; Full:5.15 ± 6.36, 0.25-30 | 10.38 ± 3.85; 5-21 | 12.53 ± 4.46; 5-23 | 24.25 ± 6.08; 19-33 | 14.8 ± 9.38; 3-30 | 24.17 ± 3.92; 20-30 | 18.92 ± 8.96; 5-39 |
| | | | | **Clinical Outcome** | | | | |
| **Main clinical outcome assessment** | BFMDRS [1] (total score; range 0-120, with higher scores corresponding to higher symptom burden) | | UPDRS-III [2] (total score; range 0-199, with higher scores corresponding to higher symptom burden) | | YGTSS [3] (global severity score; range 0-100, with higher scores corresponding to higher symptom burden) | | Y-BOCS [4] (total score; range 0-40, with higher scores corresponding to higher symptom burden) | |
| **Time of FU (in months)** | 6 | Main*: mean = 19.09 ± 15.60 SD, range 6-84; Full: mean = 18.00 ± 15.56 SD, range 2-84 | 12 | 12 | 6 | 6 | 3 or 6 (post-amSTN phase)[+] | 12 |
| **Comparison basis for calculation of improvement values (baseline to DBS stimulation ON conditions)** | Pre- to postoperative | Pre- to postoperative | Postoperative ON vs. OFF stimulation (ON dopaminergic medication) | Postoperative ON vs. OFF stimulation (ON dopaminergic medication) | Pre- to postoperative | Pre- to postoperative | Pre- to postoperative | Pre- to postoperative |

| | | | | | | | | |
|---|---|---|---|---|---|---|---|---|
| **Score at baseline (mean ± SD)** | 16.88 ± 9.30 | Main*: 19.09 ± 17.68; Full: 16.40 ± 16.57 | 38.56 ± 12.92 | 49.84 ± 12.36 | 89.25 ± 5.25 | 69.70 ± 10.19 | 36.17 ± 1.83 | 33.54 ± 3.76 |
| **Score at time of FU under stimulation ON condition (mean ± SD)** | 6.88 ± 5.90 | Main*: 6.02 ± 9.21; Full: 5.42 ± 8.33 | 20.12 ± 8.82 | 24.47 ± 10.62 | 34.25 ± 15.97 | 26.20 ± 17.92 | 19.83 ± 10.57 | 18.92 ±10.74 |
| **Rel. improvement (mean ± SD; in %)** | 52.07 ± 41.71 | Main*: 69.90 ± 23.46; Full: 65.24 ± 28.81 | 45.34 ± 23.03 | 49.46 ± 23.97 | 61.50 ± 17.81 | 62.31 ± 26.21 | 45.00 ± 29.33 | 43.59 ± 31.56 |
| **Abs. improvement (mean ± SD)** | 10.00 ± 10.83 | Main*: 13.07 ± 14.72; Full: 10.97 ± 13.72 | 18.44 ± 11.31 | 25.37 ± 13.30 | 55.00 ± 17.00 | 43.50 ± 18.61 | 16.33 ± 10.67 | 14.62 ± 10.75 |
| **Imaging and DBS Specifications** | | | | | | | | |
| **Imaging modality (postoperatively)** | MRI (N = 12) | Main*: CT (N = 44); Full: CT (N = 58) | MRI (N = 45), CT (N = 6) | CT (N = 43) | CT (N = 4) | CT (N = 10) | MRI (N = 6) | MRI (N = 10), CT (N = 3) |
| **Electrode models** | MDT 3389 (N = 12) | Main*: MDT 3387 (N = 17), PINS L302 (N = 17), SR 1210 (N = 10); Full: MDT 3387 (N = 22), PINS L302 (N = 26), SR 1210 (N = 10) | MDT 3389 (N = 51) | MDT 3389 (N = 44) | MDT 3389 (N = 4) | MDT 3387 (N = 2), SR 1210 (N = 7), PINS L302 (N = 1) | MDT 3389 (N = 6) | MDT 3389 (N = 14) |
| **Related citation** | Ostrem et al., 2011 [5]; 2016 [6] | Lin et al., 2019 [7]; He, 2021 [8] | Horn et al., 2017 [9]; 2019 [10] | Horn et al., 2017 [9]; 2019 [10] | Vissani et al., 2019 [11] | Dai et al., 2022 [12] | Tyagi et al., 2019 [13] | Polosan et al. 2019 [14] |

*Notes:* *The full sample of dystonia (DYT) patients from the Shanghai center (N = 58) is based on more liberal inclusion criteria compared to the sample reported on within the main manuscript (N = 44). More conservative exclusion criteria (baseline scores on the Burke-Fahn-Marsden Dystonia Rating Scale [BFMDRS] ≥ 5 and times of follow-up for postoperative assessments ≥ 6 months) were chosen for main analyses to ensure sufficiently stabilized and comparable deep brain stimulation (DBS) effects across cohorts. Results based on the full sample are reported in the supplements. +OCD patients from London were bilaterally implanted with DBS electrodes to two different target sites (four electrodes per patient) – the anteromedial subthalamic nucleus (amSTN) and the ventral capsule/ventral striatum (VC/VS). Patients were randomly assigned to either receive "pure amSTN" for the first three months or from months four to six postoperatively (and "pure VC/VS" for the remainder of the first six months). Model set-up (based on the discovery cohort) was informed on stimulation parameters and corresponding clinical scores taken after the amSTN phase, while model validation was performed based on VC/VS-stimulation related data points. AmSTN stimulation was applied via a Medtronic (MDT) model of type 3389, while MDT 3387 leads were implanted for stimulation of the VC/VS target zone (considered only for model validations, see below). *Abbreviations:* CT, computed tomography; DBS, deep brain stimulation; FU, follow-up; MRI, magnetic resonance imaging; OCD, obsessive-compulsive disorder; PD, Parkinson's disease; SD, standard deviation; SR, SceneRay; TS, Tourette's syndrome; UPDRS-III, Unified Parkinson's Disease Rating Scale – Part III; Y-BOCS, Yale-Brown Obsessive-Compulsive Scale; YGTSS, Yale Global Tic Severity Scale.

**Table S2:** Patient-wise demographic and clinical characteristics of the dystonia discovery cohorts.

**SAN FRANCISCO (N = 12)**

| Patient Nr. | Age at Surgery (within range, in years) | Sex | Disease Duration at Surgery (years) | BFMDRS Baseline (total) | BFMDRS at FU (total) | BFMDRS Percent Improvement at FU (%) | Time of FU (months) | Symptom Sites | Genetic Mutation | Postop. Imaging Modality |
|---|---|---|---|---|---|---|---|---|---|---|
| 1 | 41-45 | F | 8 | 19 | 2 | 89 | 6 | oromandibular, cervical | none | MRI |
| 2 | 46-50 | M | 7 | 14 | 14 | 0 | 6 | cervical | none | MRI |
| 3 | 41-45 | F | 2 | 34 | 1 | 97 | 6 | cervical, trunk | none | MRI |
| 4 | 36-40 | F | 9 | 8 | 3 | 63 | 6 | cervical | none | MRI |
| 5 | 51-55 | F | 3 | 12 | 3 | 75 | 6 | face, neck | none | MRI |
| 6 | 46-50 | F | 4 | 7 | 4 | 43 | 6 | neck | none | MRI |
| 7 | 51-55 | F | 32 | 10 | 4 | 60 | 6 | arms | DYT-1 mutation | MRI |
| 8 | 16-20 | M | 6 | 11,5 | 3,5 | 70 | 6 | arms, neck | DYT-1 mutation | MRI |
| 9 | 66-70 | M | 3 | 22 | 12,5 | 43 | 6 | cervical | none | MRI |
| 10 | 66-70 | F | 10 | 17 | 5,5 | 68 | 6 | mouth, neck, and left hand | none | MRI |
| 11 | 16-20 | M | 4 | 35 | 10 | 71 | 6 | right arm, neck | DYT-1 mutation | MRI |
| 12 | 51-55 | M | 26 | 13 | 20 | -54 | 6 | neck, arms | none | MRI |

**SHANGHAI (N = 44 in the main analysis, N = 58 in the supplemental analysis)**

| Patient Nr. | Age at Surgery (within range, in years) | Sex | Disease Duration at Surgery (years) | BFMDRS Baseline (total) | BFMDRS at Follow-Up (total) | Percent Improvement at Follow-Up (%) | FU Period (months) | Dystonia Classification | Genetic Mutation | Postop. Imaging Modality |
|---|---|---|---|---|---|---|---|---|---|---|
| 1 | 61-65 | F | 3 | 18 | 4,5 | 75 | 12 | segmental | none | CT |
| 2 | 41-45 | F | 1,5 | 8 | 5,5 | 31 | 15 | segmental | none | CT |
| 3 | 26-30 | F | 2 | 10 | 1 | 90 | 11 | multi-focal | none | CT |
| 4 | 21-25 | F | 5,3 | 9 | 2 | 78 | 12 | generalized | none | CT |
| 5 | 56-60 | M | 1,5 | 9 | 1,5 | 83 | 12 | segmental | none | CT |
| 6 | 56-60 | F | 1,5 | 10 | 2 | 80 | 18 | segmental | none | CT |

| | | | | | | | | | | |
|---|---|---|---|---|---|---|---|---|---|---|
| 7 | 61-65 | F | 1,5 | 19 | 7 | 63 | 18 | multi-focal | none | CT |
| 8 | 71-75 | M | 1 | 11 | 5 | 55 | 12 | segmental | none | CT |
| 9 | 66-70 | M | 10 | 6 | 0,5 | 92 | 12 | focal | none | CT |
| 10 | 6-10 | M | 2 | 26 | 6 | 77 | 9 | generalized | DYT-1 mutation | CT |
| 11 | 31-35 | F | 2 | 6 | 3 | 50 | 12 | focal | none | CT |
| 12 | 51-55 | M | 30 | 26 | 7 | 73 | 17 | multi-focal | DYT-24 mutation | CT |
| 13 | 6-10 | M | 2 | 34 | 36 | -6 | 12 | multi-focal | none | CT |
| 14 | 51-55 | M | 1,2 | 8 | 2 | 75 | 84 | focal | none | CT |
| 15 | 36-40 | F | 1 | 16,5 | 5 | 70 | 10 | segmental | none | CT |
| 16 | 36-40 | M | 6 | 47,5 | 1 | 98 | 8 | generalized | none | CT |
| 17 | 66-70 | M | 10 | 19,5 | 11 | 44 | 11 | segmental | none | CT |
| 18 | 6-10 | M | 6 | 24 | 4 | 83 | 24 | multi-focal | none | CT |
| 19 | 16-20 | M | 1,5 | 16 | 1,5 | 91 | 12 | generalized | none | CT |
| 20 | 21-25 | M | 1 | 5 | 1,5 | 70 | 10 | segmental | none | CT |
| 21 | 66-70 | F | 8 | 8,5 | 4,5 | 47 | 48 | segmental | none | CT |
| 22 | 66-70 | F | 1 | 11 | 3 | 73 | 30 | segmental | none | CT |
| 23 | 26-30 | M | 3,5 | 19 | 5 | 74 | 6 | generalized | none | CT |
| 24 | 41-45 | M | 3 | 10,5 | 4 | 62 | 24 | generalized | none | CT |
| 25 | 46-50 | F | 8 | 10 | 0,5 | 95 | 24 | segmental | none | CT |
| 26 | 41-45 | F | 15 | 8 | 2 | 75 | 30 | generalized | none | CT |
| 27 | 21-25 | F | 12 | 30 | 1 | 97 | 15 | multi-focal | none | CT |
| 28 | 11-15 | M | 2 | 21 | 2 | 90 | 18 | generalized | none | CT |
| 29 | 31-35 | M | 28 | 24 | 3 | 88 | 72 | generalized | none | CT |
| 30 | 51-55 | F | 6 | 16,5 | 4 | 76 | 7 | generalized | none | CT |
| 31 | 56-60 | F | 1,5 | 6 | 3 | 50 | 30 | focal | none | CT |
| 32 | 66-70 | F | 9 | 5 | 2,5 | 50 | 11 | segmental | none | CT |
| 33 | 51-55 | F | 9 | 49 | 49 | 0 | 12 | multi-focal | none | CT |
| 34 | 21-25 | F | 21 | 20 | 13 | 35 | 24 | segmental | none | CT |
| 35 | 16-20 | M | 2 | 92 | 19,5 | 79 | 24 | generalized | none | CT |
| 36 | 71-75 | M | 6,5 | 32,5 | 17 | 48 | 12 | segmental | none | CT |

| | | | | | | | | | | |
|---|---|---|---|---|---|---|---|---|---|---|
| 37 | 36-40 | M | 1,5 | 20 | 7,5 | 63 | 18 | generalized | none | CT |
| 38 | 6-10 | M | 1 | 74 | 11 | 85 | 10 | generalized | none | CT |
| 39 | 6-10 | F | 0,5 | 16 | 1 | 94 | 13 | multi-focal | none | CT |
| 40 | 16-20 | M | 1 | 9 | 1 | 89 | 12 | multi-focal | none | CT |
| 41 | 36-40 | F | 15 | 6 | 1 | 83 | 8 | segmental | none | CT |
| 42 | 11-15 | F | 2 | 8 | 1 | 88 | 9 | segmental | none | CT |
| 43 | 56-60 | F | 1 | 6,5 | 1,5 | 77 | 16 | segmental | none | CT |
| 44 | 21-25 | F | 14 | 9 | 1 | 89 | 36 | multi-focal | none | CT |
| *Patients in the following list are included only in the supplemental analysis.* | | | | | | | | | | |
| 45 | 66-70 | F | 2,5 | 3 | 1,5 | 50 | 12 | segmental | none | CT |
| 46 | 31-35 | M | 1,5 | 4 | 1 | 75 | 24 | segmental | none | CT |
| 47 | 11-15 | M | 1 | 26 | 14,5 | 44 | 3 | segmental | none | CT |
| 48 | 51-55 | F | 0,3 | 3,5 | 1,5 | 57 | 12 | segmental | none | CT |
| 49 | 51-55 | F | 1 | 6 | 2 | 67 | 2 | focal | none | CT |
| 50 | 66-70 | F | 8,5 | 3,5 | 3 | 14 | 7 | segmental | none | CT |
| 51 | 46-50 | M | 3,5 | 3,5 | 1 | 71 | 60 | segmental | none | CT |
| 52 | 31-35 | M | 3 | 28 | 3 | 89 | 4 | generalized | none | CT |
| 53 | 61-65 | M | 7 | 8 | 0,5 | 94 | 3 | segmental | none | CT |
| 54 | 61-65 | F | 4 | 4 | 1 | 75 | 12 | segmental | none | CT |
| 55 | 46-50 | M | 0,25 | 9 | 12 | -33 | 2 | segmental | none | CT |
| 56 | 71-75 | F | 3 | 4,5 | 5 | -11 | 24 | focal | none | CT |
| 57 | 26-30 | F | 0,5 | 4 | 3 | 25 | 24 | generalized | none | CT |
| 58 | 51-55 | F | 1 | 4 | 0,5 | 88 | 13 | multi-focal | none | CT |

*Abbreviations:* BFMDRS, Burke-Fahn-Marsden Dystonia Rating Scale; CT, computed tomography; F, female; FU, follow-up; M, male; MRI, magnetic resonance imaging; Nr., number; postop. postoperative.

**Table S3:** Patient-wise demographic and clinical characteristics of the Tourette's syndrome discovery cohorts.

**PISA/MILAN (N = 4)**

| Patient Nr. | Age at surgery (within range, in years) | Sex | Disease Duration at Surgery (years) | YGTSS Baseline | YGTSS at 6 mo. After Surgery | Percent Reduction from Baseline at 6 mo. (%) | Type of Tics | Comorbidities | Postop. Imaging Modality |
|---|---|---|---|---|---|---|---|---|---|
| 1 | 26-30 | F | 23 | 92 | 22 | 76 | pure motor | no OCD or major depression | CT |
| 2 | 26-30 | M | 19 | 89 | 20 | 78 | pure motor | no OCD or major depression | CT |
| 3 | 31-35 | F | 22 | 94 | 53 | 44 | pure motor | no OCD or major depression | CT |
| 4 | 46-50 | M | 33 | 82 | 42 | 49 | pure motor | no OCD or major depression | CT |

**SHANGHAI (N = 10)**

| Patient Nr. | Age at Surgery (within range, in years) | Sex | Disease Duration at Surgery (years) | YGTSS Baseline | YGTSS at 6 mo. After Surgery | Percent Reduction from Baseline at 6 mo. (%) | Motor Tics: Body Regions Involved | Comorbidities | Postop. Imaging Modality |
|---|---|---|---|---|---|---|---|---|---|
| 1 | 16-20 | M | 11 | 75 | 24 | 68 | neck, shoulders, legs, feet | none | CT |
| 2 | 11-15 | M | 9 | 71 | 4 | 94 | eyes, neck | none | CT |
| 3 | 11-15 | M | 3 | 68 | 6 | 91 | arms, trunk, legs | none | CT |
| 4 | 36-40 | M | 30 | 71 | 61 | 14 | nose, mouth, neck, arms, hands, legs | anxiety | CT |
| 5 | 16-20 | F | 12 | 59 | 8 | 86 | eyes, nose, mouth, neck, shoulders, arms, hands | OCD, depression | CT |
| 6 | 36-40 | M | 27 | 69 | 33 | 52 | eyes, mouth, neck, shoulders, arms, hands | none | CT |
| 7 | 16-20 | M | 6 | 76 | 36 | 53 | eyes, mouth, neck, hands, legs | none | CT |
| 8 | 26-30 | M | 16 | 63 | 17 | 73 | eyes, nose, shoulders, trunk | OCD | CT |
| 9 | 26-30 | M | 25 | 91 | 35 | 62 | neck, shoulders, arms, legs, feet | OCD | CT |
| 10 | 11-15 | M | 9 | 54 | 38 | 30 | eyes, mouth, neck | none | CT |

*Abbreviations:* CT, computed tomography; F, female; M, male; mo., months; Nr., number; OCD, obsessive-compulsive disorder; postop., postoperative; YGTSS, Yale Global Tic Severity Scale.

**Table S4:** Patient-wise demographic and clinical characteristics of the Parkinson's disease discovery cohorts.

| Pat. Nr. | Age at surgery (within range, in years) | Sex | Disease Duration at Surgery (years) | PD Phenotype* | UPDRS-III Total Preop. Baseline (OFF medication) | UPDRS-III Total Preop. Baseline (ON Medication) | UPDRS-III Total Postop. (STIM OFF at 12 Mo.) | UPDRS-III Total Postop. (STIM ON at 12 mo.) | UPDRS-III Total Percent Improvement ON vs OFF STIM at 12 mo. postop. (%) | LEDD Baseline (OFF DBS) | LEDD at 12 mo. Postop. (ON DBS) | LEDD Reduction (%) | Percent Improvement Levodopa Response | Postop. Imaging Modality |
|---|---|---|---|---|---|---|---|---|---|---|---|---|---|---|
| **BERLIN (N = 51)** | | | | | | | | | | | | | | |
| 1 | 71-75 | F | 8 | 1 | 20 | 10 | 29 | 30 | -3 | 968 | 498 | 49 | 50 | MRI |
| 2 | 56-60 | M | 10 | 1 | 33 | 20 | 41 | 23 | 44 | 900 | 725 | 19 | 39 | MRI |
| 3 | 66-70 | F | 12 | 1 | n/a | n/a | 54 | 36 | 33 | 1000 | 400 | 60 | 31 | MRI |
| 4 | 61-65 | M | 11 | 1 | 33 | 11 | 45 | 30 | 33 | 855 | 640 | 25 | 67 | MRI |
| 5 | 66-70 | M | 6 | 1 | 31 | 18 | 37 | 19 | 49 | 1035 | 1115 | -8 | 42 | MRI |
| 6 | 46-50 | M | 5 | 1 | 29 | 3 | 39 | 15 | 62 | 610 | 0 | 100 | 90 | MRI |
| 7 | 56-60 | F | 7 | 1 | n/a | n/a | 52 | 29 | 44 | 1150 | 100 | 91 | 72 | MRI |
| 8 | 71-75 | F | 18 | 1 | n/a | n/a | 13 | 11 | 15 | 1500 | 650 | 57 | 26 | CT |
| 9 | 66-70 | M | 12 | 1 | n/a | n/a | 40 | 20 | 50 | 1650 | 475 | 71 | 54 | CT |
| 10 | 61-65 | M | 11 | 1 | n/a | n/a | 44 | 26 | 41 | 900 | 600 | 33 | 46 | MRI |
| 11 | 66-70 | M | 9 | 1 | 27 | 9 | 36 | 22 | 39 | 1275 | 1305 | -2 | 67 | MRI |
| 12 | 51-55 | M | 7 | 1 | 36 | 20 | 46 | 30 | 35 | 1620 | 1000 | 38 | 44 | MRI |
| 13 | 66-70 | M | 14 | 1 | 25 | 14 | 28 | 17 | 39 | 1640 | 1398 | 15 | 44 | CT |
| 14 | 66-70 | F | n/a | 1 | n/a | n/a | 27 | 29 | -7 | n/a | n/a | n/a | n/a | MRI |
| 15 | 61-65 | M | 11 | 1 | 16 | 9 | 28 | 13 | 54 | 3065 | 400 | 87 | 44 | MRI |
| 16 | 66-70 | M | 10 | 1 | 32 | 17 | 29 | 19 | 34 | 1170 | 780 | 33 | 47 | MRI |
| 17 | 51-55 | M | 8 | 1 | n/a | n/a | 24 | 9 | 63 | 1000 | 267 | 73 | 85 | MRI |
| 18 | 56-60 | M | 7 | 1 | n/a | n/a | 32,5 | 8 | 75 | 600 | 50 | 92 | 79 | CT |
| 19 | 71-75 | M | 9 | 1 | n/a | n/a | 27 | 20 | 26 | 1300 | 700 | 46 | 43 | MRI |
| 20 | 71-75 | F | 5 | 1 | n/a | n/a | 24 | 25 | -4 | 650 | 700 | -8 | 44 | MRI |
| 21 | 51-55 | M | 16 | 1 | 36 | 16 | 33 | 6 | 82 | 700 | 375 | 46 | 56 | MRI |

| | | | | | | | | | | | | | | |
|---|---|---|---|---|---|---|---|---|---|---|---|---|---|---|
| 22 | 51-55 | M | 9 | 1 | 42 | 13 | 47 | 10 | 79 | 2000 | 553 | 72 | 69 | MRI |
| 23 | 61-65 | M | 14 | 1 | n/a | n/a | 28 | 15 | 46 | 1450 | 900 | 38 | 76 | MRI |
| 24 | 61-65 | F | 7 | 1 | n/a | n/a | 53 | 19 | 64 | 2050 | 600 | 71 | 30 | MRI |
| 25 | 61-65 | F | 8 | 1 | n/a | n/a | 46 | 28 | 39 | 700 | 200 | 71 | 71 | MRI |
| 26 | 61-65 | M | 7 | 1 | 21 | 12 | 28 | 13 | 54 | 1125 | 890 | 21 | 43 | MRI |
| 27 | 51-55 | M | 14 | 1 | n/a | n/a | 53 | 23 | 57 | 600 | 0 | 100 | 44 | MRI |
| 28 | 56-60 | F | 16 | 1 | n/a | n/a | 26 | 13 | 50 | 900 | 300 | 67 | 65 | MRI |
| 29 | 51-55 | M | 16 | 2 | 48 | 18 | 53 | 17 | 68 | 2438 | 423 | 83 | 63 | MRI |
| 30 | 56-60 | M | 13 | 1 | n/a | n/a | 59 | 14 | 76 | 1150 | 75 | 93 | 50 | MRI |
| 31 | 51-55 | F | 6 | 1 | 43 | 10 | 42 | 35 | 17 | 930 | n/a | n/a | 77 | MRI |
| 32 | 56-60 | M | 9 | 1 | n/a | n/a | 65 | 21 | 68 | 950 | 700 | 26 | 50 | MRI |
| 33 | 56-60 | M | 10 | 1 | n/a | n/a | 24 | 19 | 21 | 800 | 800 | 0 | n/a | MRI |
| 34 | 56-60 | F | 10 | 1 | n/a | n/a | 42 | 13 | 69 | 700 | 200 | 71 | 64 | MRI |
| 35 | 66-70 | M | 21 | 1 | 29 | 8 | 57 | 36 | 37 | 850 | 300 | 65 | 72 | MRI |
| 36 | 41-45 | M | 5 | 1 | n/a | n/a | 47 | 17 | 64 | 500 | 0 | 100 | 62 | MRI |
| 37 | 46-50 | F | 12 | 1 | n/a | n/a | 36 | 14 | 61 | 683 | 150 | 78 | n/a | MRI |
| 38 | 46-50 | F | 8 | 1 | n/a | n/a | 29 | 18 | 38 | 1400 | 0 | 100 | 52 | MRI |
| 39 | 56-60 | M | 7 | 1 | n/a | n/a | 21 | 9 | 57 | 333 | 50 | 85 | n/a | CT |
| 40 | 51-55 | M | 14 | 2 | 42 | n/a | 44 | 11 | 75 | 875 | 200 | 77 | n/a | MRI |
| 41 | 66-70 | M | 15 | 1 | 28 | 11 | 25 | 7 | 72 | 1328 | 360 | 73 | 61 | CT |
| 42 | 61-65 | M | 10 | 1 | 34 | 14 | 33 | 11 | 67 | 1100 | 720 | 35 | 59 | MRI |
| 43 | 66-70 | M | 17 | 2 | 60 | 21 | 71 | 40 | 44 | 1453 | n/a | n/a | 65 | MRI |
| 44 | 51-55 | F | 9 | 1 | n/a | n/a | 67 | 39 | 42 | 1200 | 375 | 69 | n/a | MRI |
| 45 | 41-45 | F | 12 | 2 | n/a | n/a | 40 | 27 | 33 | 1025 | 500 | 51 | 53 | MRI |
| 46 | 61-65 | M | 16 | 1 | n/a | n/a | 39 | 30 | 23 | 500 | 650 | -30 | 31 | CT |
| 47 | 51-55 | M | 11 | 1 | n/a | n/a | 32 | 10 | 69 | 400 | 0 | 100 | 32 | MRI |
| 48 | 61-65 | F | 10 | 1 | 37 | 12 | 35 | 26 | 26 | 945 | 688 | 27 | 68 | MRI |
| 49 | 71-75 | M | 6 | 2 | 18 | 16 | 33 | 21 | 36 | 400 | 800 | -100 | 11 | MRI |
| 50 | 51-55 | F | 5 | 1 | n/a | n/a | 45 | 13 | 71 | 750 | 50 | 93 | 25 | MRI |

| | | | | | | | | | | | | | | |
|---|---|---|---|---|---|---|---|---|---|---|---|---|---|---|
| 51 | 51-55 | M | 6 | 3 | 13 | 7 | 18 | 20 | -11 | 515 | 600 | -17 | 46 | MRI |
| **WÜRZBURG (N = 43)** | | | | | | | | | | | | | | |
| 1 | 51-55 | M | 13 | 1 | 78 | 46 | 78 | 36 | 54 | 1280 | 380 | 70 | 41 | CT |
| 2 | 56-60 | M | 13 | 1 | 53 | 25 | 53 | 28 | 47 | 1700 | 0 | 100 | 53 | CT |
| 3 | 56-60 | M | 20 | 1 | 43 | 12 | 43 | 15 | 65 | 2400 | 300 | 88 | 72 | CT |
| 4 | 66-70 | M | 16 | 1 | 49 | 18 | 49 | 30 | 39 | 1333 | 300 | 77 | 63 | CT |
| 5 | 46-50 | F | 21 | 1 | 36 | 3 | 36 | 33 | 8 | 1650 | 200 | 88 | 92 | CT |
| 6 | 56-60 | F | 15 | 1 | 43 | 13 | 43 | 25 | 42 | 1000 | 450 | 55 | 70 | CT |
| 7 | 61-65 | M | 15 | 1 | 43 | 8 | 43 | 23 | 47 | 1400 | 700 | 50 | 81 | CT |
| 8 | 66-70 | M | 16 | 1 | 55 | 21 | 55 | 40 | 27 | 2000 | 1050 | 48 | 62 | CT |
| 9 | 51-55 | M | 6 | 1 | 56 | 30 | 56 | 38 | 32 | 1275 | 850 | 33 | 46 | CT |
| 10 | 61-65 | F | 9 | 1 | 56 | 15 | 56 | 40 | 29 | 1800 | 700 | 61 | 73 | CT |
| 11 | 46-50 | F | 10 | 1 | 73 | 39 | 73 | 22 | 70 | 290 | 0 | 100 | 47 | CT |
| 12 | 66-70 | M | 13 | 1 | 71 | 36 | 71 | 40 | 44 | 1600 | 800 | 50 | 49 | CT |
| 13 | 66-70 | F | 12 | 1 | 47 | 14 | 47 | 14 | 70 | 1100 | 200 | 82 | 70 | CT |
| 14 | 61-65 | F | 12 | 1 | 31 | 10 | 31 | 36 | -16 | 1450 | 950 | 34 | 68 | CT |
| 15 | 56-60 | F | 15 | 1 | 50 | 15 | 50 | 26 | 48 | 650 | 350 | 46 | 70 | CT |
| 16 | 71-75 | F | 17 | 1 | 59 | 15 | 59 | 14 | 76 | 1400 | 500 | 64 | 75 | CT |
| 17 | 46-50 | M | 23 | 1 | 52 | 29 | 52 | 12 | 77 | 1100 | 100 | 91 | 44 | CT |
| 18 | 56-60 | M | 6 | 1 | 43 | 10 | 43 | 13 | 70 | 2350 | 950 | 60 | 77 | CT |
| 19 | 66-70 | F | 14 | 1 | 60 | 19 | 60 | 17 | 72 | 500 | 300 | 40 | 68 | CT |
| 20 | 61-65 | M | 9 | 1 | 87 | 18 | 67 | 27 | 60 | 2000 | 950 | 53 | 79 | CT |
| 21 | 61-65 | M | 10 | 1 | 29 | 14 | 29 | 32 | -10 | 1350 | 900 | 33 | 52 | CT |
| 22 | 51-55 | F | 6 | 1 | 47 | 18 | 47 | 20 | 57 | 2850 | 250 | 91 | 62 | CT |
| 23 | 56-60 | M | 18 | 1 | 40 | 10 | 40 | 31 | 23 | 1700 | 900 | 47 | 75 | CT |
| 24 | 66-70 | M | 14 | 1 | 51 | 38 | 51 | 23 | 55 | 1100 | 450 | 59 | 25 | CT |
| 25 | 51-55 | M | 5 | 1 | 26 | 3 | 26 | 5 | 81 | 650 | 400 | 38 | 88 | CT |
| 26 | 46-50 | M | 11 | 1 | 46 | 11 | 46 | 17 | 63 | 1100 | 200 | 82 | 76 | CT |

| | | | | | | | | | | | | | | |
|---|---|---|---|---|---|---|---|---|---|---|---|---|---|---|
| 27 | 56-60 | M | 11 | 1 | 43 | 24 | 43 | 22 | 49 | 1000 | 150 | 85 | 44 | CT |
| 28 | 51-55 | M | 19 | 1 | 50 | 11 | 50 | 15 | 70 | 1800 | 600 | 67 | 78 | CT |
| 29 | 51-55 | M | 10 | 1 | 69 | 14 | 69 | 25 | 64 | 2600 | 550 | 79 | 80 | CT |
| 30 | 51-55 | F | 11 | 1 | 55 | 4 | 55 | 14 | 75 | 1320 | 980 | 26 | 93 | CT |
| 31 | 71-75 | M | 18 | 1 | 57 | 16 | 57 | 29 | 49 | 2020 | 688 | 66 | 72 | CT |
| 32 | 46-50 | M | 21 | 1 | 64 | 41 | 64 | 48 | 25 | 2320 | 813 | 65 | 36 | CT |
| 33 | 71-75 | M | 13 | 1 | 44 | 16 | 44 | 45 | -2 | 1680 | 1640 | 2 | 64 | CT |
| 34 | 66-70 | F | 6 | 1 | 64 | 31 | 64 | 23 | 64 | 1033 | 460 | 55 | 52 | CT |
| 35 | 71-75 | M | 9 | 1 | 38 | 35 | 38 | 7 | 82 | 270 | 0 | 100 | 8 | CT |
| 36 | 61-65 | M | 11 | 3 | 38 | 22 | 38 | 19 | 50 | 988 | 105 | 89 | 42 | CT |
| 37 | 66-70 | M | 11 | 1 | 40 | 18 | 40 | 24 | 40 | 2040 | 1880 | 8 | 55 | CT |
| 38 | 46-50 | M | 13 | 1 | 44 | 23 | 44 | 24 | 45 | 1300 | 800 | 38 | 48 | CT |
| 39 | 46-50 | M | 9 | 1 | 37 | 22 | 37 | 22 | 41 | 1000 | 413 | 59 | 41 | CT |
| 40 | 61-65 | M | 10 | 1 | 67 | 18 | 67 | 39 | 42 | 2395 | 832 | 65 | 73 | CT |
| 41 | 61-65 | M | 9 | 1 | 52 | 23 | 52 | 19 | 63 | 916 | 733 | 20 | 56 | CT |
| 42 | 66-70 | M | 7 | 1 | 46 | 26 | 46 | 7 | 85 | 1020 | 443 | 57 | 43 | CT |
| 43 | 66-70 | M | 12 | 1 | 31 | 2 | 31 | 13 | 58 | 1068 | 0 | 100 | 94 | CT |

*Notes:* *Classification into Parkinson's disease (PD) phenotypes according to Schiess et al (2000) [15]: 1= axial-rigid type. 2 = Mixed type, 3 = tremor dominant type. *Abbreviations*: CT, computed tomography; F, female LEDD, levodopa equivalent daily dosage; M, male; Mo., months; MRI, magnetic resonance imaging; Nr., number; postop., postoperative; preop., preoperative; STIM, stimulation; UPDRS-III, Unified Parkinson's Disease Rating Scale – Part III.

**Table S5:** Patient-wise demographic and clinical characteristics of the obsessive-compulsive disorder discovery cohorts.

**GRENOBLE (N = 13)**

| Patient Nr. | Age at Surgery (within range, in years) | Sex | Disease Duration at Surgery (years) | Y-BOCS Baseline | Y-BOCS at 12 Mo. after Surgery | Percent Reduction from Baseline (%) | OCD Subtype | Comorbidities | Postop. Imaging Modality |
|---|---|---|---|---|---|---|---|---|---|
| 1 | 31-35 | M | 13 | 32 | 26 | 19 | checking, repeating | none | MRI |
| 2 | 36-40 | M | 17 | 32 | 28 | 13 | washing, ordering | anankastic personality | MRI |
| 3 | 36-40 | F | 18 | 25 | 12 | 52 | washing | none | MRI |
| 4 | 26-30 | F | 24 | 34 | 30 | 12 | checking | skin picking, hypothyroidia | MRI |
| 5 | 51-55 | F | 25 | 40 | 16 | 60 | washing | none | MRI |
| 6 | 41-45 | M | 26 | 33 | 32 | 3 | checking, hoarding | social phobia | MRI |
| 7 | 51-55 | F | 22 | 33 | 2 | 94 | checking | depression, Minkowski-Chauffard syndrome | MRI |
| 8 | 36-40 | F | 11 | 36 | 6 | 83 | washing | none | MRI |
| 9 | 26-30 | M | 10 | 38 | 22 | 42 | checking, washing | hypersomnia | CT |
| 10 | 46-50 | F | 39 | 35 | 25 | 29 | checking | None | CT |
| 11 | 36-40 | F | 15 | 36 | 30 | 17 | washing | history of eating disorder, history of one seizure, alcoholism, pruritus | MRI |
| 12 | 36-40 | F | 5 | 32 | 3 | 91 | checking | none | MRI |
| 13 | 31-35 | F | 21 | 30 | 14 | 53 | checking | dermatillomania | CT |

**LONDON (N = 6)**

| Patient Nr. | Age at Surgery (within range, in years) | Sex | Disease Duration at Surgery (years) | Y-BOCS Baseline | Y-BOCS after "amSTN only" Phase+ | Percent Reduction from Baseline (%) | OCD Subtype | Comorbidities | Postop. Imaging Modality |
|---|---|---|---|---|---|---|---|---|---|
| 1 | 36-40 | F | 22 | 38 | 32* at 6 mo. | 16 | washing, repeating | none | MRI |
| 2 | 36-40 | M | 22 | 34 | 26 at 6 mo. | 23 | ordering, checking, repeating | body dysmorphic disorder | MRI |
| 3 | 61-65 | M | 30 | 37 | 17 at 6 mo. | 55 | washing, checking, repeating | none | MRI |
| 4 | 36-40 | M | 20 | 38 | 20 at 3 mo. | 47 | washing, checking, ordering | recurrent depression | MRI |
| 5 | 51-55 | M | 23 | 34 | 23 at 3 mo. | 32 | hoarding | recurrent depression | MRI |
| 6 | 41-45 | M | 28 | 36 | 1 at 3 mo. | 97 | checking | generalized anxiety disorder, depression | MRI |

*Notes:* *OCD patients from London were bilaterally implanted with DBS electrodes to two different target sites (four electrodes per patient) – the anteromedial subthalamic nucleus (amSTN) and the ventral capsule/ventral striatum (VC/VS). Patients were randomly assigned to either receive “pure amSTN” for the first three months or from months four to six postoperatively (and “pure VC/VS” for the remainder of the first six months). Model set-up (based on the discovery cohort) was informed on stimulation parameters and corresponding clinical scores taken after the amSTN phase, while model validation was performed based on VC/VS-stimulation related data points. AmSTN stimulation was applied via a Medtronic (MDT) model of type 3389, while MDT 3387 leads were implanted for stimulation of the VC/VS target zone (considered only for model validations, see below). *Abbreviations:* amSTN, anteromedial subthalamic nucleus; CT, computed tomography; F, female; M, male; mo., months; MRI, magnetic resonance imaging; Nr., number; OCD, obsessive-compulsive disorder; postop., postoperative; Y-BOCS, Yale-Brown Obsessive-Compulsive Scale.

**Table S6.** Summary of demographic and clinical patient characteristics within each retrospective model validation cohort.

| **Disease cohort** | **PD** | **OCD** | | |
|---|---|---|---|---|
| | | **Demographic Information** | | |
| **Cohort** | Würzburg | Boston | Cologne | London |
| **Surgical DBS center** | University Hospital Würzburg | Massachusetts General Hospital | University Hospital Cologne | National Hospital for Neurology and Neurosurgery |
| **N (female)** | 32 (10) | 7 (4) | 22 (13) | 6 (1) |
| **Age at time of surgery (mean ± SD; range; in years)** | 58.00 ± 7.73; 46-79 | 38.86 ± 16.80; 21-64 | 42.20 ± 13.38; 21-64 | 45.50 ± 10.52; 37-62 |
| **Disease duration at time of surgery (mean ± SD; range; in years)** | 10.06 ± 4.21; 3-20 | 25.14 ± 14.21; 10-44 | 25.27 ± 12.29; 6-49 | 24.17 ± 3.92; 20-30 |
| | | **Clinical Outcome** | | |
| **Surgical target** | STN | VC/VS | VC/VS | VC/VS |
| **Main clinical outcome assessment** | UPDRS-III [2] (total score; range 0-199, with higher scores corresponding to higher symptom burden) | Y-BOCS [4] (total score; range 0-40, with higher scores corresponding to higher symptom burden) | Y-BOCS [4] (total score; range 0-40, with higher scores corresponding to higher symptom burden) | Y-BOCS [4] (total score; range 0-40, with higher scores corresponding to higher symptom burden) |
| **Time of FU (in months)** | mean: 11.5 ± 3.08 SD; range: 6-22 | 18 | 12 | 3 or 6 (post-VC/VS phase)⁺ |

| | | | | |
|---|---|---|---|---|
| **Comparison basis for calculation of improvement values (baseline to DBS stimulation ON conditions)** | Postoperative ON vs. OFF stimulation (ON dopaminergic medication) | Pre- to postoperative | Pre- to postoperative | Pre- to postoperative |
| **Score at baseline (mean ± SD)** | 45.84 ± 10.93 | 33.57 ± 2.37 | 31.12 ± 4.30 | 36.17 ± 1.83 |
| **Score at time of FU under stimulation ON condition (mean ± SD)** | 24.16 ± 10.65 | 20.29 ± 10.40 | 20.70 ± 7.70 | 17.00 ± 8.74 |
| **Rel. improvement (mean ± SD; in %)** | 46.90 ± 21.11 | 40.13 ± 29.84 | 31.00 ± 20.5 | 52.60 ± 25.51 |
| **Abs. improvement (mean ± SD)** | 21.69 ± 10.96 | 13.29 ± 9.55 | 9.60 ± 6.50 | 19.17 ± 9.39 |
| | | **Imaging and DBS Specifications** | | |
| **Imaging modality (postop.)** | CT (N = 32) | CT (N = 7) | CT (N = 22) | MRI (N = 6) |
| **Electrode models** | BSV (N = 23) or BSV Directed (N = 9) | MDT 3387 (N = 7) | MDT 3387 (N = 3) or MDT 3389 (N = 19) | MDT 3387 (N = 6) |
| **Related citations** | Butenko et al., 2022 [16] | McLaughlin et al., 2021 [17] | Baldermann et al., 2019 [18]; Li et al., 2020 [19]; 2021 [20] | Tyagi et al., 2019 [13] |

*Notes:* $^{+}$OCD patients from London were bilaterally implanted with DBS electrodes to two different target sites (four electrodes per patient) – the anteromedial subthalamic nucleus (amSTN) and the ventral capsule/ventral striatum (VC/VS). Patients were randomly assigned to either receive “pure VC/VS” for the first three months or from months four to six postoperatively (and “pure amSTN” for the remainder of the first six months). Model set-up (based on the discovery cohort) was informed on stimulation parameters and corresponding clinical scores taken after the amSTN phase, while model validation was performed based on VC/VS-stimulation related data points. VC/VS stimulation was applied via a Medtronic (MDT) model of type 3387 (considered only for model validations), while MDT 3389 leads were implanted for stimulation of the amSTN target zone (considered only for the model set-up step). *Abbreviations:* Abs., absolute; BSV, Boston Scientific Vercise; CT, computed tomography; DBS, deep brain stimulation; FU, follow-up; MRI, magnetic resonance imaging; OCD, obsessive-compulsive disorder; PD, Parkinson’s disease; postop., postoperative; Rel., relative; SD, standard deviation; UPDRS-III, Unified Parkinson’s Disease Rating Scale – Part III; Y-BOCS, Yale-Brown Obsessive-Compulsive Scale.

**Table S7:** Patient-wise demographic and clinical characteristics of the Parkinson's disease validation cohorts.

| Pat. Nr. | Age at Surgery (within range, in years) | Sex | Disease Duration at Surgery (years) | UPDRS-III Total Preop. Baseline (OFF Medication) | UPDRS-III Total Preop. Baseline (ON Medication) | UPDRS-III Total Postop. (STIM ON at FU) | UPDRS-III Total Percent Improvement ON vs. OFF STIM at FU (%) | LEDD Baseline (OFF DBS) | Time of FU (months) | Postop. Imaging Modality |
|---|---|---|---|---|---|---|---|---|---|---|
| **WÜRZBURG (N = 32)** | | | | | | | | | | |
| 1 | 56-60 | M | 12 | 48 | 9 | 17 | 65 | 81 | 12 | CT |
| 2 | 56-50 | F | 9 | 62 | 27 | 38 | 39 | 56 | 12 | CT |
| 3 | 61-65 | F | 12 | 42 | 22 | 17 | 60 | 48 | 12 | CT |
| 4 | 46-50 | M | 18 | 52 | 34 | 34 | 35 | 35 | 10 | CT |
| 5 | 56-60 | M | 7 | 56 | 9 | 17 | 70 | 84 | 16 | CT |
| 6 | 56-60 | M | 12 | 49 | 20 | 24 | 51 | 59 | 12 | CT |
| 7 | 71-75 | M | 10 | 40 | 20 | 28 | 30 | 50 | 22 | CT |
| 8 | 56-60 | F | 11 | 44 | 10 | 8 | 82 | 77 | 12 | CT |
| 9 | 61-65 | M | 5 | 45 | 12 | 27 | 40 | 73 | 12 | CT |
| 10 | 51-55 | F | 3 | 61 | 34 | 39 | 36 | 44 | 12 | CT |
| 11 | 61-65 | M | 11 | 45 | 11 | 32 | 29 | 76 | 12 | CT |
| 12 | 51-55 | F | 6 | 31 | 10 | 16 | 48 | 68 | 12 | CT |
| 13 | 46-50 | M | 12 | 47 | 20 | 36 | 23 | 57 | 12 | CT |
| 14 | 46-50 | M | 7 | 44 | 20 | 14 | 68 | 55 | 13 | CT |
| 15 | 46-50 | M | 7 | 38 | 13 | 9 | 76 | 66 | 12 | CT |
| 16 | 66-70 | M | 8 | 45 | 5 | 48 | -7 | 89 | 13 | CT |
| 17 | 51-55 | M | 9 | 50 | 13 | 26 | 48 | 74 | 12 | CT |
| 18 | 46-50 | M | 9 | 44 | 17 | 21 | 52 | 61 | 12 | CT |
| 19 | 61-65 | M | 6 | 39 | 18 | 35 | 10 | 54 | 12 | CT |
| 20 | 51-55 | M | 8 | 16 | 8 | 10 | 38 | 50 | 12 | CT |
| 21 | 56-60 | M | 10 | 52 | 38 | 24 | 54 | 27 | 12 | CT |
| 22 | 56-60 | F | 18 | 41 | 14 | 22 | 46 | 66 | 12 | CT |

| | | | | | | | | | | |
|---|---|---|---|---|---|---|---|---|---|---|
| 23 | 66-70 | F | 17 | 45 | 18 | 7 | 84 | 60 | 12 | CT |
| 24 | 61-65 | M | 9 | 44 | 23 | 19 | 57 | 48 | 12 | CT |
| 25 | 76-80 | M | 5 | 42 | 26 | 25 | 40 | 38 | 12 | CT |
| 26 | 61-65 | F | 16 | 39 | 14 | 28 | 28 | 64 | 12 | CT |
| 27 | 56-60 | M | 8 | 42 | 33 | 39 | 7 | 21 | 12 | CT |
| 28 | 46-50 | F | 12 | 40 | 19 | 20 | 50 | 53 | 6 | CT |
| 29 | 56-60 | M | 20 | 85 | 42 | 42 | 51 | 51 | 6 | CT |
| 30 | 56-60 | M | 13 | 51 | 25 | 20 | 61 | 51 | 6 | CT |
| 31 | 51-55 | F | 6 | 41 | 7 | 14 | 66 | 83 | 6 | CT |
| 32 | 61-65 | M | 6 | 47 | 22 | 17 | 64 | 53 | 6 | CT |

*Abbreviations*: CT, computed tomography; F, female; FU, follow-up; LEDD, levodopa equivalent daily dosage; M, male; MRI, magnetic resonance imaging; Nr., number; postop., postoperative; preop., preoperative; STIM, stimulation; UPDRS-III, Unified Parkinson's Disease Rating Scale – Part III.

**Table S8:** Patient-wise demographic and clinical characteristics of the obsessive-compulsive disorder validation cohorts.

**BOSTON (N = 7)**

| Patient Nr. | Age at Surgery (within range, in years) | Sex | Disease Duration at Surgery (years) | Y-BOCS Baseline | Y-BOCS at 18 Mo. after Surgery | Percent Reduction from Baseline (%) | OCD Subtype | Comorbidities | Postop. Imaging Modality |
|---|---|---|---|---|---|---|---|---|---|
| 1 | 56-60 | M | 41 | 32 | 0 | 100 | doubt, checking | n/a | CT |
| 2 | 21-25 | F | 13 | 35 | 16 | 54 | doubt, checking | n/a | CT |
| 3 | 61-65 | F | 44 | 32 | 21 | 34 | contamination, cleaning | n/a | CT |
| 4 | 21-25 | M | 10 | 33 | 29 | 12 | doubt, checking | n/a | CT |
| 5 | 26-30 | F | 11 | 34 | 22 | 35 | doubt, checking | n/a | CT |
| 6 | 41-45 | F | 27 | 31 | 22 | 29 | contamination, cleaning | n/a | CT |
| 7 | 36-40 | M | 30 | 38 | 32 | 16 | contamination, cleaning | n/a | CT |

**COLOGNE (N = 22)**

| Patient Nr. | Age at Surgery (within range, in years) | Sex | Disease Duration at Surgery (years) | Y-BOCS Baseline | Y-BOCS at 12 Mo. after Surgery | Percent Reduction from Baseline (%) | OCD Subtype | Comorbidities | Postop. Imaging Modality |
|---|---|---|---|---|---|---|---|---|---|
| 1 | 36-40 | M | 24 | 30 | 8 | 73 | magical thinking, incompleteness, responsibility | recurrent major depressive disorder (moderate) | CT |
| 2 | 31-35 | M | 15 | 34 | 34 | 0 | washing, contamination | none | CT |
| 3 | 36-40 | M | 27 | 33 | 24 | 27 | washing, contamination, symmetry, catastrophic fears | none | CT |
| 4 | 51-55 | M | 6 | 34 | 12 | 65 | washing, controlling, contamination fears, aggresive thought | none | CT |
| 5 | 26-30 | M | 22 | 27 | 14 | 48 | contamination, aggressive impulses, symmetry | none | CT |
| 6 | 21-25 | M | 18 | 25 | 20 | 20 | repetition, keeping order, mental exactness, checking | none | CT |
| 7 | 21-25 | F | 15 | 35 | 23 | 34 | bodydysmorphobia, checking, magical thinking | hypochondriasis | CT |
| 8 | 46-50 | M | 24 | 32 | 22 | 31 | somatic obsessions, contamination fears | none | CT |
| 9 | 31-35 | M | 19 | 37 | 34 | 8 | cleanliness, washing, contamination fears, digust | none | CT |

| | | | | | | | | | |
|---|---|---|---|---|---|---|---|---|---|
| 10 | 46-50 | F | 19 | 28 | 13 | 54 | checking, aggressive fears, responsibility | none | CT |
| 11 | 56-60 | F | 49 | 36 | 22 | 39 | aggressive thoughts/impulses, mental urges | recurrent major depressive disorder (severe without psychotic features) | CT |
| 12 | 26-30 | F | 6 | 26 | 16 | 38 | aggressive impulses, checking, doubting | none | CT |
| 13 | 61-65 | F | 20 | 25 | 18 | 28 | catastrophic fears, responsibility | anxiety disorder (not otherwise specified); recurrent major depressive disorder (moderate) | CT |
| 14 | 56-60 | M | 35 | 35 | 25 | 29 | catastrophic fears, responsibility | recurrent major depressive disorder (moderate) | CT |
| 15 | 26-30 | M | 18 | 33 | 22 | 33 | aggressive impulses, responsibility | recurrent major depressive disorder (severe without psychotic features) | CT |
| 16 | 56-60 | M | 44 | 25 | 20 | 20 | aggressive impulses, responsibility | recurrent major depressive disorder (severe without psychotic features) | CT |
| 17 | 36-40 | F | 29 | 30 | 28 | 7 | magical thinking, incompleteness, mental urges | recurrent major depressive disorder (moderate) | CT |
| 18 | 41-45 | F | 34 | 26 | 21 | 19 | aggressive impulses, responsibility, checking | none | CT |
| 19 | 41-45 | F | 39 | 37 | 27 | 27 | mental rituals/urges, magical thinking | none | CT |
| 20 | 51-55 | M | 42 | 34 | 29 | 15 | contamination, cleanliness | none | CT |
| 21 | 36-40 | M | 11 | 36 | 36 | 0 | responsibility, catarstrophic fears, checking | recurrent major depressive disorder (moderate) | CT |
| 22 | 56-60 | F | 40 | 32 | 11 | 66 | somatic obsessions, sexual obsessions | sedative, hypnotic, or anxiolytic dependence | CT |

**LONDON (N = 6)**

| Patient Nr. | Age at Surgery (within range, in years) | Sex | Disease Duration at Surgery (years) | Y-BOCS Baseline | Y-BOCS after "VC/VS only" Phase$^{+}$ | Percent Reduction from Baseline (%) | OCD Subtype | Comorbidities | Postop. Imaging Modality |
|---|---|---|---|---|---|---|---|---|---|
| 1 | 36-40 | F | 22 | 38 | | 16 | washing, repeating | none | MRI |
| 2 | 36-40 | M | 22 | 34 | | 23 | ordering, checking, repeating | body dysmorphic disorder | MRI |

| | | | | | | | | |
|---|---|---|---|---|---|---|---|---|
| 3 | 61-65 | M | 30 | 37 | 55 | washing, checking, repeating | none | MRI |
| 4 | 36-40 | M | 20 | 38 | 47 | washing, checking, ordering | recurrent depression | MRI |
| 5 | 51-55 | M | 23 | 34 | 32 | hoarding | recurrent depression | MRI |
| 6 | 41-45 | M | 28 | 36 | 97 | checking | generalized anxiety disorder, depression | MRI |

*Notes:* +OCD patients from London were bilaterally implanted with DBS electrodes to two different target sites (four electrodes per patient) – the anteromedial subthalamic nucleus (amSTN) and the ventral capsule/ventral striatum (VC/VS). Patients were randomly assigned to either receive “pure VC/VS” for the first three months or from months four to six postoperatively (and “pure amSTN” for the remainder of the first six months). Model set-up (based on the discovery cohort) was informed on stimulation parameters and corresponding clinical scores taken after the amSTN phase, while model validation was performed based on VC/VS-stimulation related data points. VC/VS stimulation was applied via a Medtronic (MDT) model of type 3387 (considered only for model validations), while MDT 3389 leads were implanted for stimulation of the amSTN target zone (considered only for the model set-up step). *Abbreviations:* CT, computed tomography; F, female; M, male; mo., months; MRI, magnetic resonance imaging; Nr., number; OCD, obsessive-compulsive disorder; postop., postoperative; VC/VS, ventral capsule/ventral striatum; Y-BOCS, Yale-Brown Obsessive-Compulsive Scale.

**Table S9:** Peak voxel coordinates of subthalamic sweet spots and of disease-specific cortical sites of interconnection with sweet tracts.

| | | **DYT** | | | **TS** | | | **PD** | | | **OCD** | | |
|---|---|---|---|---|---|---|---|---|---|---|---|---|---|
| | | **(x** | **y** | **z)** | **(x** | **y** | **z)** | **(x** | **y** | **z)** | **(x** | **y** | **z)** |
| **Cortical sites of interconnection with sweet tracts (in mm)** | Left | (-22.00 | -20.00 | 72.00) | (-3.32 | -29.65 | -45.59) | (-14.00 | 2.00 | 70.00) | (-8.23 | -10.29 | -16.50) |
| | Right | (23.50 | -18.75 | 72.00) | (10.00 | -12.00 | -16.50) | (10.00 | 2.00 | 70.00) | (7.88 | -9.50 | -16.50) |
| **Subthalamic sweet spots (in mm)** | Left | (-14.72 | -14.38 | -4.10) | (-12.96 | -11.08 | -6.30) | (-12.08 | -13.94 | -6.74) | (-8.78 | -11.52 | -9.60) |
| | Right | (14.54 | -13.50 | -4.30) | (13.00 | -9.98 | -6.08) | (11.90 | -13.28 | -6.74) | (8.38 | -10.86 | -9.38) |

*Abbreviations:* DYT, dystonia; mm, millimeters; OCD, obsessive-compulsive disorder; PD, Parkinson’s disease; TS, Tourette’s syndrome.

**Table S10:** Overlap between connected streamlines shared among disorders and disease-specific sweet streamlines.

| | DYT | PD | TS | OCD |
|---|---|---|---|---|
| **Overlap (%)** | 4.41 | 2.21 | 1.75 | 5.42 |
| **Four-sample test for equality of proportions** | p < 2.2e-16 | | | |
| Pairwise test for equality of proportions | | | | |
| **DYT** | | | | |
| **PD** | p < 2.2e-16 | | | |
| **TS** | p < 2.2e-16 | p < 2.84e-14 | | |
| **OCD** | p < 1.35e-11 | p < 2.2e-16 | p < 2.2e-16 | |

*Abbreviations:* DYT, dystonia; mm, millimeters; OCD, obsessive-compulsive disorder; PD, Parkinson's disease; TS, Tourette's syndrome.

**Table S11:** Patient-wise demographic and clinical characteristics of patient cases for prospective model validations.

**PD**

| Age at Surgery (within range, in years) | Sex | Disease Duration at Surgery (years) | PD Phenotype | UPDRS-III Total Postop. (STIM OFF, Med. OFF at 3 Mo.) | UPDRS-III Total Postop. (STIM ON under Clinical Settings, Med. OFF at 3 Mo.) | UPDRS-III Total Postop. (STIM ON under Model-Based Settings, Med. OFF at 3 Mo.) | UPDRS-III Total Improvement ON vs OFF STIM at 3 Mo. postop. (under Clinical Settings, Med. OFF , %) | UPDRS-III Total Improvement ON vs OFF STIM at 3 Mo. postop. (under Model-Based Settings, Med. OFF , %) | Electrode Model | Clinical Settings | Model-Based Settings | Postop. Imaging Modality |
|---|---|---|---|---|---|---|---|---|---|---|---|---|
| ***PD patient implanted to the STN at Würzburg University Hospital (Würzburg, Germany)*** | | | | | | | | | | | | |
| 66-70 | M | 9 | Akinetik-rigid | 35 | 14 | 10 | 60 | 71 | BSV Directed | Left hemisphere: C+, 1- (60%), 2- (40%) / 2.4 mA / 60 µs / 130 Hz; Right hemisphere.: C+, 1- (20%), 2- (20%), 3- (3%), 4- (57%) / 2.6 mA / 60 µs / 130 Hz | Left hemisphere: C+, 4- (70%), 7- (30%) / 3 mA / 60 µs / 130 Hz; Right hemisphere: C+, 7- (30%), 8- (70%) / 3 mA / 60 µs / 130 Hz | CT |

| OCD | | | | | | | | | | | | | | |
|---|---|---|---|---|---|---|---|---|---|---|---|---|---|---|
| **Age at Surgery (within range, in years)** | **Sex** | **Disease Duration at Surgery (years)** | **OCD Phenotype** | **Comor-bidities** | **Previous treatments** | **Y-BOCS Total Preop.** | **Y-BOCS Total Postop. (STIM ON under Clinical Settings at 1 Mo. Postop.)** | **Y-BOCS Total Postop. (STIM ON under Model-Based Settings at 1 Mo. Postop.)** | **Y-BOCS Total Improvement Preop. vs. STIM ON at 1 Mo. Postop. (under Clinical Settings, %)** | **Y-BOCS Total Improvement Preop. Vs. STIM ON at 1 Mo. Postop. (under Model-Based Settings, %)** | **Electrode Model** | **Clinical Settings** | **Model-Based Settings** | **Postop. Imaging Modality** |
| ***OCD patient implanted to the VC/VS region at Massachusetts General Hospital (Boston, USA)*** | | | | | | | | | | | | | | |
| 21-25 | F | 13 | obsessions about food and water intake, compulsions involving ingestion events; skin picking | major depressive disorder | 11 different medication trials (including multiple SSRIs, clomipramine, and ketamine), intensive ERP, ECT | 35 | 29 | 22 | 17 | 37 | MDT 3387 | Left hemisphere: C+, 1- , 2- / 3 mA / 140 µs / 130 Hz; Right hemisphere: C+, 9- / 2.4 mA / 140 µs / 130 Hz; | Left hemisphere: C+, 2- / 4.2 mA / 80 µs / 130 Hz; Right hemisphere: C+, 10- / 3.4 mA / 80 µs / 130 Hz; | CT |
| ***OCD patient implanted to the STN at Clínica de Dor e Funcional (São Paulo, Brazil)*** | | | | | | | | | | | | | | |
| 31-35 | M | 14 | compulsion of noting words unknown to the patient and transcribing their meaning from dictionaries; death-related intrusions leading to compulsive religious rituals | apathy, major depressive disorder | 48 sessions of ECT, CBT, biofeedback, mindfulness training, 3 different protocols of TMS with over 50 sessions | 26 | - | 6 | - | 77 | BSV | - | Left hemisphere: C+, 1- (100%) / 3 V / 90 µs / 130 Hz; Right hemisphere: C+, 9- (100%) / 3 V / 90 µs / 130 Hz; | CT |

*Abbreviations*: BSV, Boston Scientific Vercise; CBT, cognitive behavioral therapy; CT, computed tomography; ECT, electroconvulsive therapy; ERP, exposure and response prevention therapy; F, female; Hz, Hertz; M, male; mA, milliampere; Mo., months; MDT, Medtronic; postop., postoperative; preop., preoperative; postop., postoperative; SSRI, selective serotonin inhibitor; STIM, stimulation; STN, subthalamic nucleus; TMS, transcranial magnetic stimulation; UPDRS III, Unified Parkinson's Disease Rating Scale – Part III; VC/VS, ventral capsule/ventral striatum; V, volt; Y-BOCS, Yale-Brown Obsessive-Compulsive Scale.
